# Estimating the impact of decreasing vaccination response times for outbreaks of vaccine-preventable diseases in low and middle-income countries

**DOI:** 10.1101/2025.02.04.25321698

**Authors:** Dominic Delport, Alina M. Muellenmeister, Jane Greig, Romesh G. Abeysuriya, Nick Scott

## Abstract

**Background:** Globally, infectious disease outbreaks cause a major health and economic burden. For infectious diseases which have existing vaccines, reactive vaccination programs during an outbreak are a powerful tool for reducing disease transmission — preventing infections and deaths. However, despite awareness, monitoring, and accessible stockpiles of vaccines, it can take months for vaccines to be delivered once outbreaks are detected in low-resource settings.

The 7-1-7 targets are gaining traction as measurable targets for assessing a country’s outbreak readiness. The targets are outbreak detection within seven days of emergence; notification to health authorities within one day; and key early response actions commenced within another seven days. For outbreaks of measles, cholera, yellow fever, and meningococcal meningitis, we aim to estimate the impact of initiating outbreak response immunisation (ORI) within 15 days of outbreak emergence, relative to the mean ORI response time for each disease in low and middle-income countries (LMICs) since 2000.

While only one component of outbreak response, initiating ORI within 15-days of outbreak emergence aligns with 7-1-7 targets and supports outbreak containment.

**Methods:** Using calibrated agent-based models for four diseases, a status-quo and series of ‘Faster response’ scenarios were compared for simulated outbreaks of each disease, with a 15-day ORI response time as the minimum.

**Results:** In a synthetic model population, a 15-day ORI response could avert: 80% of cases from cholera outbreaks relative to a historical response time of 105 days; 35% of cases from meningococcal meningitis outbreaks relative to a historical response time of 75 days; 0 – 35% of cases from yellow fever outbreaks relative to a historical response time of 105 days (depending on routine vaccine coverage and environmental suitability); and 0 – 55% of cases from measles outbreaks relative to a historical response time of 120 days (depending on routine vaccine coverage).

**Conclusions:** Improvements made to ORI response time could reduce disease burden and decrease the risk of large outbreaks of vaccine-preventable diseases in LMICs. Efforts to improve ORI timeliness should be prioritised to higher risk settings, and it was clear that even a slow vaccination response was beneficial relative to no response at all.

## Background

The COVID-19 pandemic and 2013 – 2016 West African Ebola epidemic are two stark, recent examples of how vital it is to have effective systems of detection, notification, and response to combat disease outbreaks, and how disruptive and devastating they can become if the systems are unprepared. The 7-1-7 targets developed by Resolve to Save Lives are gaining traction as metrics for assessing a country’s outbreak readiness; they propose that outbreaks be detected within seven days; notification occur within one day; and key early response actions be started within another seven days.[1] One of the seven early response actions is to initiate appropriate public health countermeasures in affected communities, which can include starting immunisation. Relatively few countries have adopted these metrics yet, and of 41 public health events assessed across five low and middle-income countries (LMICs) between 2018 – 2022, only 27% fully satisfied the 7-1-7 targets.[2] Logistical challenges (such as cold-chain constraints for vaccines), weak healthcare infrastructure, and understaffed workforces are some of the primary hurdles to overcome when improving outbreak response timeliness.[3] Ideally, use of the 7-1-7 targets with bottleneck and enabler identification can make every outbreak an opportunity for public health system performance improvement, and are a tool for clear communication of improvement to stakeholders.[2]

Outbreaks of vaccine-preventable diseases (VPDs) frequently occur in LMICs, often requiring outbreak response immunisation (ORI) programs and other containment interventions.[4, 5] Immunisation is a highly effective tool available for prevention of and response to VPD outbreaks.[6–8] However, despite awareness, monitoring, and accessible stockpiles of vaccines,[9] it can still take weeks or months for vaccines to be delivered once disease outbreaks are detected.[4, 8, 10, 11] Governments and organisations which fund outbreak response programs seek metrics by which to assess the timeliness of these responses, as a measure of success and to identify areas for improvement, but in LMICs the lack of consistent reporting makes benchmarking and target setting challenging.[2, 10, 12–14] The Sustainable Development Goals (SDGs) Target 3.3 calls for the end of epidemics of very high-burden infectious diseases such as HIV, tuberculosis, and malaria; and the combatting of communicable diseases more broadly.[15] Unfortunately, there is a great deal of work left to do to reduce the burden of infectious diseases to the levels indicated by SDG 3.3, and the current approach does not appear to be working fast enough.[16]

It is expected that non-pharmaceutical interventions (e.g., contact tracing or public health messaging) can typically be initiated quickly in outbreak contexts by utilising existing workforces and systems, but accessing vaccine stockpiles can be slowed by logistical challenges (e.g., physical distance or cold-chain constraints). Therefore, there is clear scope for improvement in a critical element of outbreak response, and mathematical models can be used to quantify the impact of improving ORI timeliness for policy makers.[17]

Previous modelling work found that for outbreaks of four diseases — measles, meningococcal meningitis, yellow fever, and cholera — the time between outbreak detection and immunisation response had considerable impact on the proportion of disease burden averted by ORI.[8] Faster immunisation responses (typically around one month) consistently prevented a higher proportion of disease burden than outbreaks with slower responses (typically three to six months).[8] Despite this, the typical immunisation response time for outbreaks of these diseases in LMICs is two to four months, with the Immunization

Agenda 2030 scorecard indicating that less than 30% of outbreaks had a timely detection and response between 2021 – 2023.[18] To our knowledge, there are a limited number of publications investigating the relationship between ORI timeliness and disease burden for outbreaks of VPDs using mathematical modelling techniques,[19–29] and none which estimate the impacts of achieving it within a period that supports 7-1-7 targets.

In this study, we estimate the impact of improving ORI timeliness for outbreaks of measles, cholera, yellow fever, and meningococcal meningitis in LMICs, down to a minimum ORI response time of 15 days to assess the incremental benefits of ORI timeliness gains in the context of the 7-1-7 targets. For the purposes of this analysis, we considered ‘ORI response time’ to be the time between an outbreak beginning and the initiation of an immunisation response delivered to the population at-risk, as this captures the 15-day period covered by the 7-1-7 targets.[1]

## Methods

### Data

We used a previously formulated dataset, which contains relevant epidemiological and programmatic response data for 51 measles, 40 cholera, 24 meningococcal meningitis, and 88 yellow fever outbreaks in LMICs between 2000 – 2023 to calibrate our disease models.[8, 30] The mean ORI response time and daily vaccination rate from the set of outbreaks from each disease were used to inform Baseline scenario parameters. The dataset is available in a Zenodo repository.[30]

### Models

We used previously developed agent-based models for measles, cholera, meningococcal meningitis (serogroups C, W, X, Y), and yellow fever,[8] which were built using the Starsim toolbox.[31] Starsim is an open-source agent-based framework developed by the Institute for Disease Modeling at the Bill and Melinda Gates Foundation in collaboration with the Burnet Institute and other partners.[31, 32] The framework is written in Python and allows for rapid development of agent-based disease models that can be customised to arbitrary levels of detail. The Starsim framework provides several key components that can be used to build models:

- Disease ‘modules’: capturing different health states and disease progression within individuals.
- Contact networks: to simulate interactions between agents that can give rise to transmission. Multiple networks can be used to allow for different mechanisms of transmission and different settings (e.g., household networks, school networks, random community contacts).
- Interventions: allow changes to be made to the model during simulations, such as delivering vaccines, performing contact tracing, or changing transmission levels due to non-pharmaceutical interventions brought in once an outbreak is detected.

We used the Starsim framework to build the models in this study to provide a single common software and analysis architecture for all models, while simultaneously having the flexibility to account for very different modes of transmission across the diseases being modelled.

#### Disease models

Each of the four disease models (i.e., Starsim disease modules) handles transmission events differently:

- The measles model simulates transmission through random human-human contacts via networks generated randomly at each time step;
- The meningococcal meningitis model simulates transmission through random human-human contacts (regenerated each time step) and static, fully connected household networks;
- The cholera model simulates transmission through random contacts with tainted water sources (calculated each time step based on cholera prevalence in the water) and human-human contact in static, fully connected household networks;
- The yellow fever model simulates transmission through random mosquito-human and human-mosquito contacts (calculated each time step based on prevalence in both populations).

Table 1 contains a summary of key model characteristics for each disease considered, and Table 2 describes the key parameters and assumptions for each disease model and vaccine used in the ORI programs. Figures S1, S7, S11, and S16 in ‘Additional file 1’ show the schematic representation of each disease model. Additional details of each model can be found in the ‘Model overview’ sections of Sections 2 – 5 in ‘Additional file 1’.

**Table 1:** Summary of key model characteristics for each disease considered. *Pentavalent conjugate vaccines for serogroups ACWYX were prequalified and recommended for inclusion in routine vaccination programs by the WHO for countries in the African meningitis belt,[33, 34] however uptake is not yet widespread and has not been captured in this analysis.[35, 36]

|  | Modelled population | Transmission | Routine/preventative vaccination | Asymptomatic cases | Non-pharmaceutical interventions | Mediating parameters for simulation |
| --- | --- | --- | --- | --- | --- | --- |
| Measles | 0 – 5 | Direct | Y | N | N | Routine vaccine coverage |
| Cholera | All ages | Direct + Environmental (water) | N | Y | Y |  |
| Yellow fever | All ages | Vector-borne | Y | Y | N | Routine vaccine coverage, environmental transmission level |
| Meningitis | All ages | Direct (inc. asymptomatic) | N (MenA outbreaks not included)* | Y | N | Asymptomatic carrier prevalence |

**Table 2:**
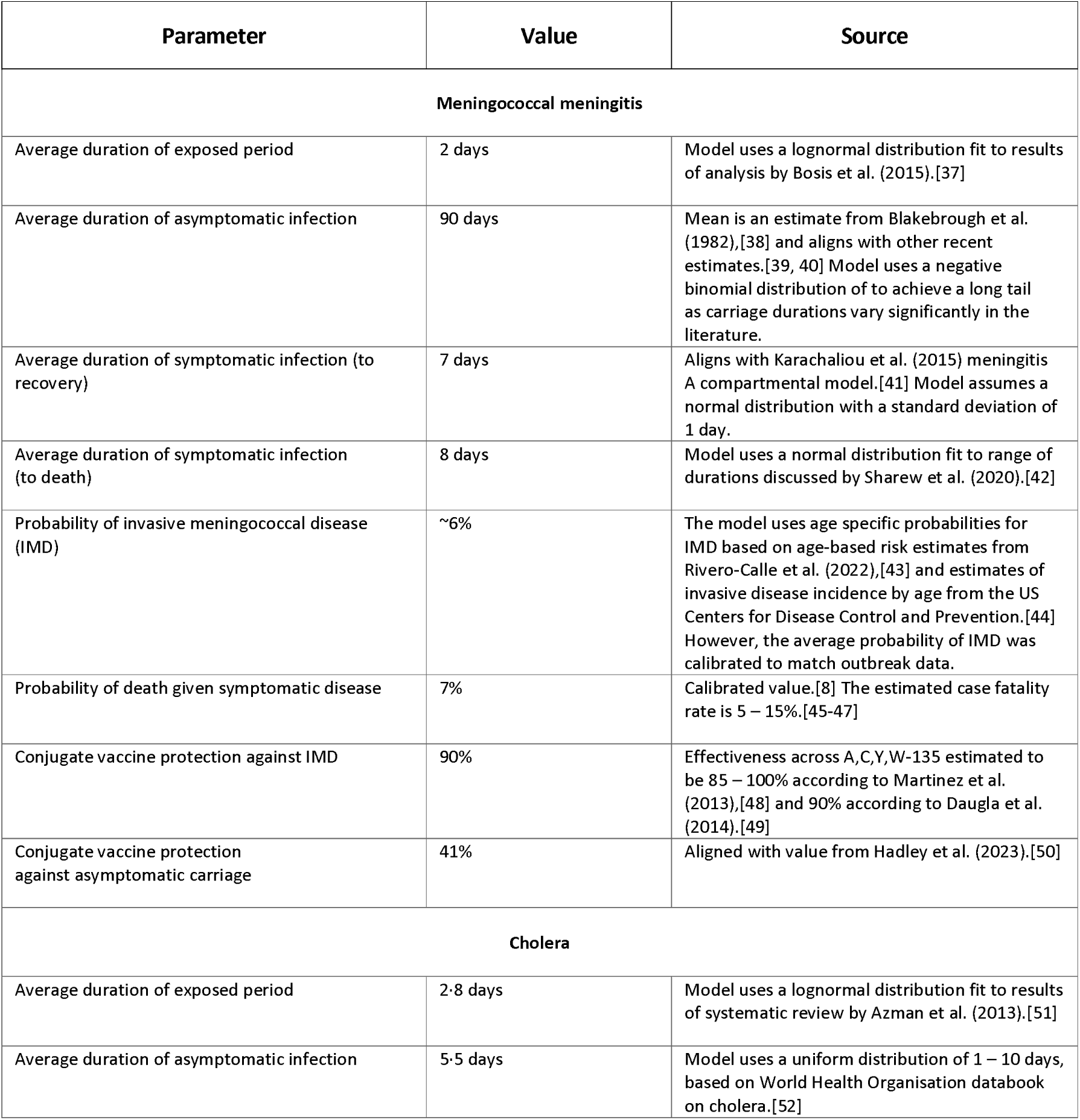
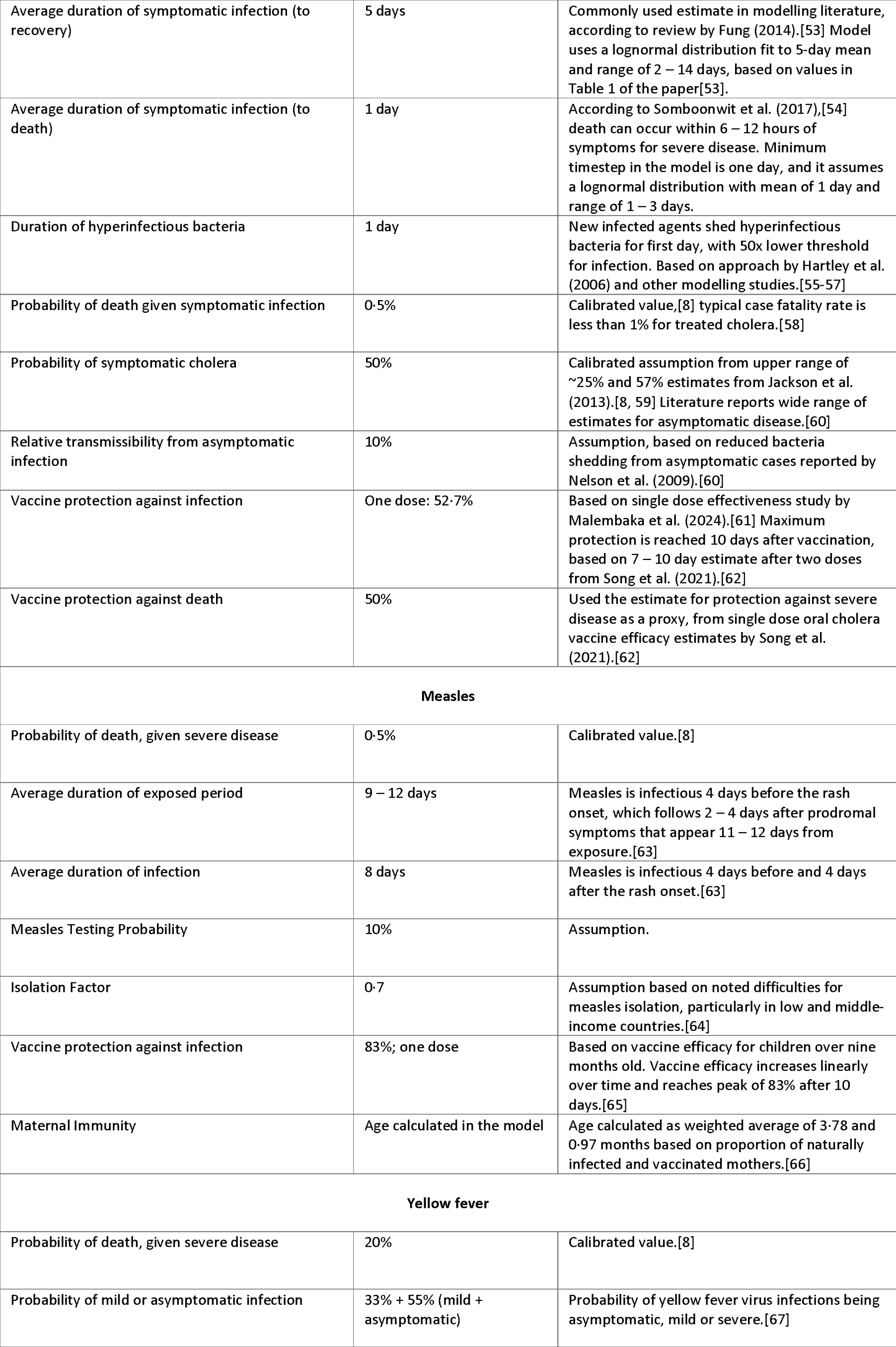
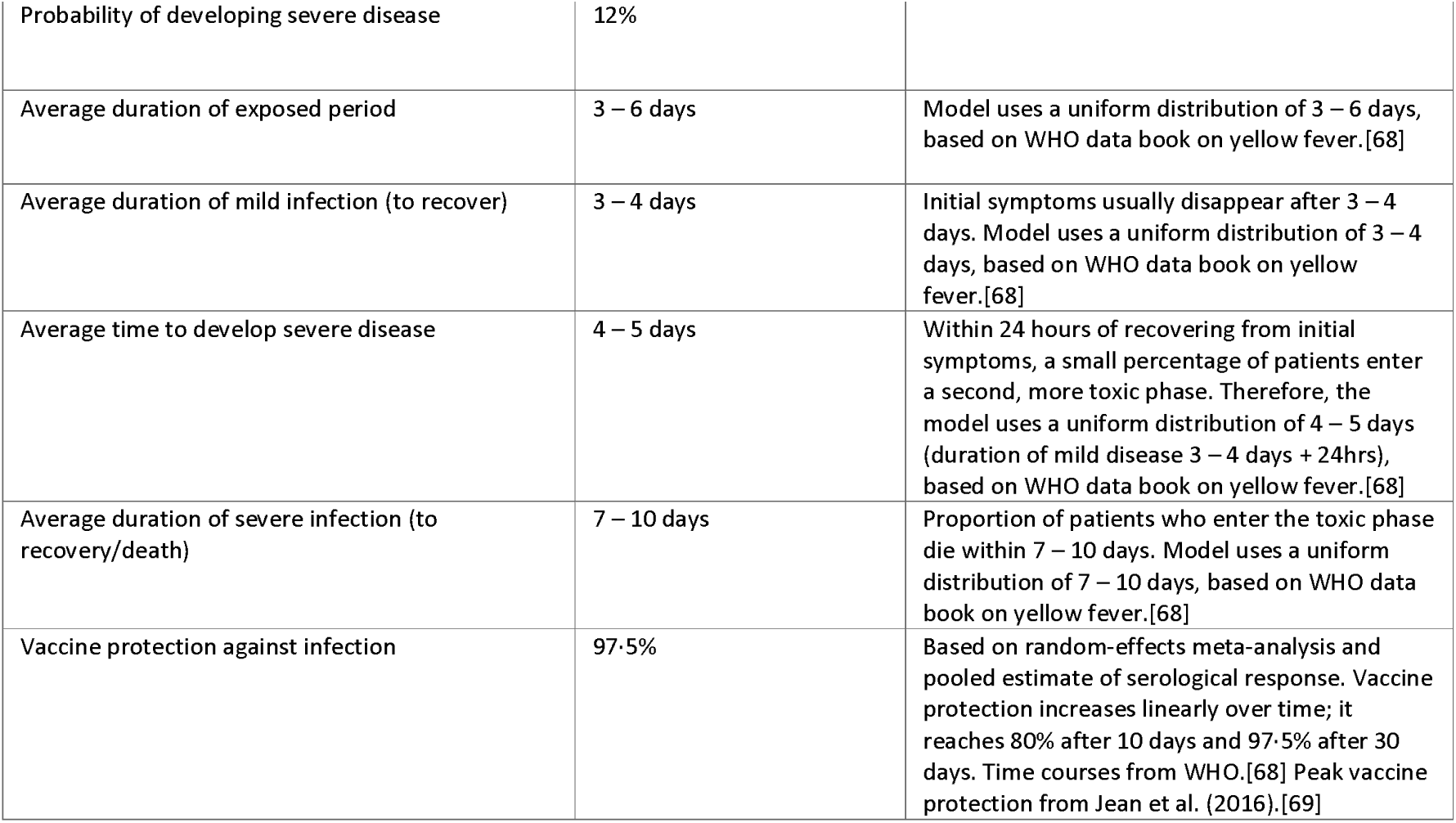
Disease model and vaccine parameters for meningococcal meningitis, cholera, measles, and yellow fever.

#### Contact networks

Each network in the models can have a different structure, with agents being connected to their contacts randomly or agents being grouped into disconnected household clusters.

Each agent in the models (except the yellow fever model, which has no human-human transmission) has a specified number of contacts in at least one network. For those who have a non-zero number of contacts in a particular network, if the contact network is random, then their number of contacts per day is drawn from a Poisson distribution with a mean of 11, 9, and 3·5 for meningococcal meningitis, cholera, and measles, respectively. For each disease, these means were estimated from Prem et al. (2017),[70] with values averaged over all non-household contact rates and all ages, for all countries with outbreaks in the outbreak dataset.[30]

If the contact network is a household, then the size of each cluster is drawn from a synthetic household size distribution generated by summing over household size data (from the United Nations, Department of Economic and Social Affairs, Population Division)[71] for each country with an outbreak included in the outbreak dataset.[30]

#### Vaccine characteristics

For each modelled disease there was a vaccine intervention included in the simulations, for use during ORI. The vaccine effects varied by disease (with relevant characteristics and efficacies detailed in Table 2), but all acted to at least moderately protect vaccinated agents in the outbreak simulations against infection. For all vaccines, the implementation within the models assumed a linear increase in protection against infection over time, achieving 52.7%, 83.0%, and 97.5% for cholera, measles, and yellow fever, respectively. Both the polysaccharide and conjugate meningitis vaccines provide 90% protection against invasive disease, but the conjugate vaccines also provide 41% protection against asymptomatic infection. The times to maximal protection were 10 days for the cholera and measles vaccines[62, 65]; eight days for the polysaccharide and conjugate meningitis vaccines[48]; and 30 days for the yellow fever vaccine[68].

### Model populations

The disease models were used to run simulations of future outbreaks in a synthetic model population intended to be representative of LMIC contexts where outbreaks have typically occurred, with ORI response parameters based on the mean values from the outbreak dataset (unless varied as a part of a scenario analysis).[30] Each disease model simulated a population of 50,000 agents, intended to be large enough to capture widespread transmission in an outbreak area but small enough to limit computational intensity. The age distributions of the agents in each model were generated by summing over data from countries where outbreaks occurred in our dataset, informed by single-year age distribution data from United Nations, Department of Economic and Social Affairs, Population Division.[71]

### Calibration

The model calibration was unchanged from previous work, which has been published in detail elsewhere.[8] In brief, each model was calibrated to a set of historical outbreaks in LMICs which received ORI as a part of the outbreak response. The calibration ensured that model simulations could reproduce the range of outbreak sizes and durations observed in the dataset.[30]

The calibration method controlled for epidemiological and ORI programmatic parameters such as response time, daily vaccination rate, routine vaccine coverage, the mosquito-to-human transmission modifier, or the asymptomatic carriage prevalence.[8] The transmission modifier parameter was used to capture the impact of temperature and rainfall on mosquito population size and biting behaviours on yellow fever transmission in outbreak settings. Asymptomatic carriage prevalence was a parameter used to modify the initial growth rate of an outbreak as an epidemic season begins in meningococcal meningitis outbreak settings. Further details about the calibration process have been published previously,[8] and a summary can be found in Section 1 in ‘Additional file 1’. Further, additional details about the specific calibrations for each model are available in Sections 2 – 5 in ‘Additional file 1’.

Unless varied during a scenario (i.e., response time, achieved coverage, or vaccination rate) almost all disease and intervention parameters in the four models are informed by estimates from the literature or were determined during calibration (detailed in Table 2).

The only three exceptions to this were the routine vaccine coverage, asymptomatic carriage prevalence, or mosquito-to-human transmission modifier parameters, which were used to investigate outbreaks in settings with different levels of risk. The primary parameters for each disease which were determined during calibration were the overall transmissibility; the probability of death, given severe infection; and the number of seed infections (except for meningitis, which was triggered by asymptomatic carriage in the population). For the cholera model, the probability of symptomatic disease was also estimated during calibration.

### Outbreak detection and response

For cholera, measles, and yellow fever, the threshold for outbreak declaration in the models was defined as one case. According to the WHO, measles outbreaks are typically declared if there are five or more epidemiologically linked cases.[72] Cholera outbreaks are considered ‘suspected’, ‘probable’, or ‘confirmed’ for a series of thresholds, beginning with two suspected cases or one suspected case with a positive rapid diagnostic test.[73] The WHO also states that outbreaks are declared in the ‘presence of at least one confirmed case of yellow fever’.[74] As the models do not differentiate between suspected and confirmed cases, the assumption for the threshold is optimistic for measles; however for cholera and yellow fever, the thresholds used in the model are identical or similar to declaration thresholds stated by the WHO. The case is assumed to be detected on the first day of the simulation, a one-day delay is assumed for notification, and the ORI begins after a further (N-2)-days, where N is the response time used for each scenario.

For meningococcal meningitis, the threshold for outbreak declaration in the model is five cases of invasive disease within one week. This threshold aligns with the typical threshold of 10 cases per 100,000 population used for districts with a population greater than 30,000.[75] Once this threshold is crossed, it is detected within one simulation day and the timeline for response is the same as for the other disease models.

These detection and notification timelines are optimistic, but the purpose of this analysis is to consider the impact of achieving a set of very ambitious timeliness targets.

### Scenarios

To estimate the impact of ORI response time on outbreak outcomes for each disease we ran two main scenarios and two sensitivity analyses:

- Baseline; response time and daily vaccination rate were based on the mean value from the outbreak dataset for each disease, and ORI was assumed to achieve 75% coverage in the target population (except for measles, which we assumed achieved 100% coverage);
- Faster response time; the ORI response time was varied between 15 days and the Baseline response time in steps of 15 days;
- Alternate coverage (sensitivity); the ORI coverage achieved was increased to 100% (except for measles, which was varied between 60% and 90% coverage in steps of 10%) in the target population. For meningococcal meningitis we also ran an additional sub-scenario with broadened age-eligibility and 75% coverage;
- Alternate vaccination rate (sensitivity); the daily vaccination rate was varied to produce either a one-day campaign or a one-month campaign for each disease.

Each scenario analysed comprised 1000 stochastic simulations. Table 3 describes the parameter values varied in the scenarios for each disease, and Figure 1 represents the scenario impacts on the outbreak simulations. Specific justifications (and sources) for the parameter values chosen for each disease under each modelled scenario are available in Sections 2 – 5 in ‘Additional file 1’.

**Figure 1:**
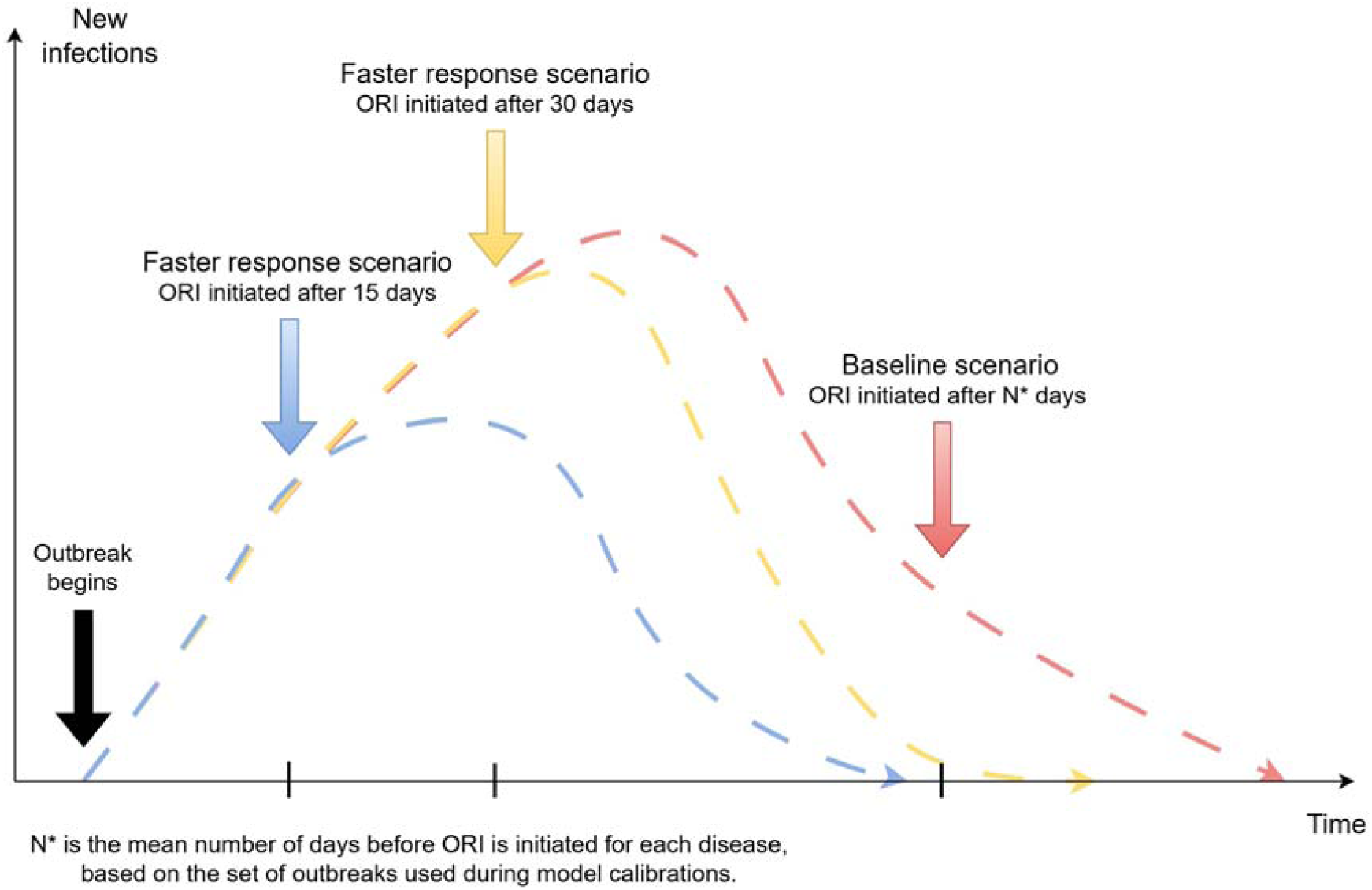
Representation of how the modelled scenarios impact the outbreak simulations. Outbreak response immunisation (ORI) implemented earlier in an outbreak is expected to reduce the total size and duration of the outbreak.

**Table 3:**
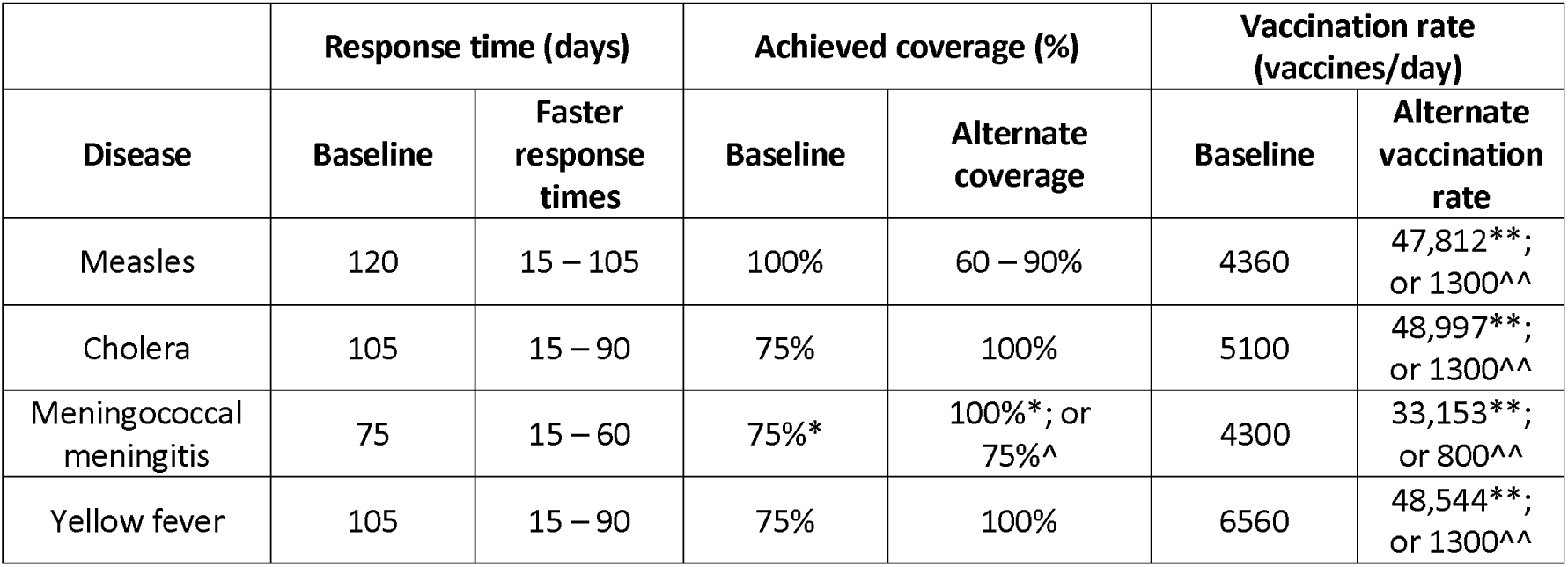
Parameter values used for each scenario analysed for measles, cholera, meningococcal meningitis, and yellow fever. * targeting ages 1 – 29 years; ^ targeting ages 1+ years; ** vaccinating the eligible population in one day; ^^ vaccinating the eligible population in one month.

### Setting archetypes

As an additional dimension of this analysis, we aimed to explore the impact of ORI response time for different ‘setting archetypes’ in which transmission was expected to be relatively higher or lower for each disease. In previous work, key parameters were identified for the measles, meningococcal meningitis, and yellow fever models, which varied by outbreak setting and influenced the expected size of outbreaks in our models.[8] For each scenario considered in this analysis, we simulated outbreaks over a range of values for these parameters (routine vaccine coverage, asymptomatic carriage prevalence, or mosquito-to-human transmission modifier), which represent outbreaks occurring in settings with different levels of risk that the outbreak may grow particularly large.

### Impact estimation and outbreak size thresholds

For each disease, setting archetype, and scenario, we recorded the distribution of cumulative cases and deaths which occurred across the outbreak simulations. The mean difference in cumulative cases and deaths between scenarios and associated uncertainty in the mean were estimated from these collections of simulations using bootstrap resampling (i.e., for each outbreak to produce estimates of cases averted with a faster or further reaching response).

We produced a distribution of outbreak sizes for each disease, setting archetype, and scenario, by defining a series of outbreak size thresholds for each disease and assigned each simulation to its appropriate threshold range. This allows us to estimate the frequency with which outbreaks exceeding certain total case thresholds occur, given a response time and achieved coverage by the ORI, based on methods adapted from previous analyses of COVID-19[76] and Ebola.[77] The impact of ORI on reducing the risk of large outbreaks was estimated by comparing the distribution of cumulative cases across simulations in the Baseline scenario against the distributions in each of the Faster response time scenarios. All impacts were disaggregated by setting archetypes, where relevant.

## Results

Across the four diseases, improvements to ORI response times reduced the proportion of simulations exceeding different outbreak size thresholds in higher risk setting archetypes (i.e., those with lower routine vaccine coverage, higher initial prevalence, or higher transmission modifiers) (Figure 2, left column). This behaviour was not observed (or the relationship was weaker) in the lower risk settings (Figure 2, right column). Additional examples of these figures for different setting archetypes are in Sections 2, 4, and 5 in ‘Additional file 1’, however those in Figure 2 were chosen as the ‘worst’ and ‘best’ archetypes for each disease (except cholera) which show the highest and lowest overall risks for large outbreaks.

**Figure 2:**
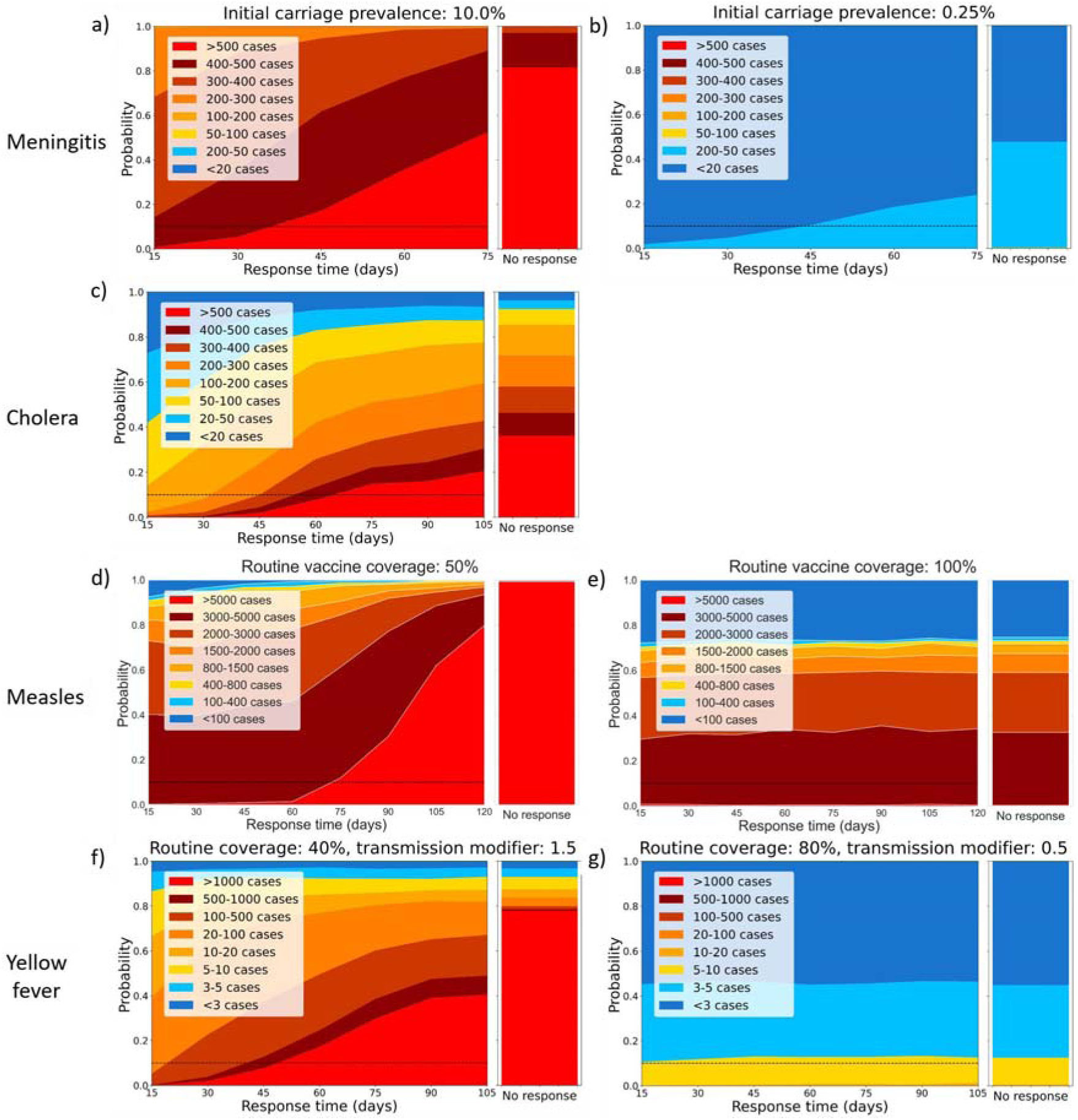
Area plots representing the proportion of outbreak simulations which exceed certain case thresholds. These proportions are plotted as a function of outbreak response immunisation (ORI) response time; disaggregated for different diseases (rows) and for worst case (left column) and best case (right column) setting archetypes considered. For each panel, the righthand stacked bar represents the distribution of outbreak sizes if no outbreak response immunisation is delivered, and the dashed line represents a 10% threshold for visualization. Panels a) and b) are meningococcal meningitis for settings with 10% and 0·25% initial meningococcal meningitis carriage prevalence, respectively. Panel c) is for cholera (only one setting archetype considered). Panels d) and e) are measles for settings with 50% and 100% routine measles vaccine coverage. Panels f) and g) are yellow fever for settings with 40% and 80% routine yellow fever vaccine coverage (BC) and with a mosquito-human transmission modifier of 1·5 and 0·5, respectively.

Each subplot in Figure 2 also has a secondary stacked bar graph next to the area graphs showing the distribution of model simulations in which outbreaks exceed the defined case thresholds. The stacked bar plot shows the same distribution for a set of model simulations in which ORI was not implemented during the outbreaks. In the high-risk setting archetypes (and the single cholera subplot), the proportion of simulations which exceeded the highest case thresholds when no ORI was used was always higher than for the slowest response time simulations. This was also observable in the figures for different setting archetypes in Sections 2, 4, and 5 in ‘Additional file 1’. In these modelled settings, even a slow ORI response was beneficial relative to no response at all.

### Meningococcal meningitis

The expected size of the meningococcal meningitis outbreaks was strongly dependent on the initial carriage prevalence (Figure S4, Figure S5 in Section 2 in ‘Additional file 1’). When the initial prevalence was low outbreaks tended to be quite small and ORI response times had minimal impact on expected outbreak size. However, when the initial prevalence was high the expected outbreak size was larger, and response time had a greater impact on the proportion of simulations which exceeded different case thresholds (Figure 2 and Figure S6 in Section 2 in ‘Additional file 1’).

While the ORI response time was more critical for absolute numbers of cases in settings with higher initial prevalence, the relative impact of a faster ORI response time was consistent across the choice of initial prevalence (Figure 3). For example, a 15-day response time was estimated to avert ∼35% of the case burden compared to the mean historical response time (75 days) between 2011 – 2022 regardless of initial prevalence. Similarly, a 30-day response time averted ∼25% of the cases, a 45-day response averted ∼15%, and a 60-day response averted ∼7% (Table S2 in Section 2 in ‘Additional file 1’).

**Figure 3:**
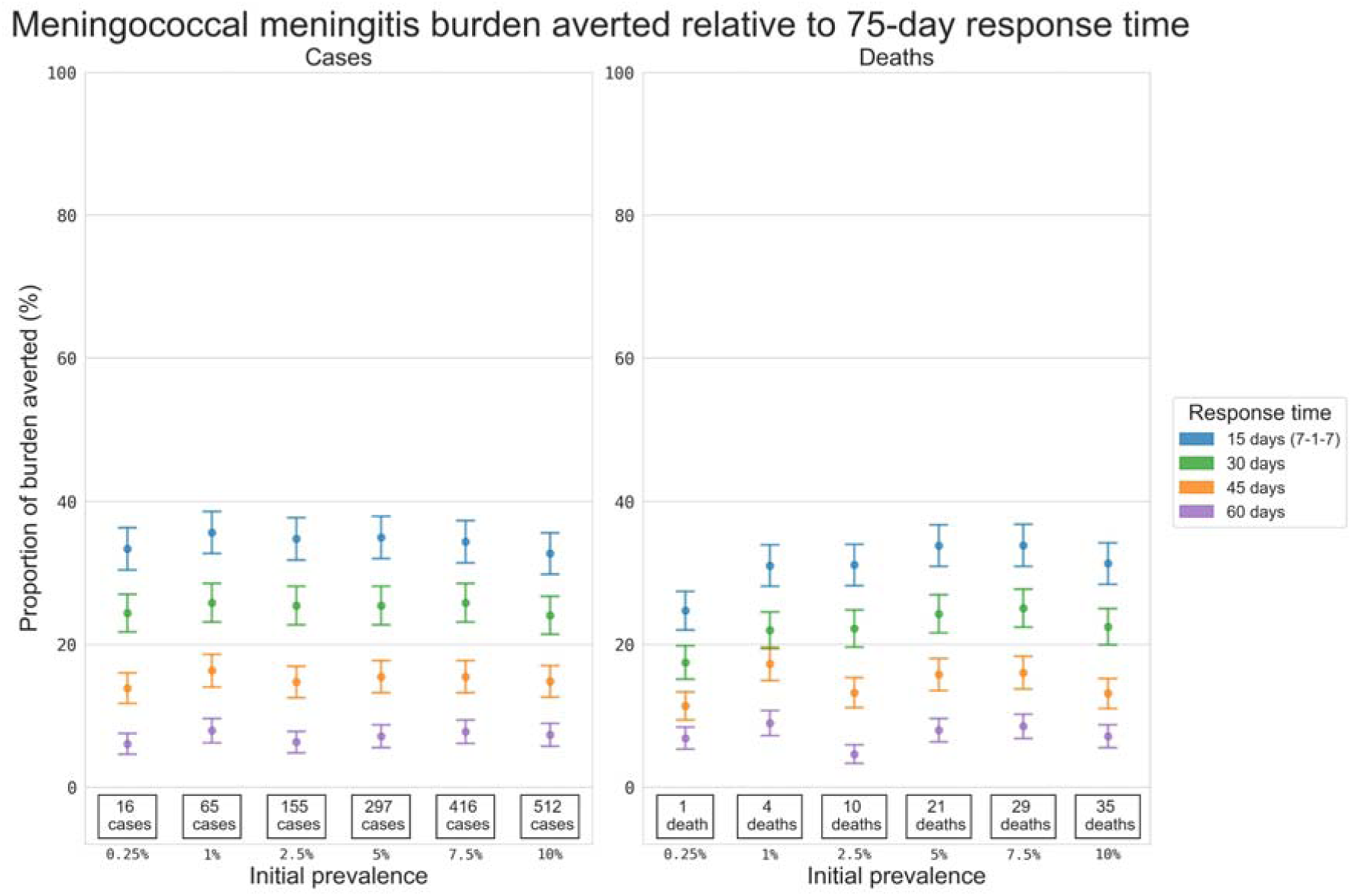
Proportional reduction in the mean cumulative cases and deaths for the simulated meningococcal meningitis outbreaks. Results include outbreaks with ORI response times of 15, 30, 45, and 60 days, relative to the set of outbreaks with a response time of 75 days, estimated using bootstrap resampling. Proportional impacts are grouped vertically by initial asymptomatic carriage prevalence, and the values stated at the bottom of each column are the mean cumulative cases or deaths produced by meningitis outbreaks which received ORI after 75 days, i.e., the value serving as the denominator for the proportional reduction estimate.

### Cholera

The expected size of the cholera outbreaks was strongly impacted by the ORI response time (Figure S9 in Section 3 in ‘Additional file 1’), and the proportion of simulations exceeding certain case thresholds also increased with ORI response time (Figure 2 and Figure S10 in Section 3 in ‘Additional file 1’). For example, slower response times noticeably increased the proportion of simulations which exceeded 400 or 500 cumulative cases. When comparing outcomes with faster ORI response times to outcomes with the mean historical response time (105 days) between 2015 – 2023, a 15-day response time was estimated to avert an additional ∼80% of the cases. Similarly, a 30, 45, 60, 75, or 90-day response averted an additional ∼70%, ∼55%, ∼40%, ∼30%, or ∼25% of the cases, respectively (Figure 4 and Table S3 in Section 3 in ‘Additional file 1’).

**Figure 4:**
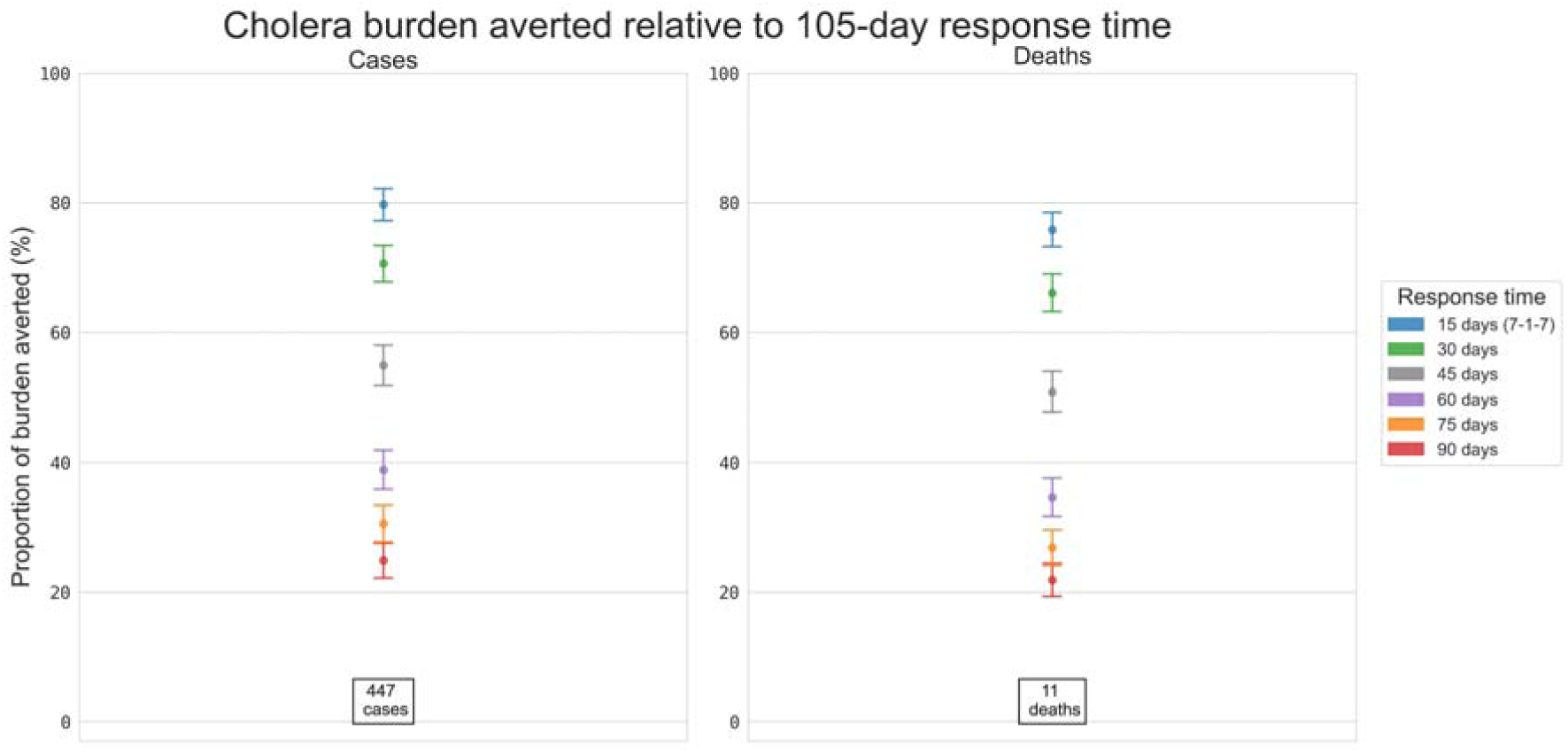
Proportional reduction in the mean cumulative cases and deaths for the simulated cholera outbreaks. Results include outbreaks with response times of 15, 30, 45, 60, 75 and 90 days, relative to the set of outbreaks with a response time of 105 days, estimated using bootstrap resampling. The values stated at the bottom of the column are the mean cumulative cases or deaths produced by cholera outbreaks which received ORI after 105 days, i.e., the value serving as the denominator for the proportional reduction estimate.

### Measles

The expected size of the measles outbreaks was strongly dependent on the routine vaccine coverage (Figure S13, Figure S14 in Section 4 in ‘Additional file 1’), and the proportion of simulations exceeding certain high case thresholds decreased with routine coverage and increased with ORI response time (Figure 2 and Figure S15 in Section 4 in ‘Additional file 1’). When the routine vaccine coverage was high (i.e., in a low-risk setting archetype), outbreaks tended to be quite small, even with longer ORI response times. However, in high-risk settings, the expected outbreak size increased and ORI response time had a greater impact on outcomes, with slower ORI response time scenarios having an increased proportion of simulations which exceeded 5000 cumulative cases.

ORI response time was more critical for absolute numbers of cases in higher risk setting archetypes, and the proportional impact of a faster ORI response time was also greater for outbreaks in these settings. Interestingly, the estimated impacts of faster response times decreased to 0% as the routine vaccine coverage approached 100% (Figure 5), which occurred because high routine vaccine coverage settings had smaller outbreaks and saw less benefit from fast ORI response times due to slower transmission. For example, in a setting with 50% routine vaccination coverage, a 15-day response time was estimated to avert ∼55% of the cases compared to the mean historical response time (120 days) between 2000 – 2022, while a 105-day response time averted ∼13%. However, in a setting with 90% routine vaccine coverage, a 15-day response time was estimated to avert ∼8% of the cases, while a 105-day response time averted no additional cases compared to a 120-day response time (Table S4 in Section 4 in ‘Additional file 1’).

**Figure 5:**
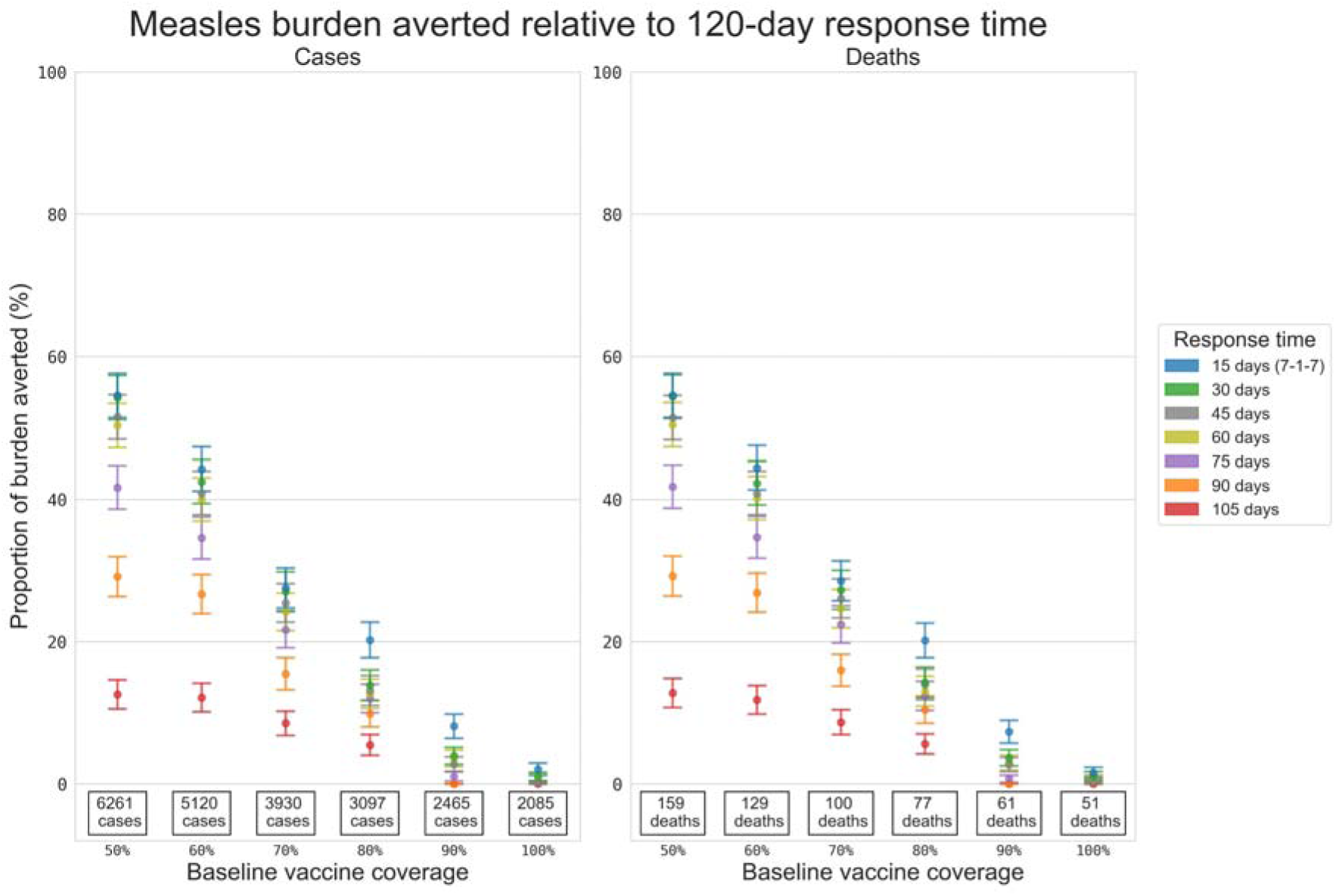
Proportional reduction in the mean cumulative cases and deaths for the simulated measles outbreaks. Results include outbreaks with response times of 15, 30, 45, 60, 75, 90 and 105 days relative to the set of outbreaks with a response time of 120 days, estimated using bootstrap resampling. Proportional impacts are grouped vertically by routine measles vaccine coverage, and the values stated at the bottom of each column are the mean cumulative cases or deaths produced by measles outbreaks which received ORI after 120 days, i.e., the value serving as the denominator for the proportional reduction estimate.

### Yellow fever

The expected size of the yellow fever outbreaks was strongly dependent on both the routine vaccine coverage (Figure S17, Figure S19 in Section 5 in ‘Additional file 1’) and the mosquito-human transmission modifier (Figure S18, Figure S20 in Section 5 in ‘Additional file 1’). The proportion of simulations exceeding certain high case thresholds decreased with routine coverage, increased with ORI response time (Figure 2 and Figure S21 in Section 5 in ‘Additional file 1’), and increased with the transmission modifier (Figure S22 in Section 5 in ‘Additional file 1’). When the routine vaccine coverage was high or the transmission modifier was low (i.e., in a low-risk setting archetype), outbreaks tended to be quite small even with longer response times. However, in a high-risk setting archetype, the expected outbreak size increased and slower response time scenarios increased the proportion of simulations which exceeded 500 or 1000 cumulative cases.

ORI response time was more critical for absolute numbers of cases in high-risk setting archetypes, and the proportional impact of a faster response time was also greater for outbreaks in these settings (Figure 6 and Figure S23 in Section 5 in ‘Additional file 1’). For example, when the transmission modifier was fixed at 1·0, in a 50% routine vaccination setting, a 15-day response time was estimated to avert ∼25% of the cases compared to the mean historical response time (105 days) between 2000 – 2022, while a 90-day response time averted no additional cases. While in an 80% coverage setting, a 15-day response time was estimated to avert ∼11% of the cases, and a 90-day response time averted no additional cases compared to a 105-day response time (Tables S5, Table S6 in Section 5 in ‘Additional file 1’).

**Figure 6:**
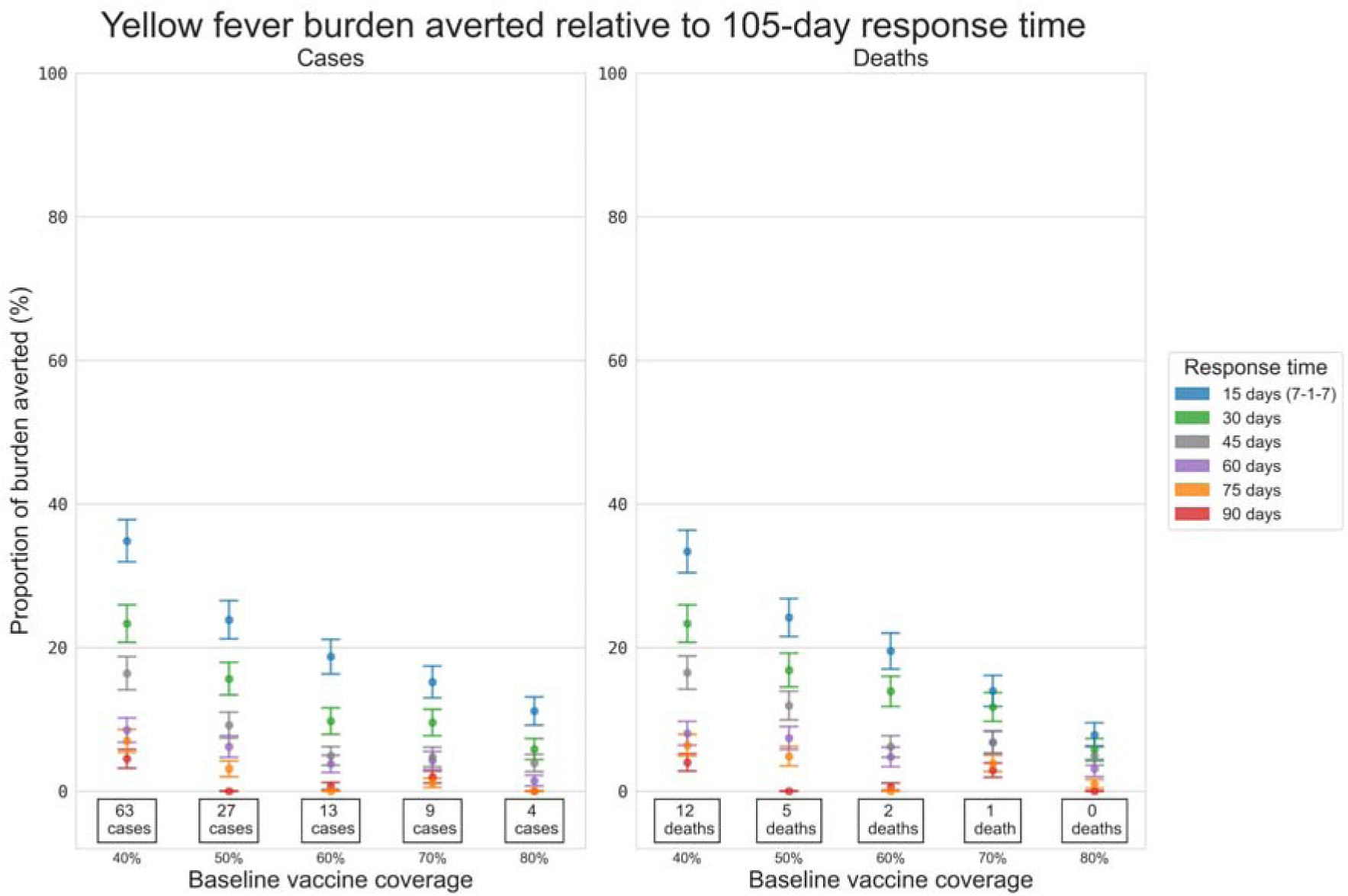
Proportional reduction in the mean cumulative cases and deaths for the simulated yellow fever outbreaks. Results include outbreaks with response times of 15, 30, 45, 60, 75, and 90 days relative to the set of outbreaks with a response time of 105 days, estimated using bootstrap resampling and assuming a transmission modifier of 1·0 for the mosquito vectors. Proportional impacts are grouped vertically by routine yellow fever vaccine coverage, and the values stated at the bottom of each column are the mean cumulative cases or deaths produced by outbreaks which received ORI after 105 days, i.e., the value serving as the denominator for the proportional reduction estimate.

### Sensitivity analysis

The results of the alternate ORI coverage sensitivity analysis can be seen in Figures S24 – 40 in Section 6 in ‘Additional file 1’. For meningococcal meningitis and cholera outbreaks, increasing achieved ORI coverage decreased the cumulative cases by up to 20 – 25%. For measles outbreaks, we found that when the achieved ORI coverage was low, outbreaks grew quite large even with rapid ORI response times, because large proportions of the population were missed by the response. The effect varied by setting archetype, and we observed less impact from gains in ORI coverage achieved if the routine coverage level was higher.

The results of the daily vaccination rate sensitivity analysis can be seen in Figures S41 – 48 in Section 6 in ‘Additional file 1’. Meningococcal meningitis, cholera, and measles outbreaks showed little sensitivity to increasing the daily vaccination rate to deliver all doses in a single day, as the rates used in the primary scenarios were already high (varying by disease).

However, yellow fever outbreaks were more sensitive to this change, with total cases decreasing by up to 40% across varying levels of response time and routine vaccine coverage, for a transmission modifier value of 1·5. For lower transmission modifier values, the decrease was smaller but still noticeable. This is likely due to the relatively longer time for the immunity to fully develop within the vaccinated agents (30 days for yellow fever[68], relative to ten days or fewer for the other diseases[48, 62, 65]), making earlier vaccine delivery relatively more impactful than for the other diseases. The impact of decreasing the vaccination rate such that the doses are delivered over the course of a month varied by disease, and within each disease varied by response time or by setting archetype. The cholera, yellow fever, and measles ORI were sensitive to the decrease, particularly for faster ORI response times and settings with lower routine vaccine coverage (for measles). Yellow fever outbreaks in the highest risk setting archetypes were up to three times larger if the ORI took one month to vaccinate the population instead of one week – which also likely results from the slower immunity development – however such a slow rollout would be atypical.

## Discussion

We used agent-based models to estimate the impact of faster ORI response times for outbreaks of measles, cholera, yellow fever, and meningococcal meningitis in synthetic LMIC populations, stratified into setting archetypes. We found that ORI responses initiated within the timeframe of the 7-1-7 framework (15 days of the outbreak beginning) would avert a larger proportion of cases and deaths than the average response times for recent historical outbreaks in LMICs.

In the models, a 15-day ORI response time reduced the expected burden of cases and deaths from meningococcal meningitis outbreaks by approximately 35%, cholera outbreaks by 80%, and from measles and yellow fever outbreaks in settings with low routine vaccine coverage by up to 55% and 35%, respectively. This is consistent with previous modelling analyses, which estimated that implementing vaccination responses faster for outbreaks of measles, Ebola, cholera, yellow fever, poliovirus, and meningococcal meningitis reduced disease burden.[8, 19–26, 28, 29] This also supports the push for frameworks like 7-1-7, because the diseases considered here are responsible for a large health burden in LMICs,[78] and the time taken to initiate ORI in LMICs is typically on the order of months.[8, 11] If ORI timeliness can be improved then delivering the same number of vaccines will have a much greater protective impact for the population at-risk, which could help accelerate progress towards targets like SDG 3.3. While these results are not granular enough and the models used not detailed enough to directly inform how ORI timeliness could be improved, ideally these impact estimates could motivate bottleneck analyses and other studies which could directly inform investment priorities.

Caution is needed when focusing on ambitious targets like 7-1-7, particularly when they are optimistic compared to current timeliness metrics for ORI in LMICs,[4, 10] because it may be viewed as unrealistic to achieve in practice. In the case of the diseases modelled here, achieving the 7-1-7 targets for the ORI component of an outbreak response reflects a more than 80% reduction in response times relative to the historical averages used in the Baseline simulations, which may be challenging to achieve in many low-resource settings.

In high-income settings, robust reporting principles, established public health surveillance systems, and strong infrastructure with a well-trained workforce facilitate rapid detection of outbreaks and rapid initiation of appropriate response measures. In contrast, in LMICs, delays to each stage of detection, notification, and response are more common because of the relatively weaker systems, infrastructure, and workforce training, among other causes. Even when these delays are known, the lack of consistent reporting during outbreaks in LMICs can make benchmarking and identifying areas for improvement a challenge.[2, 10, 12–14] It should be noted, however, that these results still indicate that even a slow ORI response is noticeably better than no response at all (Figure 2). Reactive vaccination is an effective intervention, even if delayed, but our results indicate that improving timeliness could be a clear method for maximising the protective impact achievable during an outbreak.

In low-resource settings, a more nuanced approach, with incremental targets or a graded assessment of progress towards specified targets is preferable to an ‘all-or-nothing’ goal, and is more in-line with the intent of the 7-1-7 Alliance and Resolve to Save Lives to identify and address bottlenecks rapidly in outbreak response.[1, 2] For example, consider a given year in which a country had no outbreak responses achieve 7-1-7, but 90% of its responses achieved 7-1-37 (for a total time of 45 days). If those responses were estimated to have averted a large proportion of disease burden relative to the country’s average response time from previous years, then the level of progress should be noted and a subsequent goal perhaps set towards 30 days, rather than being assessed as making no progress toward 7-1-7. Our results indicate that achieving a 15-day ORI response time could avert an additional 35 – 80% of cases and deaths, but we also examined the benefits of incrementally faster ORI response times, finding that any improvements relative to the average historical response times reduces both the expected outbreak size and negative health outcomes.

The benefits of incrementally improving ORI response times varied widely across diseases and setting archetypes, suggesting that there may be utility in having context-specific timeliness targets, and that efforts to improve ORI response times could be focused for increased impact. When outbreaks were simulated in higher risk settings (i.e., those with lower routine vaccine coverage, higher initial prevalence, or higher transmission modifiers) we consistently found a stronger relationship between ORI response time and outbreak size than in lower risk settings. By computing risk profiles for setting archetypes (as in Figure 2), context-dependent timeliness targets could be designed which minimise the risk of large outbreaks but are evidence-based and support prioritisation of limited resources for greatest benefit. For example, in a setting where the meningococcal meningitis carriage prevalence in the lead-up to an epidemic season was suspected to be high (e.g., 10% in a district), a 30-day ORI response time may be adequate to keep the risk of an outbreak larger than 500 cases below 10%. For yellow fever, in low-risk outbreak settings, ORI response time may make minimal difference to outcomes and so efforts for improvement may be better directed elsewhere. This information allows decision makers to prioritise efforts where incremental improvements have the greatest impact.

Another factor to consider when designing ambitious targets for response time is the potential for diminishing returns. While it seems clear that a shorter response time would not lead to worse outcomes during an outbreak, our results indicate that it may not always produce better outcomes. In low-risk setting archetypes the impact of improving ORI response time on both outbreak size and proportional reductions was typically quite small, but the reasons for this are clear. High routine vaccine coverage reduces both the pool of susceptible people and the rate of transmission during an outbreak of measles or yellow fever, for example. What is more interesting is that there may be diminishing returns in high-risk settings as well, depending on the disease. For measles outbreaks in settings with low routine vaccine coverage there were benefits from each incremental improvement to ORI response time from the historical average of 120 days down to 60 days, with each 15-day improvement to this time reducing total cases by 10 – 15%, however for even faster responses there was almost no additional benefit. In contrast, for meningococcal meningitis, the expected size of an outbreak was modified by the initial carriage prevalence in the population before an epidemic season, but the proportional impact of each improvement in ORI response time was largely independent of this. We found that each 15-day improvement reduced the total cases by 7 – 10% for each level of initial carriage.

One of our sensitivity analyses considered the impact of achieving varying levels of ORI coverage during an outbreak. Achieving higher coverage with ORI averted a higher proportion of cases, as would be expected, with a relatively smaller effect size observed when response times were longer. Interesting implications can be drawn from the results of the measles ORI coverage sensitivity analysis specifically, as it varied both ORI coverage and response time. Figure S27 in Section 6 in ‘Additional file 1’ shows that the expected size of outbreaks in the measles model was more strongly impacted by the level of ORI coverage achieved than by ORI response time (over the parameter ranges tested in our simulations). This indicates that for measles, it may be preferable to prioritise a comprehensive ORI campaign that reaches as many people as possible, even if it takes slightly longer to implement, as this would help close immunity gaps that allow continued transmission.

Future modelling could explore this potential trade-off between response speed and coverage in more depth and across other diseases. This is an important consideration in the context of reaching unvaccinated children, particularly those who live in remote communities. The logistical and sociological difficulties of reaching these subpopulations to try and achieve high levels of vaccine coverage in a geographic region are well known,[79] and while the assumption of achieving 100% coverage in the measles analysis may be overly optimistic, hopefully the results presented here can help to motivate additional efforts to reach these vulnerable populations.

This analysis focused on outbreaks of four vaccine preventable diseases — meningococcal meningitis, cholera, measles, and yellow fever — and found that the relationship between ORI response time and impact differs for all of them. However, there are some lessons that can be generalised across outbreaks of VPDs in LMICs. Where there are known factors which affect outbreak risk or the rate of transmission, ORI response time is typically more critical in higher risk settings. In the face of slowing and declining global vaccination rates (particularly since the COVID-19 pandemic) and the ongoing impacts of climate change, many countries may begin to face increasing outbreak risks[80, 81]. As such, developing and maintaining systems of disease surveillance to assess these risks could help to inform the importance of rapid outbreak response. Strong surveillance systems would facilitate faster and more sensitive outbreak detection, a core tenet of 7-1-7, making rapid responses more achievable. Through development of technology and investments in health systems, the timeliness for both detection and response has improved over recent decades, but there is still room for growth.[82, 83] We believe there is potential for future mathematical modelling work to meaningfully inform the benefits of enhancements to surveillance systems on the impact of outbreak responses using models such as ours.

### Limitations

This was a broad modelling analysis, using four different agent-based models in synthetic populations, and both the models and methodology are subject to limitations. Specific limitations for each disease model are described in Sections 2 – 5 in ‘Additional file 1’.

Broadly, the study methodology was limited by the nature of our calibration process, and by the assumptions which informed our scenarios. The calibration of our models was performed collectively on outbreak datasets of different sizes,[8] and while each model reproduces the range of outbreak sizes observed in the data, they are biased towards the most frequently observed outbreak sizes in LMICs. For the purposes of this prospective analysis, it means that the model simulations represented outbreaks similar to the average outbreak observed in LMICs between 2000 – 2023 for each disease, but future outbreaks may behave differently. One additional factor which may influence this is that the period between 2000 – 2023 covers the COVID-19 pandemic, which has had measurable impacts on routine vaccine coverage for measles and yellow fever,[84] and may have affected changes in human behaviour and contact networks. Our exploration of different vaccine coverage scenarios provides insight into the first impact, but it is unclear to what extent contact patterns may have changed and our models therefore cannot account for these changes in potential future outbreaks. As our models and synthetic populations were designed to focus on outbreaks in LMICs, these results are not generalisable outside of an LMIC context.

As our analysis only considered the impact of ORI programs on cases and deaths accrued over the course of a single outbreak for each set of simulations, we do not account for the mid to long-term protective impacts which may be imparted by the vaccines. It is unclear how this would change the estimated relative impacts of improving ORI response time which we presented, but it means that we are likely underestimating the overall protective impact imparted by the ORI campaigns. Particularly in the high-risk setting archetypes where the risk was driven by low routine vaccine coverage.

We aimed to understand the effects of the key parameter assumptions in our scenarios by showing simulations over a range of setting archetypes, and by performing a sensitivity analysis for the daily vaccination rate and achieved ORI coverage, but outbreaks and responses which exist outside of the considered range of the parameters may not be well represented by our results. While the models capture a lot of stochastic uncertainty associated with outbreaks, we did not capture uncertainty in some of our model parameters, such as vaccine efficacy or the probability of symptomatic disease for cholera and meningococcal meningitis, using point estimates from the literature.

Finally, our analysis only considered the impact of improvements to ORI timeliness, and no other components of outbreak response, noting ORI is only one aspect amongst the seven key early response actions within the 7-day response component of the 7-1-7 targets. We also did not disaggregate the ORI response times by detection-notification-response as defined by 7-1-7. As such we do not capture impacts from faster implementation of programs such as contact tracing, or distinguish whether timeliness improvements were made for detection, notification, or response.

## Conclusion

The results of this analysis indicate that achieving a 15-day vaccine response to outbreaks of measles, cholera, yellow fever, or meningococcal meningitis in LMICs, as per the 7-1-7 targets, could prevent a large disease burden. Furthermore, we found that there are benefits to be gained from incremental improvements to response time even when 7-1-7 is not achieved, and we support focusing on incremental improvements in timeliness irrespective of whether the 7-1-7 target is routinely achievable. We found a risk-associated variability in the impact of achieving faster responses, where response time gains were more impactful in higher risk settings than lower risk settings. Therefore, we expect that a graded approach paired with context-specific risk assessments could also facilitate resource prioritisation decisions if multiple outbreaks occur in a short timeframe.

## Declarations

### Ethics approval and consent to participate

Not applicable.

### Consent for publication

Not applicable.

### Availability of data and materials

Model code and outputs are published on Zenodo at <https://doi.org/10.5281/zenodo.15825915> and will be publicly available indefinitely. The outbreak data previously used for model calibration are available on Zenodo at <https://doi.org/10.5281/zenodo.15751150>.

## Funding

D.D. has received funding for a PhD scholarship from the National Health and Medical Research Council in Australia (Award Number: 2021/GNT2014056), which partially funded this work. The funding source was not involved in the study or decision to submit the manuscript for publication.

### Authors’ contributions

Conceptualisation, D.D., R.G.A. and N.S.; formal analysis, D.D.; data curation, D.D. and A.M.M.; investigation (modelling), D.D.; methodology, D.D., R.G.A. and N.S.; project administration, N.S.; software, D.D., A.M.M. and R.G.A.; supervision, R.G.A. and N.S.; validation, D.D., A.M.M., J.G., R.G.A. and N.S.; visualisation, D.D.; writing—original draft, D.D.; writing—review and editing, D.D., A.M.M., J.G., R.G.A. and N.S. Authors were not precluded from accessing data in the study, and they accept responsibility to submit for publication.

## Acknowledgements

The authors thank Gabrielle MacKechnie, Dr Stefanie Vaccher, Dr Todi Mengistu, and Dr Dan Hogan for their previous contributions to the outbreak dataset and models used in this analysis. The authors also thank Fenella McAndrew for her advice when creating some of the graphics.

## List of abbreviations

AIDS –: acquired immunodeficiency syndrome
COVID-: 19 – coronavirus disease 2019
IMD –: invasive meningococcal disease
LMIC –: low and middle-income country
ORI –: outbreak response immunisation
SDG –: Sustainable Development Goals
VPD –: vaccine-preventable disease

