## Additional file 1 for "Estimating the impact of decreasing vaccination response times for outbreaks of vaccine-preventable diseases in low and middle-income countries"

### 1 Overall approach across diseases

For each disease considered in this analysis (cholera, measles, meningococcal meningitis, and yellow fever) we used an agent-based model which had previously been calibrated to historical outbreak data since 2000, and is described in detail in one of the disease-specific sections below. The *Baseline* model simulations for each disease used the mean outbreak response immunisation (ORI) response time and daily vaccination rate from outbreaks in this dataset[1], as well as ORI coverage achieved among the designated target populations. We then produced alternate scenarios using matching sets of simulations with a range of faster ORI response times (to a minimum of 15 days, representing responses which meet the ‘7-1-7’ targets[2]), higher achieved ORI coverage (to a maximum of 100% coverage), or different daily vaccination rates (either achieving a one-day ORI duration or a one-month duration). By comparing the distribution of outcomes (cases, deaths, disability-adjusted life years [DALYs]) in a set of *Baseline* stochastic simulations to the distribution of outcomes for each scenario, we estimated the impact of the faster ORI response times.

For some diseases, the relationship between outbreak size and ORI response time (as well as ORI coverage achieved) are strongly mediated by setting-specific characteristics:

- *Measles:* a dimension of routine vaccine coverage at the time of the outbreak.
- *Yellow fever:* a dimension of routine vaccine coverage at the time of the outbreak, as well as a ‘transmission level’ accounting for environmental factors affecting the mosquito population and therefore changing the level of mosquito-related transmission.
- *Meningococcal meningitis:* a dimension of asymptomatic carrier prevalence at the time of the outbreak.

For these diseases, sets of simulations were run across a range of values for the relevant mediating parameter(s) and results were disaggregated by the parameter, with each set considered as representative of a ‘setting archetype.’ This was done so that the impact of a faster ORI for measles in a setting with 50% routine vaccine coverage could be considered separately from the impact in a setting with 80% routine coverage, for example.

#### 1.1 Calibration and the lattice-based approach

This section briefly describes the calibration method used for the diseases considered in this analysis, which has been more completely described in previously published work[3].

The number of outbreaks to which the models were calibrated, and limited or inconsistent data available from each setting made it computationally impractical to calibrate outbreaks individually. Additionally, stochastic effects meant that calibration to an individual outbreak (or small subset of outbreaks) may not have resulted in generalisable fitted parameters. To overcome this, we developed what we refer to as a ‘lattice’-based approach, with outbreak simulations precomputed on a grid of key parameter values, with individual outbreaks assigned to the best-fitting point in the lattice based on the nearest parameter values. The key parameters were those expected to be influential on model outcomes; for all diseases, this included response time and vaccination rate (vaccine doses delivered per capita per day). For measles, meningococcal meningitis, and yellow fever, additional covariates were included, as described above. Data were not available to inform values for these parameters in the models. Therefore, these lattice values for each outbreak in the dataset were estimated during calibration, informed by the maximum-likelihood estimate. For each disease at most one lattice dimension was estimated through calibration in this way. Additional details for these methods are in online supplemental file 1, ‘Kernel Density Estimation’, for our previous publication[3].

The range and spacing of the lattice points in each dimension were selected to minimise the discrepancy between individual outbreaks and their assigned lattice points, while also minimising the number of lattice points to improve computational tractability.

#### 1.2 Outbreak declaration and response threshold

Assumptions specific to some models may also impact the outbreak detection thresholds used for each disease. The measles model only includes children aged 0 – 5 years, and so background cases which may occur in the older population could contribute to the outbreak detection threshold. The meningitis model is initialised with an assumed background prevalence of asymptomatic disease built up over the endemic period of the year, and it is likely that background cases of invasive disease would have occurred as well. These components may lead to underestimates of detection time for measles and meningococcal meningitis, but the assumption that outbreaks are detected within one-day of the threshold being crossed for each disease is intentionally optimistic to balance this underestimation to some extent. The models are also calibrated based on these limitations, which are maintained for all scenarios.

#### 1.3 DALYs averted

Using the estimates of cases and deaths averted by ORI for each outbreak across disease models, DALYs averted can be estimated. This is done by multiplying cases averted by the years of healthy life lost due to disability (YLDs), plus years of life lost (YLLs) from deaths (for each outbreak, we use the unweighted average 2023 life expectancy of countries where outbreaks have occurred historically and compare it to the age of deaths in the model). Disability weights vary by disease, severity of presentation, and by the sequelae produced by the disease, so we estimate overall YLDs using weighted average disability weights associated with a given disease severity or sequela and its incidence, multiplied by the average duration, for the most common sequelae associated with the disease. The specific weights and sequalae are detailed for each disease in Table S1.

**Table S1**: Parameters for disability-adjusted life year (DALY) calculation per disease.

| DALY parameters |  |  |
| --- | --- | --- |
| Disability weights for meningitis infection | Acute disease: 0·133 Hearing loss: 0·074 Epilepsy: 0·263 Motor and cognitive impairment: 0·203 | Global Burden of Disease (2017) Disability Weight estimates[4]. Incidences of long-term sequelae estimated from Voss et al. (2022)[5]. |
| Disability weights for cholera infection | Mild: 0·074  Moderate: 0·188  Severe: 0·247 | Global Burden of Disease (2017) Disability Weight estimates[4]. |
| Disability weights for yellow fever infection | Severe: 0·133 |  |
| Disability weights for measles infection | Moderate: 0·051  Severe: 0·133 |  |
| Average life expectancy | Average 2023 value from countries with outbreaks in the outbreak dataset[1]. Varies by disease. | United Nations, Department of Economic and Social Affairs, Population Division[6]. Used to estimate years of life lost. |
| Discounting | 0% for DALYs |  |

#### 1.4 Analysis steps

For each disease, the analysis followed a similar sequence:

1. Calibrate model to outbreak dataset, such that it can reproduce the range of observed outbreak sizes and durations. This process is described more fully in previous work[3].
2. Run 1000 *Baseline* simulations, using the mean response time and daily vaccination rate from the outbreak dataset
   a. If the disease model has an initial condition parameter which varies across expected outbreak setting archetypes (e.g., routine vaccine coverage for measles or yellow fever) then run *Baseline* simulation across a range of values for the initial condition
3. Run 1000 simulations for each examined scenario, using simulation seeds which match the seeds from the set of *Baseline* simulations
   a. If the disease model has an initial condition parameter which varies across expected outbreak settings, then run scenario simulations across the range of values for the initial condition which match the *Baseline* simulations
4. Estimate the difference in cases/deaths/DALYs between the *Baseline* and each scenario by comparing the outcome distributions for each set of simulations

### 2 Meningococcal meningitis

#### 2.1 Background and motivation

Meningitis is an often-fatal inflammation around the brain and spinal cord, caused most frequently by infection with bacteria, however it can also be caused by viral, fungal, or parasitic infection[7]. Multiple bacteria are known to cause meningitis, but for this analysis only *Neisseria meningitidis*, which causes meningococcal meningitis is considered[8]. While people of all ages can develop meningitis, its burden is highest in very young children[8-10]. It is estimated that there are around 1·2 million cases and 135 thousand deaths due to meningitis globally per year[8, 9], with most of the burden occurring in the ‘meningitis belt’ in sub-Saharan Africa. Transmission of bacterial meningitis is highly seasonal, with the dry season (December – June) in the meningitis belt increasing both the rate of transmission and the incidence of invasive disease[11].

Most people infected with *Neisseria meningitidis* do not become symptomatic and will passively transmit to others as an asymptomatic carrier, with studies estimating that 1 – 35% of the population in the meningitis belt are colonised by the bacteria[12, 13]. The burden of disease is primarily in children and teens, with most invasive disease cases occurring in children under five years of age, and most asymptomatic cases occurring in young teens[8, 10, 13]. The case fatality rate for meningococcal disease is estimated to be around 5 – 15% in most settings[14], but around 20% of survivors will develop long-term sequelae as a result of the infection, including blindness, loss of hearing, epilepsy, or other motor/cognitive impairments[5, 14].

Due to the high cost of the vaccines, during outbreak responses vaccine delivery is highly age-targeted, and doses are typically delivered to people either 1 – 29 years or 2 – 29 years as they carry the highest burden of disease. Most historical responses have used multivalent polysaccharide vaccines which protect against invasive disease but do not impact asymptomatic infection, but since 2019 some outbreaks have been responded to with multivalent conjugate vaccines, which provide additional protection against asymptomatic carriage as well as invasive disease[15]. A previous analysis investigating the historical impact of ORI programs across 24 meningitis outbreaks occurring between 2000 – 2023 found that ORI averted 21,261 (20,268 – 22,254) cases and 1599 (1404 – 1794) deaths, representing 10 – 40% of the case and death burden for each outbreak[3]. It found that outbreaks which received faster responses tended to avert higher proportions of the expected meningitis burden, and that there is room for improvement as the mean response time was around two months.

#### 2.2 Model overview

The *Starsim* framework was used to create an agent-based model of meningococcal meningitis among humans, with states for susceptible, exposed, infected (symptomatically and asymptomatically), and recovered agents (Figure S1).

Agents in the model represent humans, who begin as susceptible, and each day have a probability of becoming infected that depends on their immunity status and is proportional to the prevalence of infection among the population. They are assigned an age (which affects infection outcomes and vaccine targeting), household contacts and community contacts. Additionally, agents can receive vaccines with characteristics matching a conjugate multivalent vaccine, which have been used in recent outbreaks in the meningitis belt [15-18]. Both susceptible and vaccinated people can become infected at a rate that is proportional to dynamic prevalence, however vaccinated people have reduced risk due to vaccine protection. Infection is primarily asymptomatic, with only a proportion of cases developing invasive meningococcal disease (IMD), and vaccination will either protect against both IMD and asymptomatic infection. Following infection, people have an incubation period before becoming infectious and can present either symptomatically or asymptomatically. Asymptomatic carriers will eventually recover and clear their carriage of the bacteria, and symptomatic cases can either die or recover. The model's progression and transmission pathways are based on structures used in other modelling studies[19, 20]. Symptomatic humans in the model are identified as a suspected case with an assumed 50% probability per day once they develop symptoms, so most cases will be found within one to two days. This identification is assumed to occur clinically through presentation to healthcare and subsequent reporting of suspected cases which present with invasive disease.

As the transmissibility and invasion rate of bacterial meningitis is highly dependent on seasonality[11], the model assumes a sinusoidal forcing function which increases the probability of transmission and the development of invasive disease for one half of the year (aligning with the ‘dry season’ in the African meningitis belt) and decreases the probability for the other half of the year:

$\beta\left\{ t \right\}=\beta(1+0\cdot6\cos\frac{2\pi t}{365})$ ,

where $\beta$ is the transmissibility parameter and $t$ is the daily step in the model. The form and parameterisation of this function are based on the methods of Karachaliou et al. (2015).[19] Additionally, as asymptomatic infection is known to be the main driver of transmission[21], and that approximately between 1% and 35% of the population in the meningitis belt is known to be carrying *Neisseria meningitidis* on average[12, 13], we assume that the background carriage rate at the start of the epidemic season is a significant driver of outbreak size (which is a theory which has been discussed before[22]), and use it to define the setting archetypes for the modelled scenarios.

The duration of vaccine immunity was set to well beyond the scope of the model period as it is assumed that no waning of immunity effects would be relevant over the outbreak period.

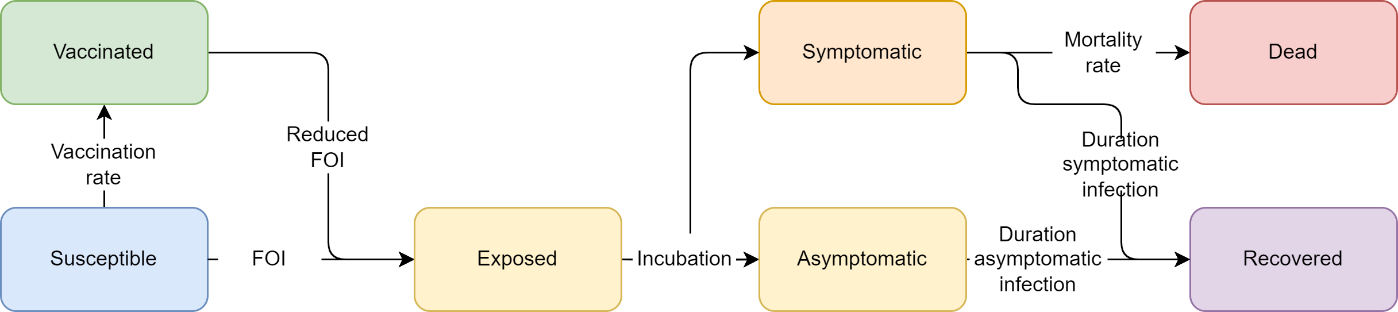

**Figure S1: Meningitis model schematic.** The Starsim framework was used to develop an agent-based model of meningococcal meningitis among humans (S-E-I-R).

#### 2.3 Model population for simulated outbreaks

The model population for each outbreak simulation represents the entire population of the specific geographic locations where outbreaks have occurred between 2000 – 2023, based on our outbreak dataset[1]. For computational reasons the model contains a maximum of 50,000 agents.

The model population was parametrised by age structure and household size distribution from the United Nations, Department of Economic and Social Affairs, Population Division[6], and age-specific household contact rates from Prem et al. (2017)[23]. The age and household size distributions in the model were generated by summing over the single year age data and household size data for each country with an outbreak included in the outbreak dataset. The age-specific household contact rates in the model were estimated using the average of the rates for each country with an outbreak included in the outbreak dataset. The age structure and household sizes are used to assign agents household contact networks, which are important for transmission in the model. The model also randomly generates ‘community’ contacts between agents, which have a much lower risk of transmission compared to household contacts[24]. These networks are randomly generated at each time step (representing a day in the model). Vaccine coverage, transmission risk and disease outcomes were modelled to vary by age, as the vaccines are highly age-targeted and both IMD and asymptomatic carriages incidences are well understood[8-10, 12, 13].

#### 2.4 Generating household networks

The household contact network was set up by explicitly modelling households, and the households size distributions were scaled to the 50,000 agents in the simulations. Each person in the model was uniquely allocated to a household. To assign ages, a single person was selected from each household as an index, whose age was randomly sampled from the age distribution used for the model. The ages of additional household members were then assigned according to age-specific household contact estimates from Prem et al. (2017)[23], by drawing the age of the remaining members from a probability distribution based on the row corresponding to the age of the index member.

#### 2.5 Diagnosis of cases, outbreak declaration and ORI

Agents within the model are assumed to be detected as suspected cases of IMD (henceforth ‘cases’) with a 50% probability per day after developing symptoms, and we assume that all cases are detected. An outbreak is declared in the model after the detection of five symptomatic agents within a week, and once this occurs the ORI will begin after N-2 days, where N is the response time for a given scenario. This threshold aligns with the typical threshold of 10 cases per 100,000 population used for districts with a population greater than 30,000[25]. Once the ORI begins, vaccines matching the characteristics of multivalent conjugate vaccines will be targeted to the defined age group for the response, assumed to be 1 – 29 years.

#### 2.6 Calibration

Calibration involves estimating the transmissibility of meningitis in the model to produce outbreaks of a sufficient size, as well as the probability of death given symptomatic disease to capture the observed case fatality rate. Given well known variations in incidence of both invasive disease and asymptomatic carriage by age, the age-based susceptibility of humans to infection and developing symptoms was adjusted in the model to reproduce the typical age distributions observed in the literature[8-10, 12, 13], seen in Figure S2. All other relevant model parameters were constrained by estimates from the literature or the outbreak dataset. The strict seasonality of outbreaks was reproducible using a sinusoidal forcing function, using the same methods as work done by Karachaliou et al. (2015)[19].

Following this initial calibration process, for a given response time and vaccination rate, outbreak simulations could be run to produce a range of stochastic outcomes that could be compared to observed outbreaks within the outbreak dataset[1]. However, these simulations did not always align with the data, with differences potentially explained by climate, pre-existing immunity, and other factors that influence heterogeneity in transmission across settings. To account for this, an additional calibration variable was introduced to specify the rate of asymptomatic carriage in the model population before the increased seasonal transmission starts an outbreak. As transmission is known to be driven by asymptomatic carriers, and the prevalence is known to be highly variable during both endemic and epidemic periods, it is used as a free parameter to drive early epidemic growth rates in the model and produce the wide range of outbreak sizes observed in the well-defined epidemic season.

The range of initial prevalence values selected for the setting archetypes to reflect the range of prevalences observed during endemic periods was: 0·25%, 1%, 2·5%, 5%, 7·5%, and 10%[22]. Figure S3 demonstrates how the initial carriage prevalence drives the initial growth, time to peak, and final size of the simulated outbreaks, while still being constrained to a 4–6-month period of high IMD burden. These epicurves were also calibrated to align with those published by WHO regional office for Africa and Inter country Support Team - West Africa in the Meningitis Weekly Bulletin reports[26-30]. Simulations initialised with an asymptomatic prevalence of 0·25% maintain a low level of asymptomatic disease and only produce a small increase in IMD cases due to the increased invasion rate during the epidemic season. However, simulations with higher initial prevalences tend to grow rapidly and produce noticeable peaks in both symptomatic and asymptomatic infections.

As the simulations are initialised with a non-zero carriage prevalence, we are in effect assuming that there was some level of endemic transmission occurring prior to when the model begins, which produced the assumed prevalence by the start of the epidemic season. This assumption means that we are ignoring the impact of population immunity which may have been developed prior to the epidemic season, and not accounting IMD cases accrued during the endemic period. These are significant simplifying assumptions, but as the rate of invasive disease outside of the epidemic season is known to be low and the model is calibrated against cases which occurred during a given epidemic season, we do not expect it to noticeably impact the calibration. The cumulative number of IMD cases and deaths produced across the range of initial prevalences allow us to reproduce the observed burdens in our outbreak data, and the estimates of increased asymptomatic prevalence during an outbreak align with estimates from the literature[12, 22].

To produce the *Baseline* simulations the model was run with a response time of 75 days and a daily vaccination rate of 4300 doses, reflecting the mean values from the outbreak dataset[1], across a range of initial carriage prevalence values (0·25%, 1%, 2·5%, 5%, 7·5%, or 10%). The daily vaccination rate results in the ORI campaign running for five days when 75% coverage is achieved in the at-risk population, and seven days when 100% coverage is achieved, which is informed by our outbreak dataset and in line with observed estimates[31]. Stochastic outbreak simulations were produced by running the model 1000 times for each prospective outbreak.

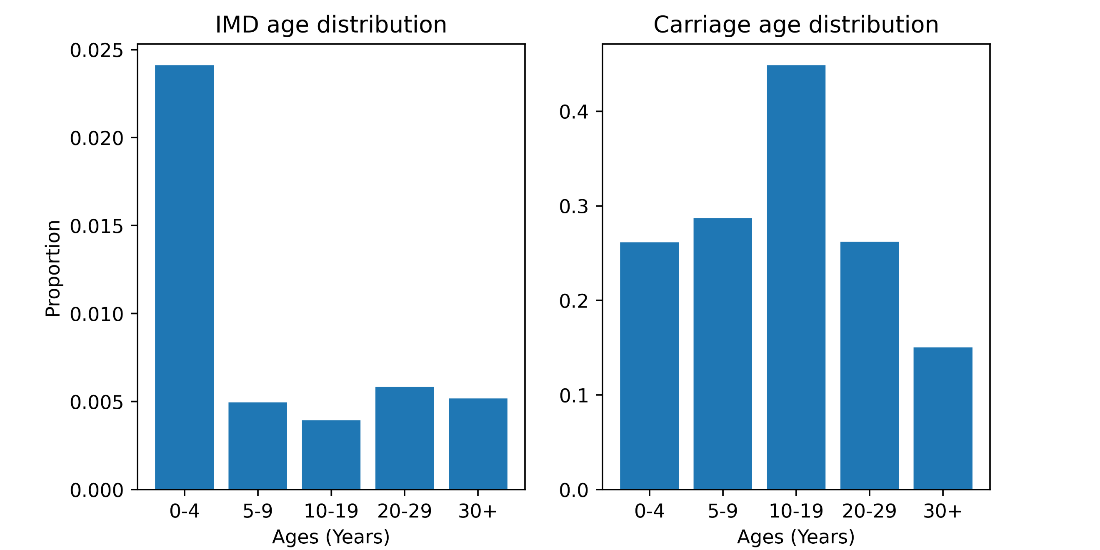

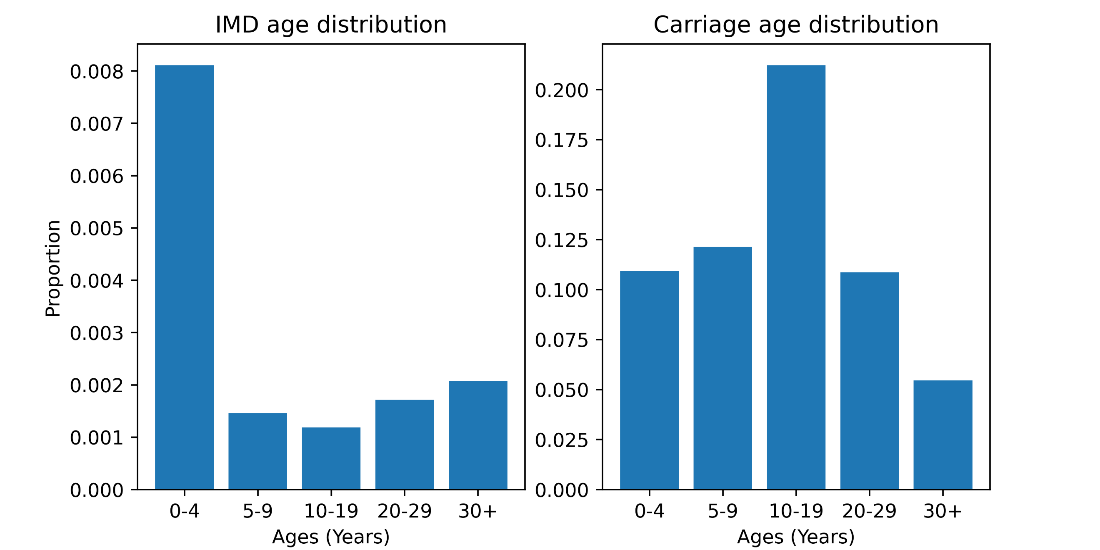

**Figure S2**: Examples of invasive meningococcal disease (IMD) case and asymptomatic infection distributions from calibrated model simulations with a vaccine response. IMD burden is highly focused in children less than one or two years of age (depending on how the target population for the response is defined), and people over 29 years of age because they typically do not receive vaccines as a part of the response. Rates of carriage in the model peak in older children and teenagers, which reflects what is observed in carriage studies from Africa.

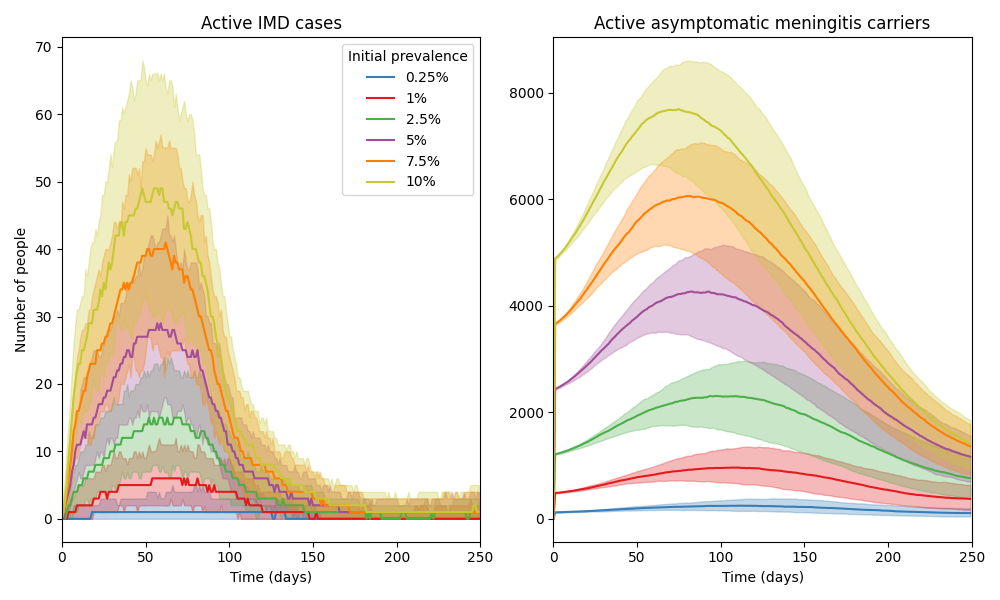

**Figure S3**: An example set of model time series (using a population of 50,000 agents) after calibration, comparing the size and timing of outbreaks produced by different initial carriage prevalences. The model is able to produce a wide range of outbreak sizes, all constrained to the epidemic season (assumed to be the first six months of model simulation), depending on the initial carriage prevalence. The initial prevalence drives the initial growth rate, time to peak, and final size of the simulated outbreaks. The colours in the figures represent the initial carriage prevalence used for the simulations.

#### 2.7 Scenarios

To explore the impact of response time and achieved ORI coverage on outbreak outcomes, we explored two scenarios and two sensitivity analyses:

- *Baseline*; response time of 75 days; daily vaccination rate of 4300 doses; achieved ORI coverage of 75% in ages 1 – 29 years,
- *Faster response time*; response time of 15, 30, 45, or 60 days; daily vaccination rate of 4300 doses; achieved ORI coverage of 75% in ages 1 – 29 years,
- *Alternate coverage* (sensitivity analysis); response time of 75 days; daily vaccination rate of 4300 doses; achieved ORI coverage of 100% in ages 1 – 29 years, or achieved coverage of 75% in ages 1+ years,
- *Alternate vaccination rate* (sensitivity analysis); response time of 75 days; daily vaccination rate of 33,153 doses, or daily vaccination rate of 800 doses; achieved ORI coverage of 75% in ages 1 – 29 years.

Each set of scenario simulations were run across the range of initial carriage prevalence values: 0·25%, 1%, 2·5%, 5%, 7·5%, or 10%. For a given initial prevalence value, the simulations used for each scenario are identical except for the scenario parameter value varied (ORI response time, achieved ORI coverage, or daily vaccination rate). The *Baseline* scenario assumes 75% at-risk population coverage was achieved by the ORI program, which is in line with other published modelling studies[31], but may be an underestimate. The effect of this assumption was investigated in our sensitivity analyses. The choices of alternate vaccination rates for the sensitivity analysis are based on the number of doses required to vaccinate the entire eligible model population in either one day or one month.

#### 2.8 Results

##### 2.8.1 Impact of response time

In the Results section of our manuscript, we show area plots representing the proportion of meningococcal meningitis outbreak simulations which exceed certain case thresholds as a function of ORI response time for settings with 10% and 0·25% initial meningococcal meningitis carriage prevalence in Figure 2. These plots also appear in Figure S6, alongside area plots for additional setting archetypes. The manuscript’s Results section also describes the proportional reduction in the mean cumulative cases and deaths for the set of meningococcal meningitis outbreaks with ORI response times of 15, 30, 45, and 60 days relative to the set of outbreaks with a response time of 75 days in Figure 3, the values for which are in Table S2. In these supplementary results we show the distribution of cumulative meningitis cases produced by the outbreak simulations (Figure S4), and a heatmap of the mean cumulative cases from the outbreak simulations for varying setting archetypes and ORI response times (Figure S5).

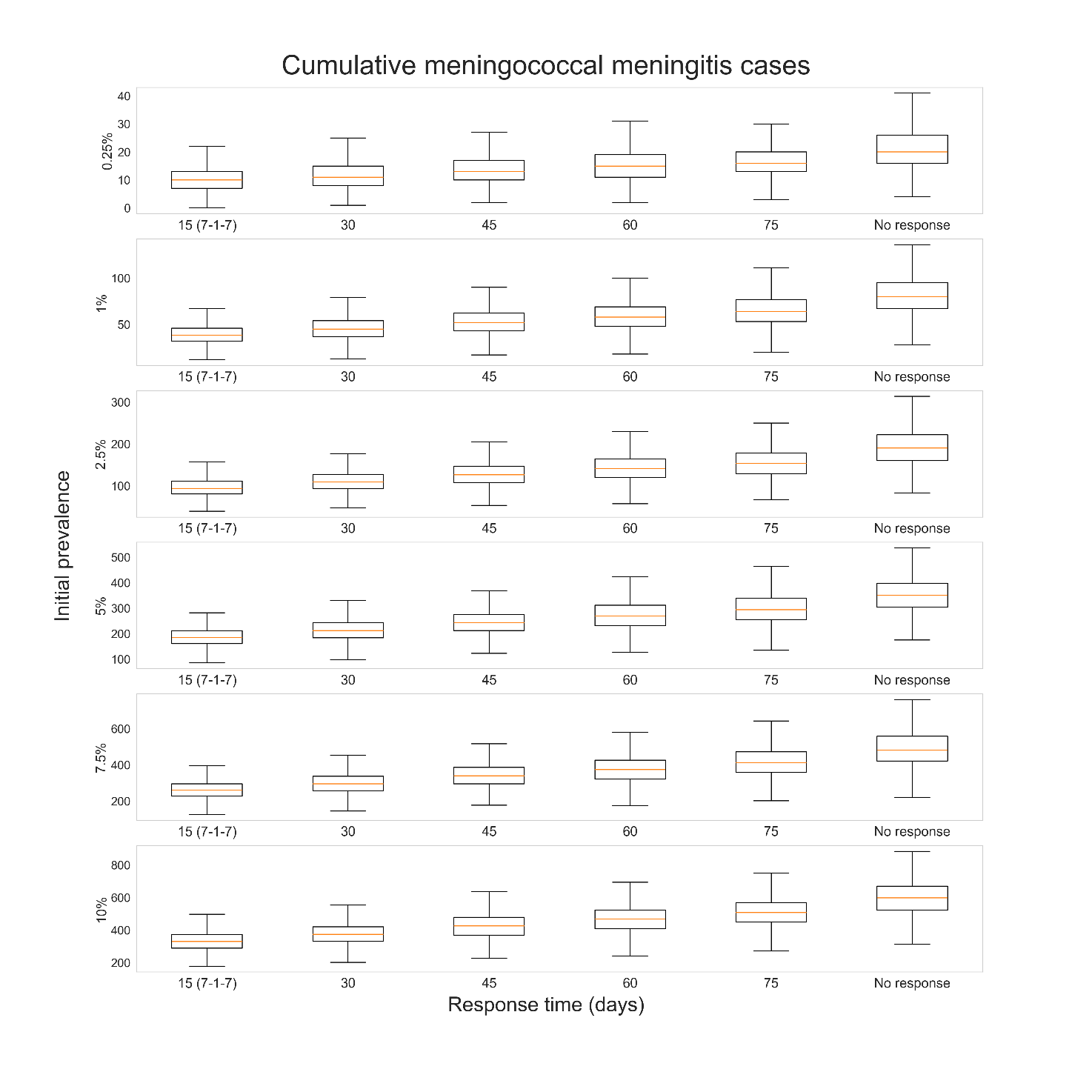

**Figure S4**: Distribution of cumulative cases produced by 1000 meningococcal meningitis outbreak simulations, given different outbreak response immunisation (ORI) response times (15, 30, 45, 60, 75 days, or no response) and initial prevalences of asymptomatic carriage in the model population before an epidemic season (0·25%, 1%, 2·5%, 5%, 7·5%, or 10%). For each initial prevalence, outbreaks without ORI produced the most cumulative cases, and when ORI occurred outbreaks which received faster responses tended to produce fewer cumulative cases.

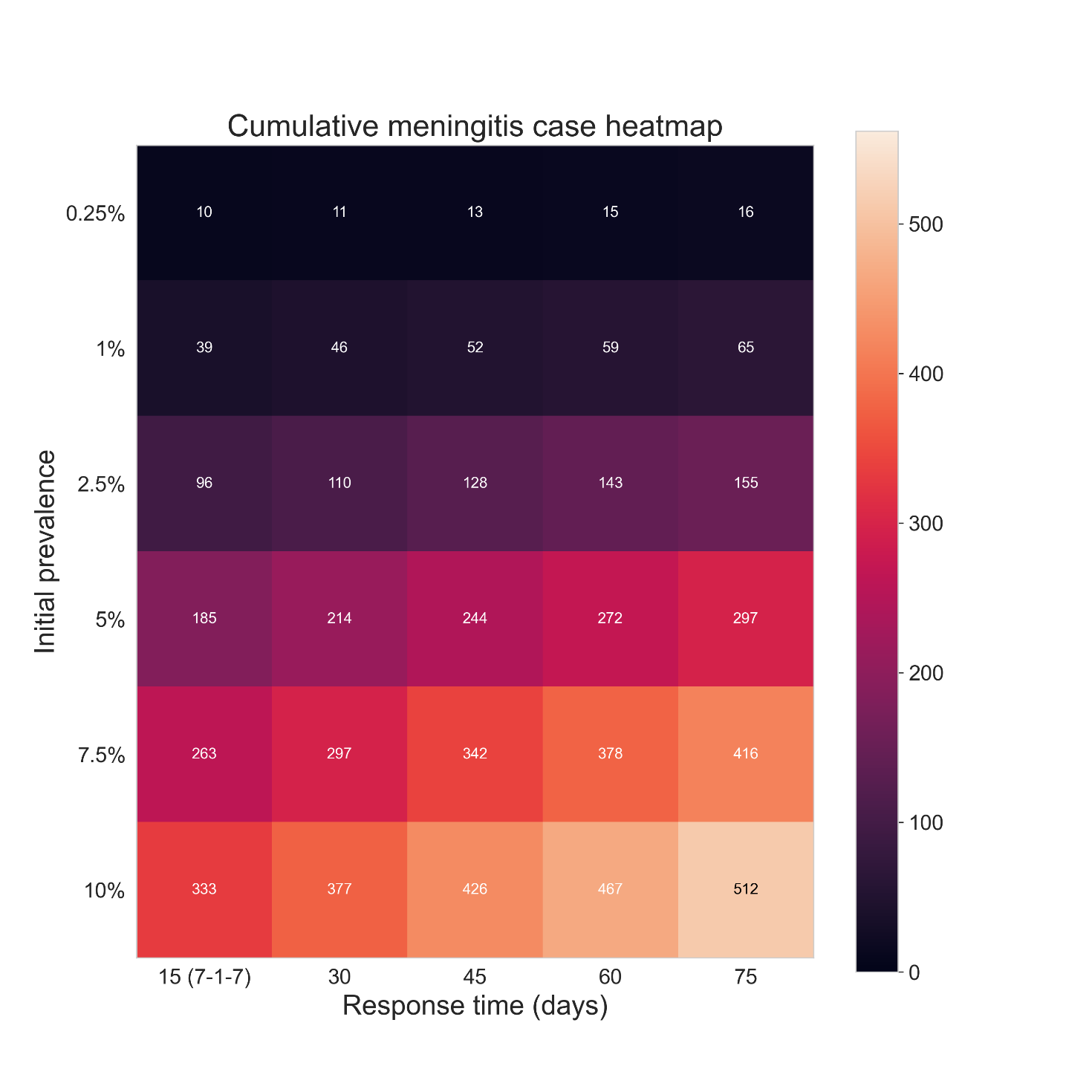

**Figure S5**: Heatmap of the mean cumulative cases produced by 1000 meningococcal meningitis outbreak simulations, given different ORI response times (15, 30, 45, 60, or 75 days) and initial prevalences of asymptomatic carriage in the model population before an epidemic season (0·25%, 1%, 2·5%, 5%, 7·5%, or 10%). For each initial prevalence outbreaks which received faster responses of ORI tended to produce fewer cumulative cases.

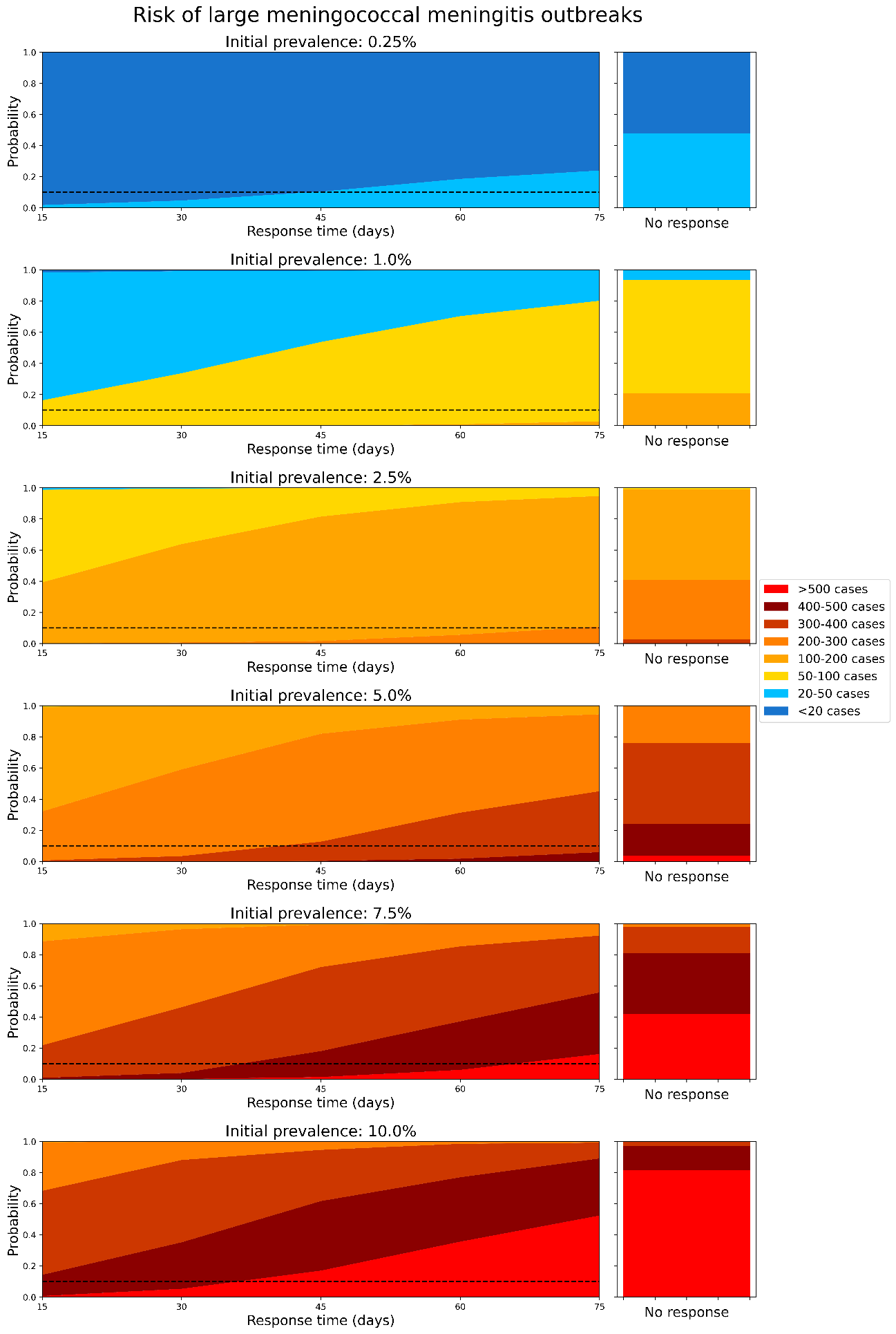

**Figure S6**: Area plots representing how the proportion of meningococcal meningitis outbreak simulations which exceed certain case thresholds changes as a function of ORI response time, assuming an initial carriage prevalence of: a)0·25%, b)1·0%, c)2·5%, d)5·0%, e)7·5%, and f)10·0% before the epidemic season begins. The dashed line represents a 10% threshold. The righthand stacked bar represents the distribution of outbreak sizes if no ORI is delivered, for reference.

**Table S2**: Proportional reduction in meningitis cases, deaths, and DALYs relative to 75-day response, for each choice of initial carriage prevalence.

| Response time (days) | Initial prevalence | Cases | Deaths | DALYs |
| --- | --- | --- | --- | --- |
| 60 | 0·25% | 0.061 (0.046, 0.075) | 0.069 (0.053, 0.084) | 0.07 (0.054, 0.085) |
| 45 |  | 0.139 (0.117, 0.16) | 0.114 (0.094, 0.133) | 0.123 (0.102, 0.143) |
| 30 |  | 0.244 (0.217, 0.27) | 0.175 (0.151, 0.198) | 0.199 (0.174, 0.223) |
| 15 |  | 0.334 (0.304, 0.363) | 0.247 (0.22, 0.274) | 0.286 (0.258, 0.314) |
| 60 | 1% | 0.063 (0.048, 0.078) | 0.046 (0.033, 0.059) | 0.053 (0.039, 0.067) |
| 45 |  | 0.147 (0.125, 0.169) | 0.132 (0.111, 0.153) | 0.162 (0.139, 0.185) |
| 30 |  | 0.254 (0.227, 0.281) | 0.222 (0.196, 0.248) | 0.262 (0.234, 0.289) |
| 15 |  | 0.348 (0.318, 0.377) | 0.311 (0.282, 0.34) | 0.364 (0.334, 0.394) |
| 60 | 2·5% | 0.079 (0.062, 0.096) | 0.09 (0.072, 0.107) | 0.095 (0.077, 0.113) |
| 45 |  | 0.163 (0.14, 0.186) | 0.173 (0.149, 0.196) | 0.199 (0.174, 0.223) |
| 30 |  | 0.258 (0.231, 0.285) | 0.22 (0.194, 0.245) | 0.247 (0.22, 0.274) |
| 15 |  | 0.357 (0.327, 0.386) | 0.31 (0.281, 0.339) | 0.329 (0.3, 0.358) |
| 60 | 5% | 0.071 (0.055, 0.087) | 0.08 (0.063, 0.096) | 0.095 (0.077, 0.113) |
| 45 |  | 0.155 (0.132, 0.177) | 0.158 (0.135, 0.18) | 0.199 (0.174, 0.223) |
| 30 |  | 0.254 (0.227, 0.281) | 0.243 (0.216, 0.269) | 0.288 (0.26, 0.316) |
| 15 |  | 0.35 (0.32, 0.379) | 0.338 (0.309, 0.367) | 0.402 (0.372, 0.433) |
| 60 | 7·5% | 0.078 (0.061, 0.094) | 0.085 (0.068, 0.102) | 0.1 (0.081, 0.118) |
| 45 |  | 0.155 (0.132, 0.177) | 0.16 (0.137, 0.183) | 0.191 (0.166, 0.215) |
| 30 |  | 0.258 (0.231, 0.285) | 0.251 (0.224, 0.277) | 0.305 (0.276, 0.333) |
| 15 |  | 0.344 (0.314, 0.373) | 0.339 (0.309, 0.368) | 0.395 (0.365, 0.425) |
| 60 | 10% | 0.073 (0.057, 0.089) | 0.071 (0.055, 0.087) | 0.078 (0.061, 0.094) |
| 45 |  | 0.148 (0.126, 0.17) | 0.131 (0.11, 0.152) | 0.161 (0.138, 0.184) |
| 30 |  | 0.241 (0.214, 0.267) | 0.225 (0.199, 0.25) | 0.274 (0.247, 0.302) |
| 15 |  | 0.327 (0.298, 0.356) | 0.313 (0.284, 0.342) | 0.373 (0.343, 0.403) |

#### 2.9 Limitations

- Actual transmission might be heterogeneous and could be a factor that limits outbreak size: there are multiple sources of heterogeneity that the model cannot capture, across the populations and transmission networks. This includes
  - Highly connected contact networks (e.g., workplaces, schools, social groups) which could lead to additional transmission networks outside of households that the randomly generated community networks do not capture well. This may lead to the model underestimating the transmissibility of meningitis, particularly the lack of school and social networks which would increase the connectivity of the high carriage younger age groups and likely increase transmission.
  - Climatic heterogeneities which impact the timing, duration, and effect of the epidemic season in each outbreak. The model uses a simple period forcing function with fixed amplitude across outbreaks, which cannot capture these potential effects.
- High uncertainty around conjugate vaccine efficacy against carriage: only a small number of studies have investigated the efficacy of conjugate vaccines against carriage of *Neisseria meningitidis*. The source for the efficacy value used in the model aligns with the value used by others in the Vaccine Impact Modeling Consortium[19], but if it is an inaccurate assumption then this would likely impact the results.
- Underestimating the long-term benefits of vaccination: our estimates of averted outcomes are explicitly focused on cases and deaths which could have occurred during a given outbreak in the absence of a vaccine response, however the long-term immunity provided to significant proportions of the population has the potential to avert future disease burden as well. For outbreaks which received large vaccine responses, the immunity gained would be equivalent to a preventative campaign. As such, our impact estimates are likely a considerable underestimate compared to impacts accrued over a longer timeframe.
- Reporting of meningococcal cases: the model is calibrated to the cumulative suspected cases reported for a set of historical outbreaks:
  - This may be inflated due to some suspected cases that are not meningococcal disease. If so, then the model may overestimate the transmissibility of meningitis which would produce overestimates of ORI impact.
  - On the other hand, reported cases are an underestimate of the true number of cases due to the limited availability of confirmatory testing. The model’s assumption that all cases of IMD are detected with one to two days is likely optimistic in this context. If so, then the model may underestimate the transmissibility of meningitis which would produce underestimates of ORI impact.
- Uncertainty around number of asymptomatic infections: there is high uncertainty around the proportion of infections which are asymptomatic as there is little data to inform this. The parameters in the model are constrained to the observed cases and deaths, and the carriage prevalence prior to and during outbreaks in the model align with ranges described in the literature, but these factors appear to be highly variable across outbreaks. It is known that the rate of invasive disease varies over time, but the degree of variation is unknown and it is unclear how to accurately parameterise it. As it is unclear whether the asymptomatic infection rate (and how it varies over time) is an under- or overestimate, it is unclear what impact this has on the results.
- Uncertainty around timing of infections over the year: the epidemic season in the meningitis belt is well-defined, and where possible the cumulative cases and deaths which were used for calibration across the set of outbreaks reflect the burden from the epidemic season. However, in some cases these data were not disaggregated by whether they occurred during the epidemic or endemic period and so the calibrations for some outbreaks may be overestimates. However, due to the typically low incidence of IMD during the wet season this impact is not expected to be high.
- Uncertainty around the shapes of the epidemic curves for the meningitis outbreaks which were included in the calibration, due to a lack of available data. While the epicurves produced by the model were calibrated to align with the epicurves published by WHO regional office for Africa and Inter country Support Team - West Africa in the Meningitis Weekly Bulletin reports, there were typically insufficient data to do this for all of the outbreaks included in the calibration.
- Uncertainty around the expected coverage achieved by ORI during an outbreak: It is unclear what level of population coverage is typically achieved by ORI campaigns, and it is likely that it would vary widely based on the outbreak context. Our assumption of 75% achieved coverage would be optimistic if outbreaks occur in remote or otherwise hard to reach populations, and would be pessimistic if the population at-risk has a strong history of vaccine uptake. We attempt to understand the impact of this assumption in our *Alternate coverage* sensitivity analysis.

### 3 Cholera

#### 3.1 Background and motivation

Cholera is a waterborne bacterial disease caused by the bacteria *Vibrio cholerae*, which can cause severe watery diarrhea and kill its host within hours if not treated[32, 33]. Infection occurs after the consumption of food or water contaminated by the bacteria, and rapid transmission can occur in populations without sources of safe drinking water or poor hygiene and sanitation conditions. Infectious people will shed bacteria, leading to further contamination of food and water, and freshly shed bacteria (within 5 – 18 hours) are highly infectious[34], which can cause increased risk of transmission within households. A significant proportion of cholera infection is asymptomatic [35, 36], so people can spread the infection without knowing and this can make detection and accurate estimation of disease burden difficult. There are around 3 million cases and 100 thousand deaths due to cholera globally per year [37], with most of the burden occurring in sub-Saharan Africa.

Outbreaks of cholera are responded to with vaccines, when possible, but supplies of the vaccine are limited and differentiation of cholera from other causes of watery diarrhea can be challenging. Provision of fresh water and improved sanitation conditions are also effective tools for preventing cholera transmission, as they reduce the risk of exposure to the bacteria[38]. A previous analysis investigating the historical impact of ORI programs across 40 cholera outbreaks occurring between 2000 – 2023 found that ORI averted 283K (273K – 292K) cases and 5215 (4879 – 5551) deaths, representing 10 – 60% of the case and death burden for each cholera outbreak [3]. It found that outbreaks which received faster responses tended to avert higher proportions of the expected cholera burden, and that there is room for improvement as the mean response time was around three months.

#### 3.2 Model overview

The *Starsim* framework was used to create an agent-based model of cholera among humans[39], with states for susceptible, exposed, infected (symptomatically and asymptomatically), and recovered agents (Figure S7). The model also has a dynamic parameter for risk of environmental transmission to humans, which is calculated at each time step based on the relative concentration of the bacteria *Vibrio cholerae* in communal water sources (assuming that infectious humans shed the bacteria and increase the concentration).

Agents in the model represent humans, who begin as susceptible, and each day have a probability of becoming infected that depends on their immunity status and is proportional to the prevalence of infection among the population and the concentration of cholera bacteria in the environment. Following infection, humans enter a latent infection ‘exposed’ state, before becoming infectious to others; this infectious state can be either symptomatic or asymptomatic. Humans in either infectious state can recover and develop immunity to further infection during the modelled outbreak, and humans with symptomatic infection can die based on a disease-specific mortality rate. People can also be vaccinated, which will provide a level of immunity against future infection. It is important to note that as the model does not include routine vaccination, and assumes no pre-existing immunity from prior infection. As such, the model population is cholera-naïve and most representative of contexts in which cholera has never been present or has been previously eliminated.

Transmission in the model can occur through two dynamic mechanisms. First, infected humans will shed bacteria during their infectious period, leading to contamination of food and water, increasing the prevalence of *Vibrio cholerae* in the environment and the risk of environmental transmission to all susceptible humans in the model. Second, people in the model are assigned a network of contacts (household and community; details in model population section), and there is a risk of human-human transmission between infected and susceptible contacts, which captures the increased infectiousness of freshly shed bacteria[34]. The prevalence of bacteria in the environment varies over time, increasing as infected humans shed fresh bacteria and decreasing as bacteria in the environment die. This prevalence is used to estimate a dynamic concentration of *Vibrio cholerae* in the environment via a dose-response relationship (parameterised as in Mukandevire et al. [2011][40]).

The model assumes an underlying level of Water, Sanitation, and Hygiene (WASH) interventions are present, which reduce the transmission risk from both human-human and environment-human transmission by improving hygiene and access to clean water, respectively. After an outbreak is detected and declared, the effectiveness of these WASH interventions is assumed to increase as people act to avoid infection.

The duration of vaccine immunity was set to well beyond the scope of the model period as it is assumed that no waning of immunity effects would be relevant over the outbreak period.

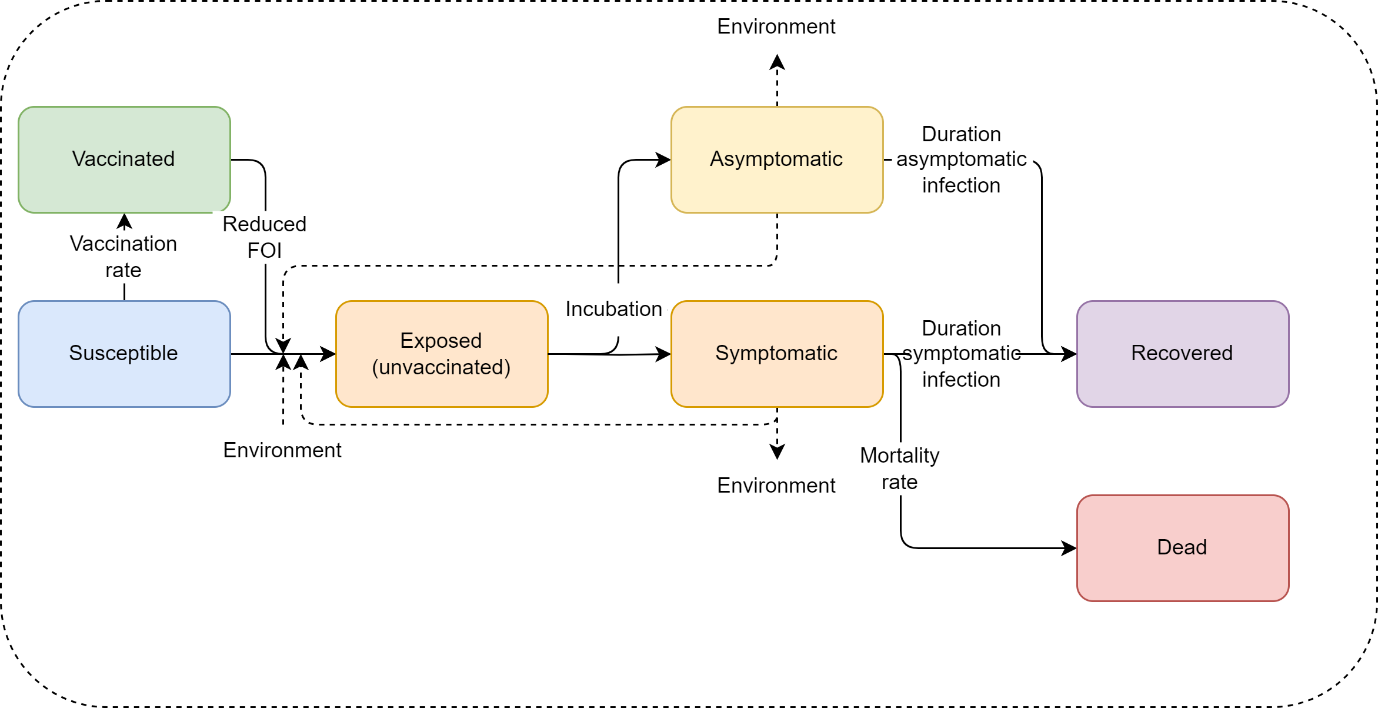

**Figure S7**: Cholera model schematic. The Starsim framework was used to develop an agent-based model of cholera among humans (S-E-I-R), which was paired with a dynamic prevalence of Vibrio cholerae in the environment to parametrise risk of environmental transmission.

#### 3.3 Model population for simulated outbreaks

The model population for each outbreak simulation represents the entire population of the specific geographic locations where outbreaks have occurred between 2000 – 2023, based on the outbreak dataset[1]. For computational reasons the model contains a maximum of 50,000 agents.

The model population was parametrised by age structure and household size distribution from the United Nations, Department of Economic and Social Affairs, Population Division[6], and age-specific household contact rates from Prem et al. (2017)[23]. The age distributions in the model were generated by summing over the single year age data for each country with an outbreak included in the outbreak dataset. The age-specific household contact rates in the model were estimated using the average of the rates for each country with an outbreak included in the outbreak dataset. The age structure and household sizes are used to assign agents household contact networks, which are important for human-human transmission in the model. The model also randomly generates ‘community’ contacts between agents, which have a much lower risk of transmission compared to household contacts[41], to capture potential transmission via freshly shed bacteria outside of an agent’s household. These community contacts are randomly generated at each time step (representing a day in the model). Vaccine coverage, transmission risk, and disease outcomes were not modelled to vary by age given limited data from the outbreak settings.

#### 3.4 Generating household networks

The household contact network was set up by explicitly modelling households, and the households size distributions were scaled to the 50,000 agents in the simulations. Each person in the model was uniquely allocated to a household. To assign ages, a single person was selected from each household as an index, whose age was randomly sampled from the age distribution used for the model. The ages of additional household members were then assigned according to age-specific household contact estimates from Prem et al.(2017)[23], by drawing the age of the remaining members from a probability distribution based on the row corresponding to the age of the index member.

#### 3.5 Diagnosis of cases, outbreak declaration and ORI

Symptomatically infected agents in the model have a daily probability (5%) of being detected as suspected cases of cholera (henceforth ‘cases’), assuming presentation at a health care facility or a diagnostic test being taken; the model does not distinguish between suspected and confirmed cases. An outbreak is declared in the model after the detection of a single symptomatic agent, and once this occurs the ORI will begin after N-2 days, where N is the response time for a given scenario. Once the ORI begins, vaccines matching the characteristics of oral cholera vaccines will be targeted to the defined age group for the response, assumed to be 1+ years. The outbreak is considered to have ended after 21 days with no new cases detected during a simulation.

#### 3.6 Calibration

Calibration involves estimating the transmissibility of cholera in the model (human-human transmission risk per contact and proportionality constant for environment-human transmission), as well as the probability of symptomatic cholera and probability of death given symptoms. This was done by fitting to outbreaks in a separate dataset, maintained by Johns Hopkins University (with summary characteristics presented by Zheng et al. (2022)[42]), which includes disaggregated time series data for 1000 cholera outbreaks from 2010 – 2020. Many of these outbreaks were not in the WHO databases as they are informed by confidential surveillance reports or were likely too small to have been reported, and most of which did not have a vaccine response so could not be included in the main analysis. These data included: outbreak location, duration, threshold for declaration, total suspected cases, attack rate, total deaths, case fatality rate, reporting frequency, country population, mean R_0_, population density, rural/urban split, start date, end date, total confirmed cases, and the time to peak for each outbreak.

The time series data included daily or weekly case numbers for each outbreak, and an observed feature is a rapid initial growth followed by a slower phase. These two phases were used to constrain the two transmission mechanisms in the model; rapid initial transmission drive by human-human household contacts that is limited by the clustering of household contact networks, followed by slower but more widespread environment-human transmission, which must be high enough to take over but not so high as to produce rapid, widespread infection in most simulations. The human-human transmission risk per contact and proportionality constant for environment-human transmission in the model were adjusted to reproduce these features. The probability of symptomatic cholera and the probability of death given symptoms were adjusted to reproduce the average case fatality rate observed within the data, while fitting within the range of values described by the literature.

As Figure S8 indicates, most outbreaks are less than 1000 cases (after scaling to a model population of 50,000), but there is a small subset which grows to be much larger. In order to constrain cholera transmission in the absence of a vaccine response and produce (a) the rapid initial growth, (b) the slower secondary growth, and (c) a typically contained final size, the model contains WASH interventions to reduce transmission via both human-human and environment-human mechanisms (implemented as a relative reduction in the force of infection). These interventions are initialised with relatively low impact/coverage in the model, but after outbreak declaration the impact is strengthened as people in the model are assumed to prioritise sources of clean water and improve hygiene practices as much as possible during an outbreak. The timing and extent to which they increase following the detection of the outbreak was calibrated to reduce the rapid human-human transmission in households once cholera prevalence in the environment was high enough to drive the second phase of transmission. The WASH interventions were also calibrated to limit the environmental force of infection during the second phase of transmission, such that very large outbreaks were much less frequent than smaller outbreaks.

Figure S8 shows how the model captures the growth rate and range of outbreak sizes without a vaccine response, compared with the Johns Hopkins University data set.

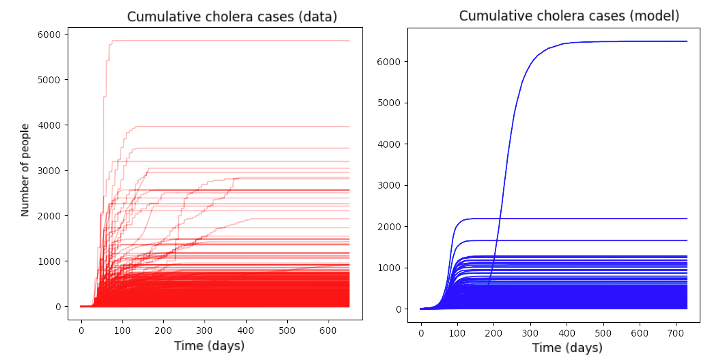

**Figure S8**: Comparison of Johns Hopkins University cholera outbreak time series (scaled to a population of 50,000) against an example set of model time series (using a population of 50,000 agents) after calibration. The model can reproduce the rapid initial epidemic growth and produces typically small outbreaks with some infrequent larger outbreaks, as is observed in the data.

Following this initial calibration process, for a given ORI response time and daily vaccination rate, outbreak simulations could be run to produce a range of stochastic outcomes that could be compared to observed outbreaks within the outbreak dataset. To produce the *Baseline* simulations the model was run with a response time of 105 days and a daily vaccination rate of 5100 doses, reflecting the mean values from the outbreak dataset[1]. The daily vaccination rate results in the ORI campaign running for seven days when 75% coverage is achieved in the at-risk population, and ten days when 100% coverage is achieved, which is informed by our outbreak dataset and in line with observed estimates[43]. Stochastic outbreak simulations were produced by running the model 1000 times for each prospective outbreak.

#### 3.7 Scenarios

To explore the impact of response time and achieved ORI coverage on outbreak outcomes, we explored two scenarios and two sensitivity analyses:

- *Baseline*; response time of 105 days, daily vaccination rate of 5100 doses; achieved ORI coverage of 75% in ages 1+ years,
- *Faster response time*; response time of 15, 30, 45, 60, 75, or 90 days, daily vaccination rate of 5100 doses; achieved ORI coverage of 75% in ages 1+ years,
- *Alternate coverage* (sensitivity analysis); response time of 105 days, daily vaccination rate of 5100 doses; achieved ORI coverage of 100% in ages 1+ years,
- *Alternate vaccination rate* (sensitivity analysis); response time of 105 days; daily vaccination rate of 48,997 doses, or daily vaccination rate of 1300 doses; achieved coverage of 75% in ages 1+ years.

The simulations used for each scenario are identical except for the scenario parameter value varied (ORI response time, achieved ORI coverage, or daily vaccination rate). The Baseline scenario assumes 75% at-risk population coverage was achieved by the ORI program, which is on the lower end of recorded estimates from coverage surveys for single-dose cholera vaccine coverage, and therefore may lead to underestimates in impact[43]. However, the estimates do not account for wastage and therefore may be overestimates of actual achieved coverage. The effect of this assumption was also investigated in our sensitivity analyses. The choices of alternate vaccination rates for the sensitivity analysis are based on the number of doses required to vaccinate the entire eligible model population in either one day or one month.

#### 3.8 Results

##### 3.8.1 Impact of response time

In the Results section of our manuscript, we show an area plot representing the proportion of cholera outbreak simulations which exceed certain case thresholds as a function of ORI response time in Figure 2. This plot also appears as Figure S10. The manuscript’s Results section also describes the proportional reduction in the mean cumulative cases and deaths for the set of cholera outbreaks with ORI response times of 15, 30, 45, 60, 75, and 90 days relative to the set of outbreaks with a response time of 105 days in Figure 4, the values for which are in Table S3. In these supplementary results we also show the distribution of cumulative cholera cases produced by the outbreak simulations (Figure S9).

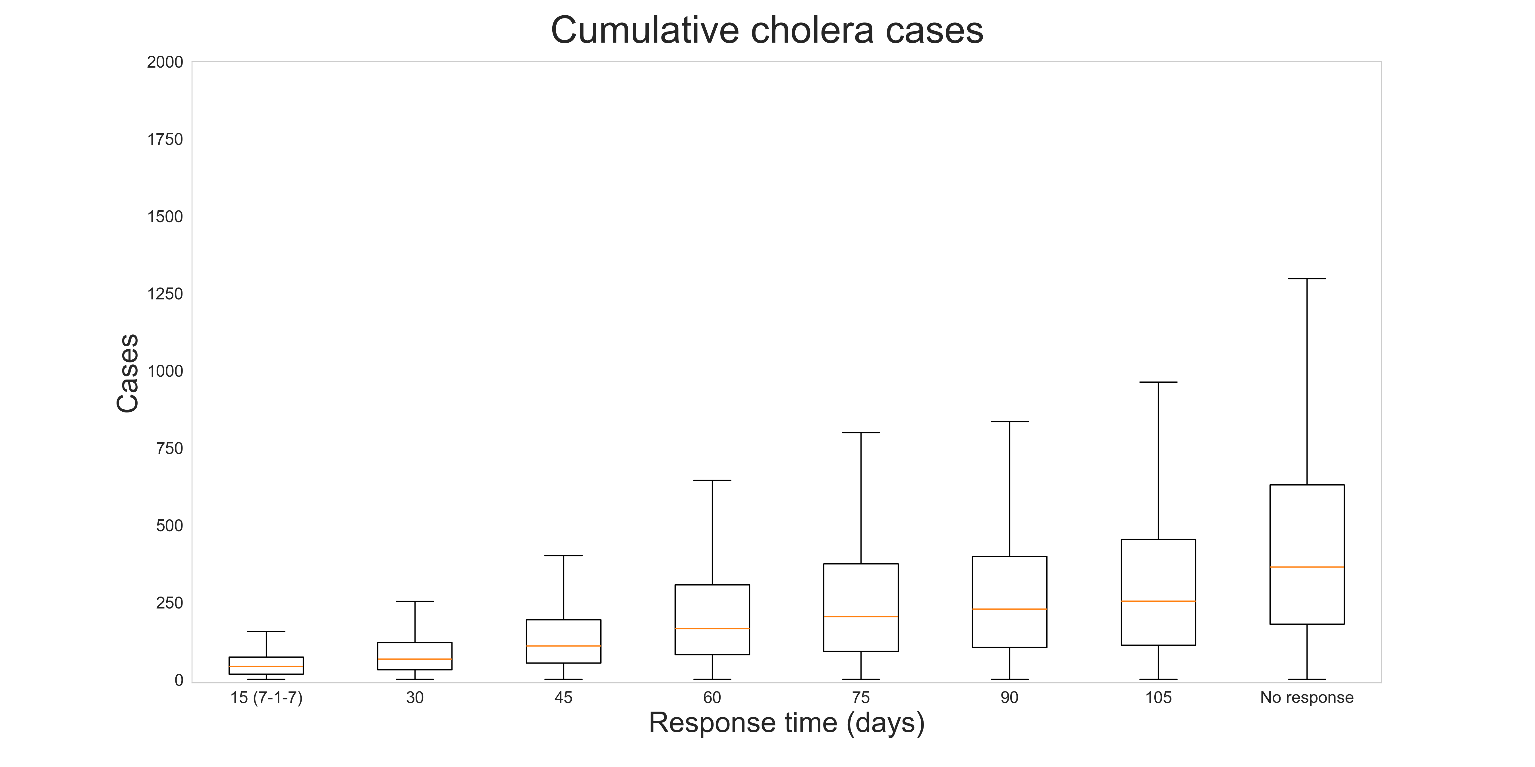

**Figure S9**: Distribution of cumulative cases produced by 1000 cholera outbreak simulations, given different ORI response times (15, 30, 45, 60, 75, 90, 105 days, or no response). Outbreaks without ORI produced the most cumulative cases, and when ORI occurred outbreaks which received faster responses tended to produce fewer cumulative cases.

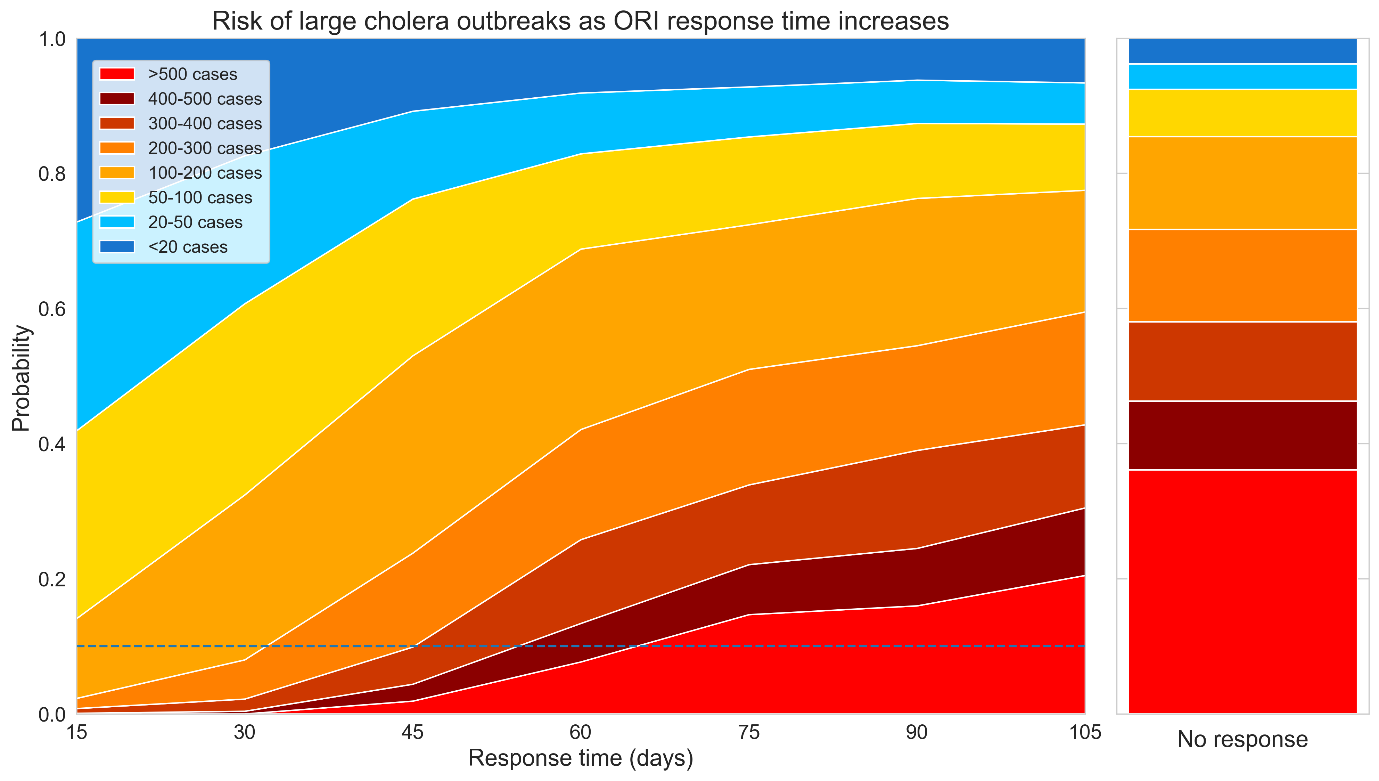

**Figure S10**: Area plot representing how the proportion of cholera outbreak simulations which exceed certain case thresholds (20, 50, 100, 200, 300, 400, or 500 cases) changes as a function of ORI response time. The dashed line represents a 10% threshold for outbreaks which exceed 500 cases, which is crossed when ORI takes more than 60 days to be initiated. The righthand stacked bar represents the distribution of outbreak sizes if no ORI is delivered, for reference.

**Table S3**: Proportional reduction in cholera cases, deaths, and DALYs relative to 105-day response.

| Response time (days) | Cases | Deaths | DALYs |
| --- | --- | --- | --- |
| 90 | 0.249 (0.222, 0.276) | 0.219 (0.193, 0.244) | 0.219 (0.193, 0.244) |
| 75 | 0.306 (0.277, 0.334) | 0.268 (0.241, 0.296) | 0.263 (0.236, 0.29) |
| 60 | 0.389 (0.359, 0.419) | 0.347 (0.317, 0.376) | 0.342 (0.312, 0.371) |
| 45 | 0.549 (0.518, 0.58) | 0.509 (0.478, 0.54) | 0.498 (0.467, 0.529) |
| 30 | 0.706 (0.678, 0.734) | 0.661 (0.632, 0.69) | 0.657 (0.627, 0.686) |
| 15 | 0.797 (0.772, 0.822) | 0.759 (0.732, 0.785) | 0.752 (0.725, 0.779) |

#### 3.9 Limitations

- Actual transmission might be heterogeneous and could be a factor that limits outbreak size: there are multiple sources of heterogeneity that the model cannot capture, across the populations, transmission networks, and environmental reservoirs for cholera. This includes:
  - Age-based contact networks beyond the household
  - Other highly connected contact networks (e.g., workplaces, schools, social groups)
  - Geographical heterogeneities in where people interact with contaminated water
  - Coverage and efficacy of WASH interventions
- High uncertainty around vaccine efficacy: only a small number of studies have investigated the efficacy or effectiveness of a single dose of oral cholera vaccines, with highly variable reported values. The efficacy value used in the model is from the most recent study with a large sample size and robust methods, but is for vaccine effectiveness rather than efficacy. As such it does not directly translate to the model’s parameter and this potentially leads to an overestimate of its impact. There was only a single study found which investigated single dose efficacy, and multiple experts suggested that the value was too low during validation of the model, so we aligned with the effectiveness value.
- Reporting of cholera cases: the model is calibrated to cumulative suspected cases reported for each outbreak:
  - This may be inflated by cases of acute watery diarrhea which are not due to cholera. If that is the case, then the model may overestimate the transmissibility of cholera which would produce overestimates of ORI impact.
  - On the other hand, reported cases are almost certainly an underestimate of true symptomatic infections due to underreporting, in which case the model may underestimate the transmissibility of cholera which would produce underestimates of ORI impact. We attempt to account for this with a calibrated case ascertainment rate of 20 – 25%, however there is little data to validate this against and it is likely highly variable across outbreaks.
- Uncertain case fatality rate: The model relies on 0·5% based on the observed mortality rate in our dataset, and the literature reports a mortality rate of less than 1% for treated cholera[44]. However, recent cholera outbreaks have had significantly higher fatality rates[45]. If the case fatality rate is greater than 0·5%, then the model may have underestimated deaths and vaccine impact.
- Uncertainty around the shapes of the epidemic curves for the cholera outbreaks which were included in the calibration, due to a lack of available data. While the epicurves produced by the model were calibrated to align with the epicurves shared by JHU, there were typically insufficient data to do this for all of the outbreaks included in the calibration.
- Uncertainty around the expected coverage achieved by ORI during an outbreak: It is unclear what level of population coverage is typically achieved by ORI campaigns, and it is likely that it would vary widely based on the outbreak context. Our assumption of 75% achieved ORI coverage would be optimistic if outbreaks occur in remote or otherwise hard to reach populations, and would be pessimistic if the population at-risk has a strong history of vaccine uptake. We attempt to understand the impact of this assumption in our *Alternate coverage* scenario.

### 4 Measles

#### 4.1 Background and motivation

Measles is an acute, highly contagious, viral airborne disease that can lead to severe complications and deaths[7]. Measles is so contagious that if one person has it, up to 90% of the people close to that person who are not immune will also become infected, with a basic reproduction number around 12 – 18[7, 46]. Symptoms include a high fever, cough, runny nose, and a rash all over the body. The highest burden is in unvaccinated or under vaccinated children under the age of five. The WHO states that measles vaccination averted 56 million deaths between 2000 and 2021. While outbreaks of measles are responded to with vaccines, ideally a sufficiently high level of immunity is maintained such that sustained transmission cannot occur, but this requires greater than 94% population immunity[47]. However, in 2022, only about 83% of the world’s children received one dose of measles vaccine by their first birthday through routine health services – the lowest since 2008[48].

A previous analysis investigating the historical impact of ORI programs across 51 measles outbreaks occurring between 2000 – 2023 found that ORI averted 4·01M (3·95M – 4·07M) cases and 20·0K (19·6K – 20·4K) deaths, representing 0 – 95% of the case and death burden for each measles outbreak, and being very dependent on routine vaccine coverage[3]. It found that ORI programs achieved the least impact in settings with very high baseline immunity because outbreaks in these settings were typically limited even in the absence of an immunisation response. Additionally, outbreaks which received faster responses tended to avert higher proportions of the expected measles burden, and that there is room for improvement as the mean response time was around four months.

#### 4.2 Model overview

The *Starsim* framework was used to create an agent-based model of measles among humans aged five years and under[39], with states for susceptible, exposed, infected, and recovered agents (Figure S11). Agents in the model represent humans, who begin as susceptible, and each day have a probability of becoming infected that depends on their immunity status and is proportional to the prevalence of infection among the population. Following infection, humans enter a latent infection ‘exposed’ state, before becoming infectious to others. Humans in the infectious state can recover and develop immunity to further infection during the modelled outbreak, and humans can die based on a disease-specific mortality rate. Agents can also be vaccinated, which will provide a level of immunity against future infection.

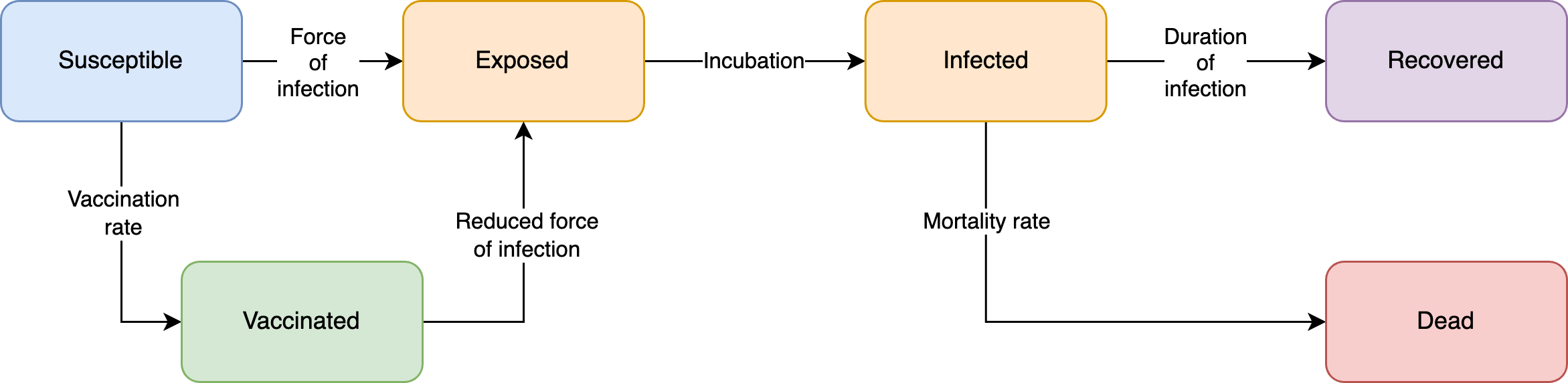

**Figure S11: Measles model schematic.** The Starsim framework was used to develop an agent-based model of measles among humans (S-E-I-R).

#### 4.3 Model population for simulated outbreaks

The model population for each outbreak simulation represents the entire population of the specific geographic locations where outbreaks have occurred between 2000 – 2023, based on the outbreak dataset[1]. For computational reasons the model contains a maximum of 50,000 agents.

Measles outbreaks predominantly occur in children under five years of age, as older people typically have immunity acquired either from vaccination or from prior infection[48]. As the ORI program target population is also typically limited to children under five years of age, we elected to only model this age group. The model was parametrised by age structure from the United Nations, Department of Economic and Social Affairs, Population Division[6] for ages zero to five years. The age distribution in the model was generated by summing over the single year age data for each country with an outbreak included in the outbreak dataset. Agents in the model are assigned integer ages, so to capture the vaccine eligibility of children older than nine months, 25% of children under one-year old were assumed eligible (i.e., assuming a uniform distribution of age within this group).

Additionally, the model considers protection against measles at birth for infants up to a certain age. It is estimated that the presence of maternal antibodies endured for a median of 2·61 – 3·78 months for infants of naturally infected women and 0·97 months for infants of vaccinated women[49]. Using the assumed routine vaccine coverage in the model, a weighted average was calculated to estimate the age where infants are no longer considered immune. It was then assumed that maternal immunity decreases linearly up to that age.

Transmission in the model occurred through community contacts between agents. These networks are randomly generated at each time step (representing a day in the model). Vaccine coverage, transmission risk, and disease outcomes were not modelled to vary by age given limited data from the outbreak settings. As we only model the 0–5-year-old population, some transmission between agents is indirect and mediated by older people is not captured in the model, and instead assume it makes up a proportion of the direct transmission used in the model.

#### 4.4 Diagnosis of cases, outbreak declaration and ORI

Agents within the model are assumed to seek a test when symptomatic, with a probability of 10% per day that they are symptomatic. Measles outbreaks are typically declared if there are five or more epidemiologically linked cases[50]. As we simulate a version of the population with a limited age range and do not model background cases, an outbreak is declared in the model after the diagnosis of a single agent, and once this occurs the ORI will begin after N-2 days, where N is the response time for a given outbreak. Some people with a diagnosed infection will undergo an isolation or otherwise limit their contact with others to reduce risk of onward transmission. However, for measles it is expected that effective isolation will be challenging, so we only include a small reduction in transmission for agents that are diagnosed. The outbreak is considered to have ended after 21 days with no new cases detected during a simulation.

#### 4.5 Calibration

Calibration involves estimating the transmissibility of measles in the model to produce outbreaks of a sufficient size, as well as the probability of death given an infection in order to capture the observed case fatality rate. For each outbreak used for the model calibration, an estimate of routine vaccine coverage is also required, which can be challenging as routine vaccine coverages can vary significantly across subnational regions of a country for various reasons. Eight of the 51 outbreaks included in the outbreak dataset reported a subnational area, including one outbreak with a reported subnational routine vaccine coverage. For the remaining seven outbreaks with a known subnational area, the Institute for Health Metrics and Evaluation (IHME)[51] modelled estimates of the first dose measles containing vaccine (MCV1) coverage (mean and confidence interval) were used to approximate the underlying routine vaccine coverage for each outbreak.

The outbreaks with known routine vaccine coverage were used to estimate the transmission parameter as follows. For each outbreak, simulations were run for a range of routine vaccine coverage and transmission parameters (and corresponding ORI response time and daily vaccination rates fixed based on the data). For each value of the transmission parameter, a model-estimated routine vaccine coverage was derived for each outbreak based on the maximum-likelihood estimate of the total number of cumulative cases[3]. The transmission parameter that was chosen was the one that minimised, across the eight outbreaks with known subnational coverage, the difference between the known subnational coverage and the model-estimated vaccine coverage (Figure S12).

For other outbreaks in the outbreak dataset with no reported subnational area, simulations were run with corresponding ORI response times and daily vaccination rates from the data[1], the transmission parameter estimated as above, and different levels of routine vaccine coverage. All other relevant model parameters were constrained by estimates from the literature.

To produce the *Baseline* simulations the model was run with a response time of 120 days and a daily vaccination rate of 4360 doses, reflecting the mean values from the outbreak dataset[1], across a range of routine vaccine coverage values (60%, 70%, 80%, 90%, or 100%). Outbreaks were simulated across these varying levels of routine coverage as representative setting archetypes, because routine coverage differs widely across outbreak contexts and it is important to understand the relationship between it and response time when considering ORI impact estimates. Stochastic outbreak simulations were produced by running the model 1000 times for each prospective outbreak.

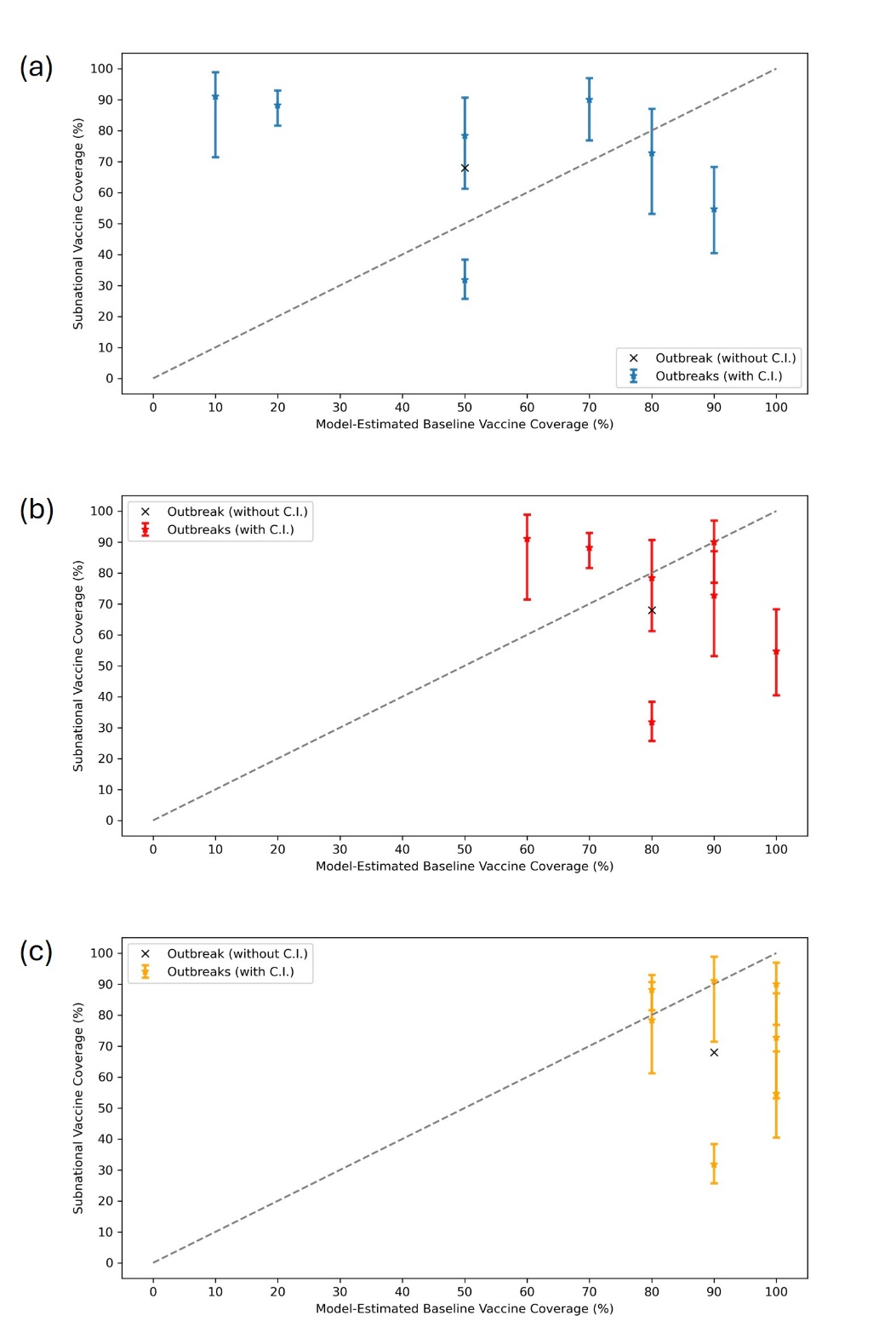

**Figure S12**: Measles calibration of transmission rate. Simulations with response time, vaccination rate, and routine vaccine coverage were run for different transmission parameters. Model-estimated vs. subnational vaccine coverage were then compared, and the final transmission parameter selected to minimise difference between model-estimated and subnational routine coverage. The transmission rate in (a) is too low as a subset of the outbreaks are assigned to routine coverages that are too low compared to the subnational coverages. The transmission rate in (c) is too high as many outbreaks are assigned to routine vaccine coverages that are too high compared to the subnational coverages. The transmission rate in (b) minimises the difference between subnational and model-estimated coverages. Bars indicate upper and lower values of Institute for Health Metrics and Evaluation (IHME) subnational measles containing vaccine dose one (MCV1) coverages, where available.

#### 4.6 Scenarios

To explore the impact of response time and achieved ORI coverage on outbreak outcomes, we explored two scenarios and two sensitivity analyses:

- *Baseline*; response time of 120 days, daily vaccination rate of 4360 doses; achieved ORI coverage of 100% in ages 9+ months,
- *Faster response time*; response time of 15, 30, 45, 60, 75, 90, or 105 days, daily vaccination rate of 4360 doses; achieved ORI coverage of 100% in ages 9+ months,
- *Alternate coverage* (sensitivity analysis); response time of 120 days, daily vaccination rate of 4360 doses; achieved ORI coverage of 50%, 60%, 70%, 80%, or 90% in ages 9+ months,
- *Alternate vaccination rate* (sensitivity analysis); response time of 120 days; daily vaccination rate of 47,812 doses, or daily vaccination rate of 1300 doses; achieved ORI coverage of 75% in ages 9+ months.

Each set of scenario simulations were run across the range of routine vaccine coverage values: 60%, 70%, 80%, 90%, or 100%. For a given routine coverage value, the simulations used for each scenario are identical except for the scenario parameter value varied (ORI response time, achieved ORI coverage, or daily vaccination rate). The *Baseline* scenario assumes 100% at-risk population coverage was achieved by the ORI program, which is informed by data provided by Gavi, the Vaccine Alliance, but may be an overestimate as it did not consider wastage. The effect of this assumption was investigated in our sensitivity analyses. The choices of alternate vaccination rates for the sensitivity analysis are based on the number of doses required to vaccinate the entire eligible model population in either one day or one month.

#### 4.7 Results

##### 4.7.1 Impact of response time

In the Results section of our manuscript, we show area plots representing the proportion of measles outbreak simulations which exceed certain case thresholds as a function of ORI response time for settings with 50% and 100% routine vaccine coverage in Figure 2. These plots also appear in Figure S15, alongside area plots for additional setting archetypes. The manuscript’s Results section also describes the proportional reduction in the mean cumulative cases and deaths for the set of measles outbreaks with ORI response times of 15, 30, 45, 60, 75, 90, and 105 days relative to the set of outbreaks with a response time of 120 days in Figure 5, the values for which are in Table S4. In these supplementary results we show the distribution of cumulative measles cases produced by the outbreak simulations (Figure S13), and a heatmap of the mean cumulative cases from the outbreak simulations for varying setting archetypes and ORI response times (Figure S14).

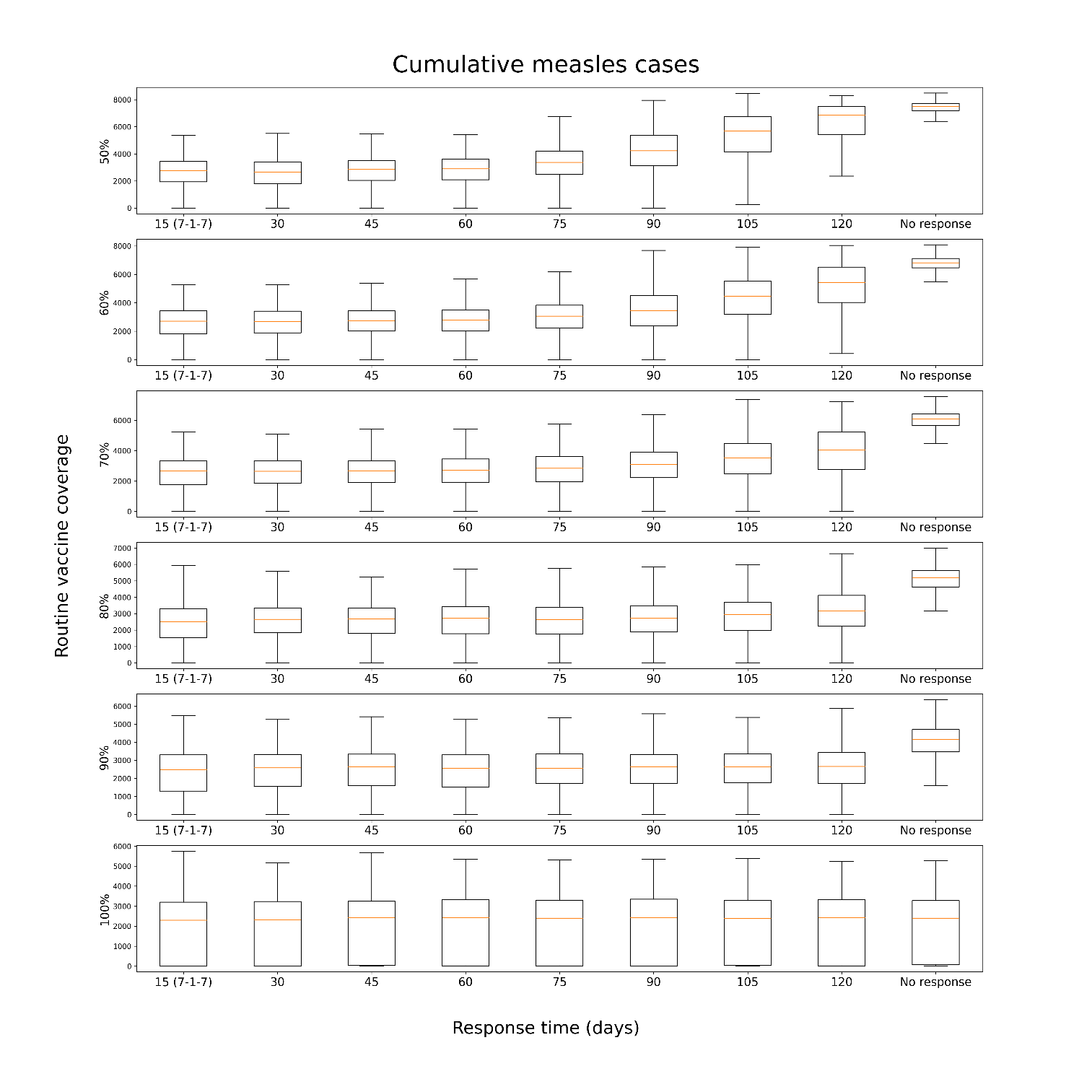

**Figure S13**: Distribution of cumulative cases produced by 1000 measles outbreak simulations, given different ORI response times (15, 30, 45, 60, 75, 90, 105, 120 days, or no response) and routine vaccine coverage in the model population before an outbreak (50%, 60%, 70%, 80%, 90%, or 100%). For each routine coverage below 100%, outbreaks without ORI produced the most cumulative cases, and when ORI occurred outbreaks which received faster responses tended to produce fewer cumulative cases, with the effect becoming more noticeable at lower coverages.

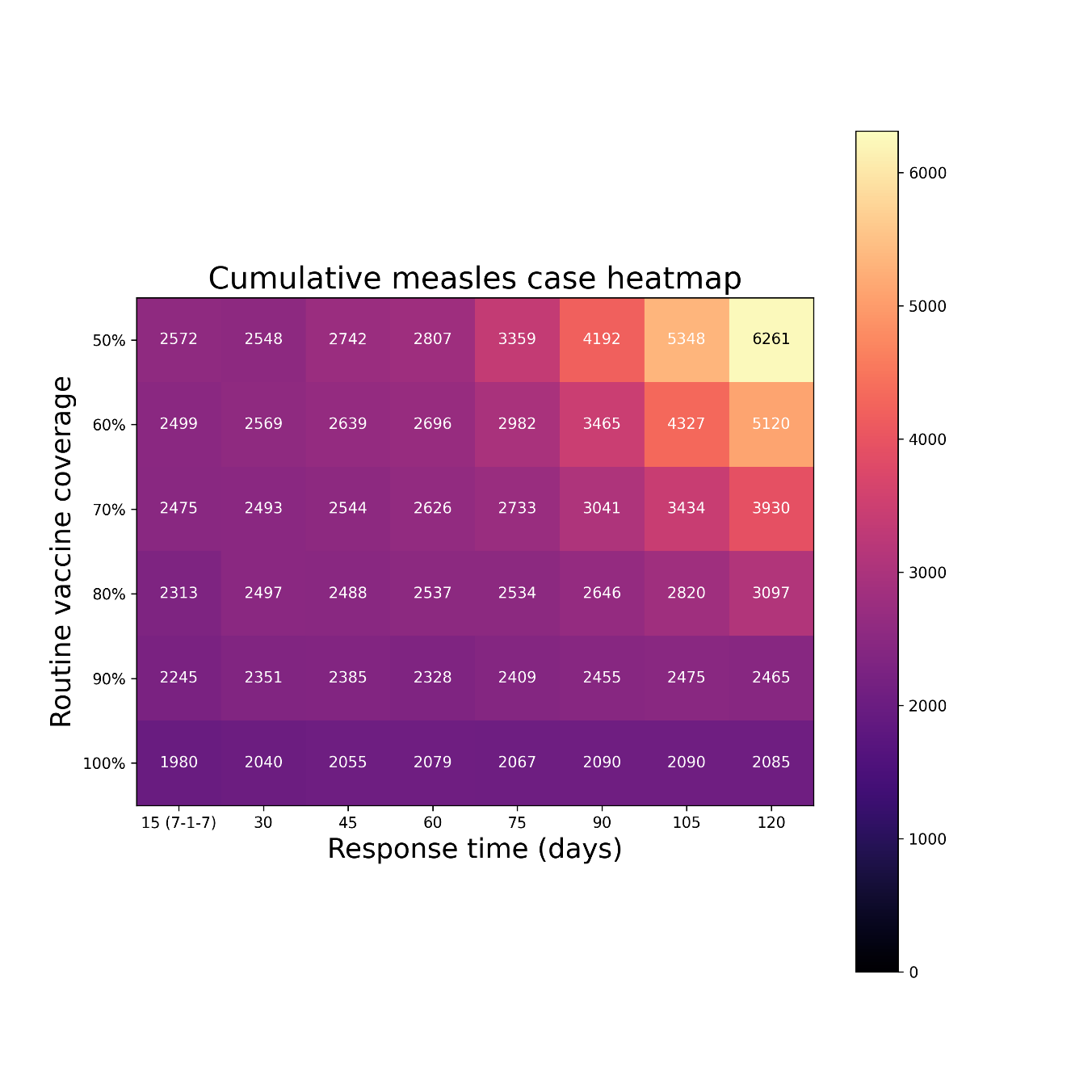

**Figure S14**: Heatmap of the mean cumulative cases by 1000 measles outbreak simulations, given different ORI response times (15, 30, 45, 60, 75, 90, 105, 120 days or no response) and routine vaccine coverage in the model population before an outbreak (50%, 60%, 70%, 80%, 90% or 100%). For each routine coverage below 100%, outbreaks which received faster responses of ORI tended to produce fewer cumulative cases, with the effect becoming more noticeable at lower coverages.

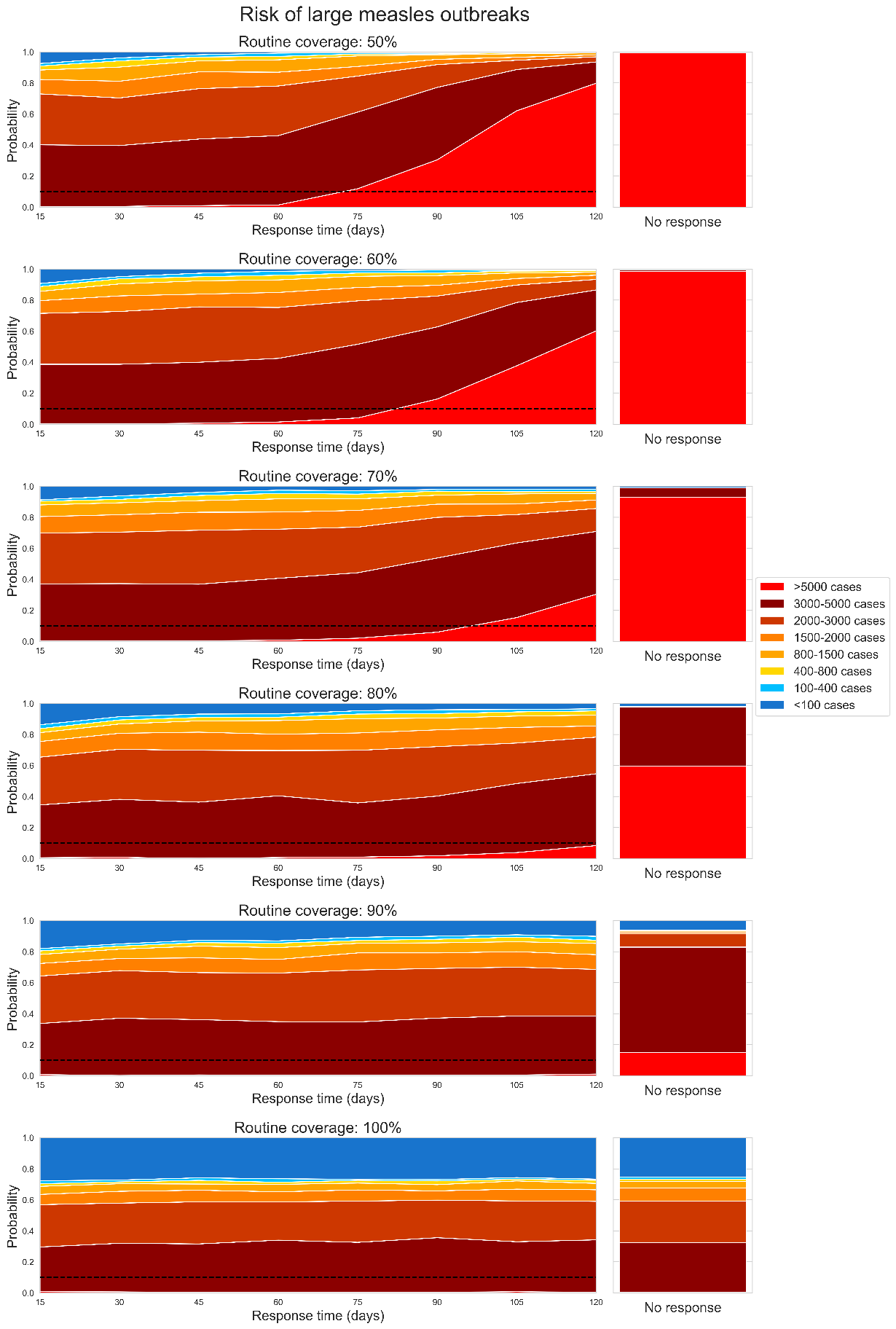

**Figure S15**: Area plot representing how the proportion of measles outbreak simulations which exceed certain case thresholds changes as a function of ORI response time, assuming a routine vaccine coverage of: a) 50%, b) 60%, c) 70%, d) 80%, e) 90%, and f) 100% before the outbreak is seeded. The dashed line represents a 10% threshold. The righthand stacked bar represents the distribution of outbreak sizes if no ORI is delivered, for reference.

**Table S4**: Proportional reduction in measles cases, deaths, and DALYs relative to 120-day response, for each choice of routine vaccine coverage.

| Response time (days) | Routine coverage | Cases | Deaths | DALYs |
| --- | --- | --- | --- | --- |
| 105 | 50% | 0.126 (0.105, 0.146) | 0.128 (0.107, 0.148) | 0.128 (0.107, 0.148) |
| 90 |  | 0.291 (0.263, 0.319) | 0.292 (0.264, 0.32) | 0.292 (0.264, 0.32) |
| 75 |  | 0.417 (0.386, 0.447) | 0.418 (0.387, 0.448) | 0.418 (0.387, 0.448) |
| 60 |  | 0.503 (0.472, 0.534) | 0.504 (0.473, 0.535) | 0.504 (0.473, 0.535) |
| 45 |  | 0.515 (0.484, 0.546) | 0.514 (0.483, 0.545) | 0.514 (0.483, 0.545) |
| 30 |  | 0.542 (0.511, 0.573) | 0.544 (0.513, 0.574) | 0.544 (0.513, 0.574) |
| 15 |  | 0.545 (0.514, 0.576) | 0.545 (0.514, 0.576) | 0.545 (0.514, 0.576) |
| 105 | 60% | 0.121 (0.101, 0.141) | 0.118 (0.098, 0.138) | 0.118 (0.098, 0.138) |
| 90 |  | 0.265 (0.238, 0.293) | 0.268 (0.241, 0.296) | 0.268 (0.241, 0.296) |
| 75 |  | 0.346 (0.316, 0.375) | 0.347 (0.317, 0.376) | 0.347 (0.317, 0.376) |
| 60 |  | 0.399 (0.369, 0.43) | 0.401 (0.371, 0.432) | 0.401 (0.371, 0.431) |
| 45 |  | 0.409 (0.378, 0.439) | 0.409 (0.378, 0.439) | 0.409 (0.378, 0.439) |
| 30 |  | 0.425 (0.394, 0.455) | 0.423 (0.392, 0.453) | 0.423 (0.392, 0.453) |
| 15 |  | 0.442 (0.411, 0.473) | 0.444 (0.413, 0.475) | 0.444 (0.413, 0.475) |
| 105 | 70% | 0.084 (0.067, 0.102) | 0.087 (0.069, 0.104) | 0.085 (0.068, 0.103) |
| 90 |  | 0.155 (0.132, 0.177) | 0.16 (0.137, 0.182) | 0.16 (0.137, 0.182) |
| 75 |  | 0.217 (0.191, 0.242) | 0.224 (0.198, 0.25) | 0.224 (0.198, 0.25) |
| 60 |  | 0.242 (0.215, 0.268) | 0.246 (0.219, 0.273) | 0.246 (0.219, 0.273) |
| 45 |  | 0.254 (0.227, 0.281) | 0.261 (0.233, 0.288) | 0.26 (0.233, 0.287) |
| 30 |  | 0.27 (0.243, 0.298) | 0.272 (0.245, 0.3) | 0.272 (0.245, 0.3) |
| 15 |  | 0.276 (0.249, 0.304) | 0.285 (0.257, 0.313) | 0.284 (0.256, 0.312) |
| 105 | 80% | 0.055 (0.041, 0.069) | 0.056 (0.042, 0.07) | 0.056 (0.041, 0.07) |
| 90 |  | 0.099 (0.08, 0.117) | 0.104 (0.085, 0.123) | 0.104 (0.085, 0.123) |
| 75 |  | 0.121 (0.1, 0.141) | 0.124 (0.103, 0.144) | 0.124 (0.103, 0.144) |
| 60 |  | 0.128 (0.107, 0.148) | 0.13 (0.109, 0.151) | 0.131 (0.11, 0.152) |
| 45 |  | 0.132 (0.111, 0.153) | 0.14 (0.118, 0.161) | 0.14 (0.118, 0.161) |
| 30 |  | 0.14 (0.118, 0.161) | 0.143 (0.121, 0.164) | 0.144 (0.122, 0.165) |
| 15 |  | 0.203 (0.178, 0.227) | 0.202 (0.177, 0.226) | 0.202 (0.177, 0.227) |
| 105 | 90% | 0 (0.0, 0.0) | 0 (0.0, 0.0) | 0 (0.0, 0.0) |
| 90 |  | 0 (0.0, 0.0) | 0 (0.0, 0.0) | 0 (0.0, 0.0) |
| 75 |  | 0.011 (0.004, 0.017) | 0.007 (0.002, 0.012) | 0.007 (0.002, 0.012) |
| 60 |  | 0.037 (0.025, 0.048) | 0.03 (0.019, 0.04) | 0.03 (0.019, 0.04) |
| 45 |  | 0.029 (0.019, 0.039) | 0.027 (0.017, 0.037) | 0.027 (0.017, 0.036) |
| 30 |  | 0.04 (0.028, 0.052) | 0.037 (0.025, 0.048) | 0.037 (0.025, 0.048) |
| 15 |  | 0.083 (0.065, 0.1) | 0.073 (0.057, 0.089) | 0.075 (0.059, 0.091) |
| 105 | 100% | 0.001 (0, 0.002) | 0 (0.0, 0.0) | 0 (0.0, 0.0) |
| 90 |  | 0.004 (0, 0.007) | 0.006 (0.001, 0.01) | 0.004 (0, 0.007) |
| 75 |  | 0.004 (0, 0.007) | 0.007 (0.002, 0.012) | 0.004 (0, 0.007) |
| 60 |  | 0.009 (0.003, 0.015) | 0.006 (0.001, 0.011) | 0.009 (0.003, 0.015) |
| 45 |  | 0.004 (0, 0.007) | 0.003 (0, 0.005) | 0.003 (0, 0.005) |
| 30 |  | 0.013 (0.006, 0.02) | 0.011 (0.004, 0.017) | 0.012 (0.005, 0.018) |
| 15 |  | 0.021 (0.012, 0.03) | 0.016 (0.008, 0.023) | 0.021 (0.012, 0.03) |

#### 4.8 Limitations

- High uncertainty around case ascertainment: The model relies on 20% based on the ratio of cumulative cases and the target population size as observed in our data set. However, the literature reports a case ascertainment rate of 1 – 5% for measles[52]. If the case ascertainment rate is much lower than 20%, then the model may have underestimated cases, deaths, and vaccine impact.
- Low outbreak declaration threshold and optimistic assumption about outbreak detection time. The model uses a single detected case of measles (which is detected one day after being introduced) to declare an outbreak. These assumptions are highly optimistic compared to standard practice, and may lead to an overestimation of impact from the ORI in the simulations. However, as the results indicate little impact on outbreak outcomes for the fastest ORI response times explored (those less than 60 days), any potential overestimation is expected to be minimal.
- Uncertain case fatality rate: The model relies on 0·5% based on the observed mortality rate in our dataset, and the literature reports a mortality rate of 2·2% for measles[53] differentiating between community-based and hospital-based settings. If the case fatality rate is greater than 0·5%, then the model may have underestimated deaths and vaccine impact.
- Uncertainty around vaccine efficacy: The efficacy value used in the model is from a study estimating the vaccine effectiveness in infants younger and older than nine months but is for vaccine effectiveness rather than efficacy[54]. As such it does not directly translate to the model’s parameter and this potentially leads to an overestimate of its impact.
- Uncertainty around re-vaccination: Re-vaccination of a proportion of the target population is expected as prior determination of the vaccination status is challenging in LMICs. The model incorporates re-vaccination of agents; however, there is no change in immunity. The results might therefore underestimate the impact of the ORI.
- Modelling young children only: The model considers young children, under five years of age. However, there could be important differences in transmission among older people, and indirect effects due to transmission from adults to children that are not captured.
- Uncertainty around the shapes of the epidemic curves for the measles outbreaks which were included in the calibration, due to a lack of available data. We were unable to calibrate the model against detailed timeseries data for the outbreaks included in the calibration dataset, only the cumulative cases and outbreaks duration. As such, the model may not accurately reflect the impact of the ORI response time if it is over or underestimating the number of cases which have occurred during an outbreak before ORI programs can be implemented.
- Uncertainty around the expected coverage achieved by ORI during an outbreak: It is unclear what level of population coverage is typically achieved by ORI campaigns, and it is likely that it would vary widely based on the outbreak context. Our assumption of 100% achieved coverage would be optimistic if outbreaks occur in remote or otherwise hard to reach populations, however based on data from Gavi, the Vaccine Alliance provided as a part of our outbreak dataset the number of vaccines delivered during ORI closely matches the estimated population at-risk for most outbreaks[3]. We attempt to understand the impact of this assumption in our *Alternate coverage* scenario.

### 5 Yellow fever

#### 5.1 Background and motivation

Yellow fever is an epidemic-prone mosquito-borne vaccine preventable disease that is transmitted to humans by the bites of infected mosquitoes, mostly from bites occurring during the day[55]. WHO considers it a high-impact high-threat disease that has the potential to spread internationally[55]. Yellow fever outbreaks have been recorded in 34 countries in Africa and 13 countries in Central and South America and are classified as either domestic (around houses), sylvatic (in forests or jungles) or semi-domestic outbreaks (both habitats)[55]. Outbreaks can also be differentiated by the transmission cycle, with ‘spillover’ outbreaks involving non-human primates and *Aedes africanus, Haemagogus spp.* and *Sabethes spp.* mosquitos, and ‘human-to-human’ outbreaks with transmission mediated by *Aedes aegypti* mosquitos[56]. The latter transmission cycle is typically associated with large, high-risk outbreaks and we therefore focus on outbreaks involving only humans in this study.

In countries where yellow fever occurs, the WHO strongly recommends routine vaccination for everyone older than nine months, and prevention of outbreaks in affected regions requires at least 80% vaccine coverage of the population at-risk[57]. A previous analysis investigating the historical impact of ORI programs across 88 yellow fever outbreaks occurring between 2000 – 2023 found that ORI averted 1·50M (1·42M – 1·58M) cases and 300K (284K – 316K) deaths, representing 0 – 98% of the case and death burden for each yellow fever outbreak, and being very dependent on routine vaccine coverage and environmental suitability to the mosquito population[3]. It found that ORI programs achieved the least impact in settings with high baseline immunity or low environmental suitability because outbreaks in these settings were typically limited even in the absence of an immunisation response. Additionally, outbreaks which received faster responses tended to avert higher proportions of the expected yellow fever burden, and that there is room for improvement as the mean response time was around three months.

#### 5.2 Model overview

The *Starsim* framework was used to create an agent-based model of yellow fever among humans[39], with states for susceptible, exposed, infected (either mild or severe), and recovered agents, which was coupled to a compartmental susceptible-exposed-infectious-susceptible (S-E-I-S) model among mosquitos so that risk of indirect, vector-borne transmission to humans could be approximated by a dynamic parameter for prevalence among mosquitoes (represented in Figure S16).

Agents in the model represent humans, who begin as susceptible or vaccinated, and each day have a probability of becoming infected that is proportional to the prevalence of infection among the mosquito population, and that further depends on their vaccination status. Following infection, humans enter a latent infection ‘exposed’ state, before developing severe or non-severe disease and becoming infectious to mosquitoes. Humans in either of these infectious states can recover and develop immunity, and humans in the severe infectious state can die based on a disease-specific mortality rate. We did not disaggregate asymptomatic infections because little data is available to inform differences in disease duration and infectiousness, and because testing behaviour is mainly driven by severe vs non-severe disease.

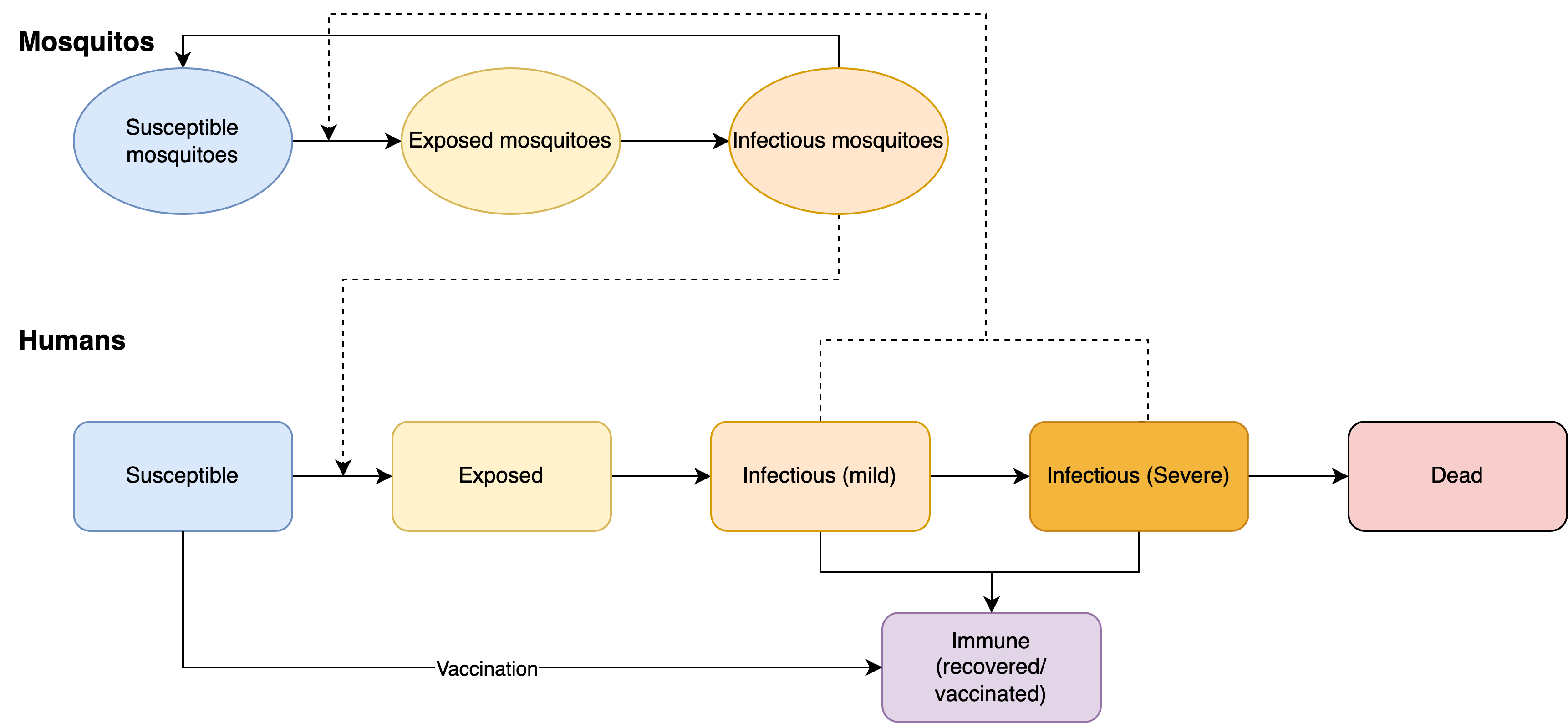

**Figure S16**: Yellow fever model schematic. The Starsim framework was used to develop an agent-based model of yellow fever among humans (S-E-I-R), which was paired with a dynamic compartmental model of infection among mosquitoes to parametrise risk of environmental transmission.

#### 5.3 Model population for simulated outbreaks

The model population for each outbreak simulation represents the entire population of the specific geographic locations where outbreaks have occurred between 2000 – 2023, based on the outbreak dataset[1]. For computational reasons the model contains a maximum of 50,000 agents. For this model, the proportion of the population eligible for the vaccine corresponds to ~97·8%, excluding children younger than nine months.

The model population for each outbreak was parametrised by age structure from the United Nations, Department of Economic and Social Affairs, Population Division[6], which provides an age distribution in one-year increments. The age distribution in the model was generated by summing over the single year age data for each country with an outbreak included in the outbreak dataset[1]. The routine vaccination coverage was estimated from the WHO/UNICEF Estimates of National Immunization Coverage (WUENIC)[58]. Agents in the model are assigned integer ages, so to capture the vaccine eligibility of children older than nine months[55], 25% of children under one year old were assumed eligible (i.e., assuming a uniform distribution of age within this group). Aside from vaccine eligibility for children less than nine months old, vaccine coverage, transmission risk, and disease outcomes were not modelled to vary by age given limited data from the outbreak settings. Transmission in the model occurred through interactions with the mosquito population (e.g., per day probability of infection that depended on prevalence) rather than direct human-human contact, hence no contact network structure among agents in the model was required.

#### 5.4 Diagnosis of cases, outbreak declaration and ORI

An estimated 12% of infections become severe[59], and so assuming only severe cases get reported a 12% case ascertainment rate was used[56]. The smallest outbreak in the outbreak dataset recorded one case (i.e., one severe case)[1], indicating that outbreaks are declared with just one diagnosis. The WHO also states that outbreaks are declared in the ‘presence of at least one confirmed case of yellow fever’[60]. Therefore, an outbreak was considered declared as soon as one agent progressed to the severe state, and all severe cases were assumed to be reported. The ORI then begins after N-2 days, where N is the response time for a given outbreak. The outbreak is considered to have ended after 21 days with no new cases detected during a simulation.

As we assume a high diagnosis rate for severe infections and negligible diagnosis for non-severe infections, in the following analysis ‘cases’ is synonymous with the number of severe infections.

#### 5.5 Calibration

Calibration involves estimating the proportionality constants Beta linking transmission from mosquitoes to humans and from humans to mosquitoes. The transmission from humans to mosquitoes affects the amount of change in the mosquito prevalence, which gets updated at each time step (representing a day) in the simulation using the human prevalence. The transmission from mosquitoes to humans and the mosquito prevalence control the probability of a susceptible agent becoming infected. The calibration of the proportionality constants was done by using known estimates of the reproduction number R_0_ (provided by Fraser et al. (2024)[61]) and the final size equation:

$$\pi=1- e^{-R_{0}\pi}$$

The final size equation links R_0_ to the expected cumulative number of infections when an outbreak is simulated in a naïve population in an SEIR model. In a naïve population, the transmission between mosquitoes to humans and humans to mosquitoes is likely to be very similar and so assumed to be equivalent. Using the maximum estimated R_0_ of 1·45, we expect that ~55% of a naïve population gets infected in the absence of a response. In the model, the overall transmission probability was therefore calibrated such that this occurred when an outbreak was simulated without any prior immunity or any response.

For each outbreak used for the model calibration, an estimate of routine vaccine coverage is also required, which were based on WUENIC data[58]. However, the overall R_0_ value and routine vaccine coverage alone do not capture variability in transmission levels for different settings, with differences likely arising from climate and other factors that influence heterogeneity in transmission across settings. To account for this, an additional calibration variable was introduced to classify settings as low, medium-low, medium, medium-high, high transmission risks, implemented as factor multipliers on the calibrated Beta value, to modify mosquito-to-human transmission. These multiplying factors are 0·5, 0·75, 1·0, 1·25, and 1·5, and were determined during calibration against the outbreak dataset[1]. All other relevant model parameters were constrained by estimates from the literature.

To produce the *Baseline* simulations the model was run with an ORI response time of 105 days and a daily vaccination rate of 6560 doses, reflecting the mean values from the outbreak dataset[1], across a range of routine vaccine coverage values (40%, 50%, 60%, 70% or 80%) and transmission modifiers (low, medium-low, medium, medium-high, high). Outbreaks were simulated across these varying levels of routine coverage and transmission modifier as representative setting archetypes, as these parameters differ widely across outbreak contexts and it is important to understand the effect that they and response time have on the ORI impact estimates. Stochastic outbreak simulations were produced by running the model 1000 times for each prospective outbreak.

#### 5.6 Scenarios

To explore the impact of response time and achieved ORI coverage on outbreak outcomes, we explored two scenarios and two sensitivity analyses:

- *Baseline*; response time of 105 days; daily vaccination rate of 6560 doses; achieved coverage of 75% in ages 9+ months,
- *Faster response time*; response time of 15, 30, 45, 60, 75, or 90 days; daily vaccination rate of 6560 doses; achieved coverage of 75% in ages 9+ months,
- *Alternate coverage* (sensitivity analysis); response time of 105 days; daily vaccination rate of 6560 doses; achieved coverage of 100% in ages 9+ months,
- *Alternate vaccination rate* (sensitivity analysis); response time of 105 days; daily vaccination rate of 48,544 doses, or daily vaccination rate of 1300 doses; achieved ORI coverage of 75% in ages 9+ months.

Each set of scenario simulations were run across the range of routine vaccine coverage values: 40%, 50%, 60%, 70%, or 80% and transmission modifiers: low, medium-low, medium, medium-high, and high. For a given routine coverage value and transmission modifier, the simulations used for each scenario are identical except for the scenario parameter value varied (ORI response time, achieved ORI coverage, or daily vaccination rate). The Baseline scenario assumes 75% at-risk population coverage was achieved by the ORI program, which is in the range of estimates from the WHO[62] but may be an underestimate. The effect of this assumption was investigated in our sensitivity analyses. The choices of alternate vaccination rates for the sensitivity analysis are based on the number of doses required to vaccinate the entire eligible model population in either one day or one month.

#### 5.7 Results

##### 5.7.1 Impact of response time

In the Results section of our manuscript, we show area plots representing the proportion of yellow fever outbreak simulations which exceed certain case thresholds as a function of ORI response time for settings with 40% and 80% routine vaccine coverage and with a mosquito-human transmission modifier of 1·5 and 0·5, respectively, in Figure 2. These plots also appear in Figure S21 and Figure S22, alongside area plots for additional setting archetypes. The manuscript’s Results section also describes the proportional reduction in the mean cumulative cases and deaths for the set of yellow fever outbreaks with ORI response times of 15, 30, 45, 60, 75, and 90 days (and disaggregated by routine vaccine coverage) relative to the set of outbreaks with a response time of 105 days in Figure 6, the values for which are in Table S5. Figure S23 shows an equivalent plot, disaggregated by transmission modifier and with values shown in Table S6. In these supplementary results we show the distribution of cumulative yellow fever cases produced by the outbreak simulations (Figure S17 and Figure S18), and heatmaps of the mean cumulative cases from the outbreak simulations for varying setting archetypes and ORI response times (Figure S19 and Figure S20).

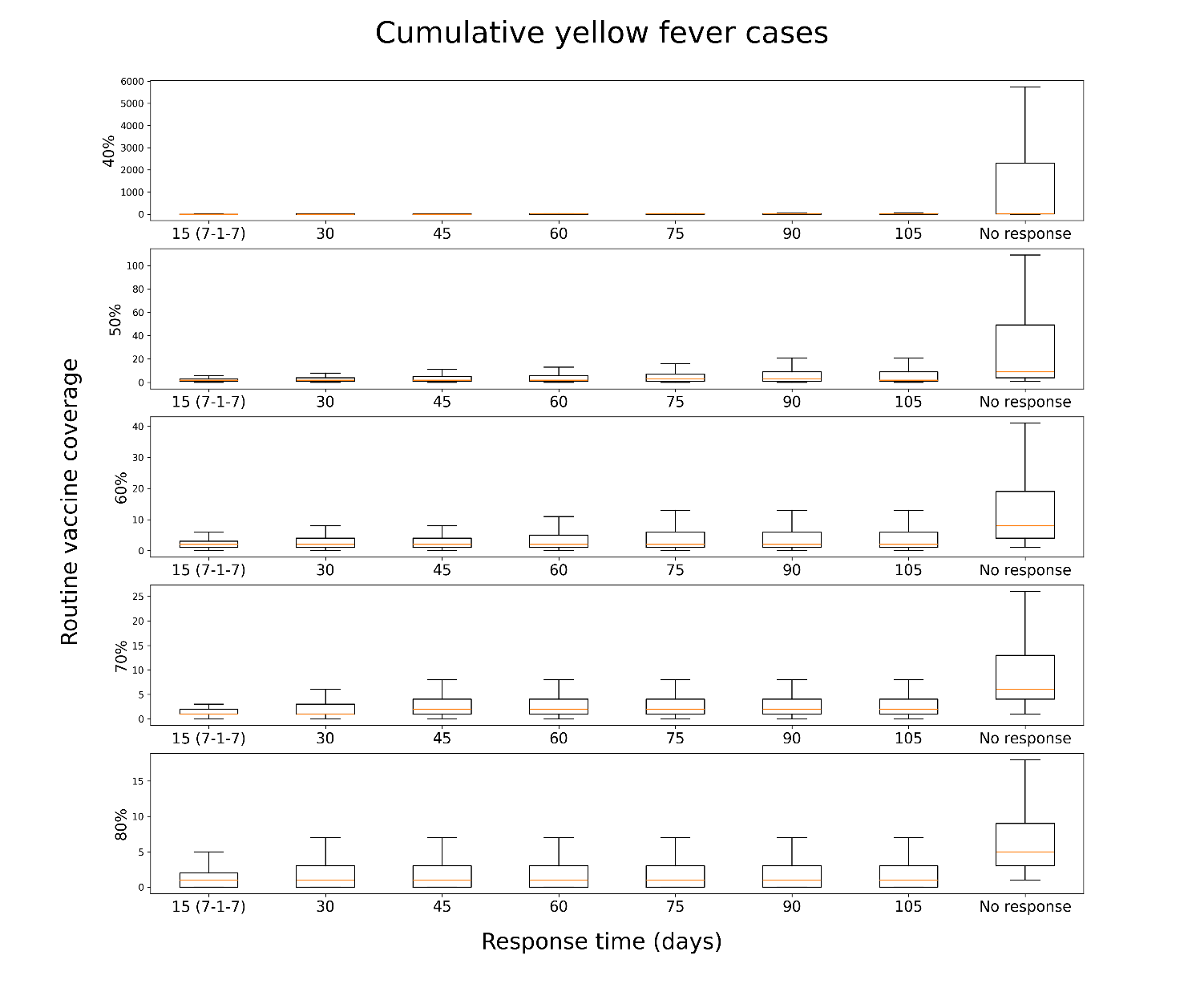

**Figure S17**: Distribution of cumulative cases produced by 1000 yellow fever outbreak simulations, given different ORI response times (15, 30, 45, 60, 75, 90, 105 days, or no response) and routine vaccine coverage in the model population before an outbreak (40%, 50%, 60%, 70%, or 80%), assuming a transmission modifier of 1·0 for the mosquito vectors. Outbreaks without ORI produced the most cumulative cases, and when ORI occurred outbreaks which received faster responses tended to produce fewer cumulative cases, with the effect becoming more noticeable at lower coverages.

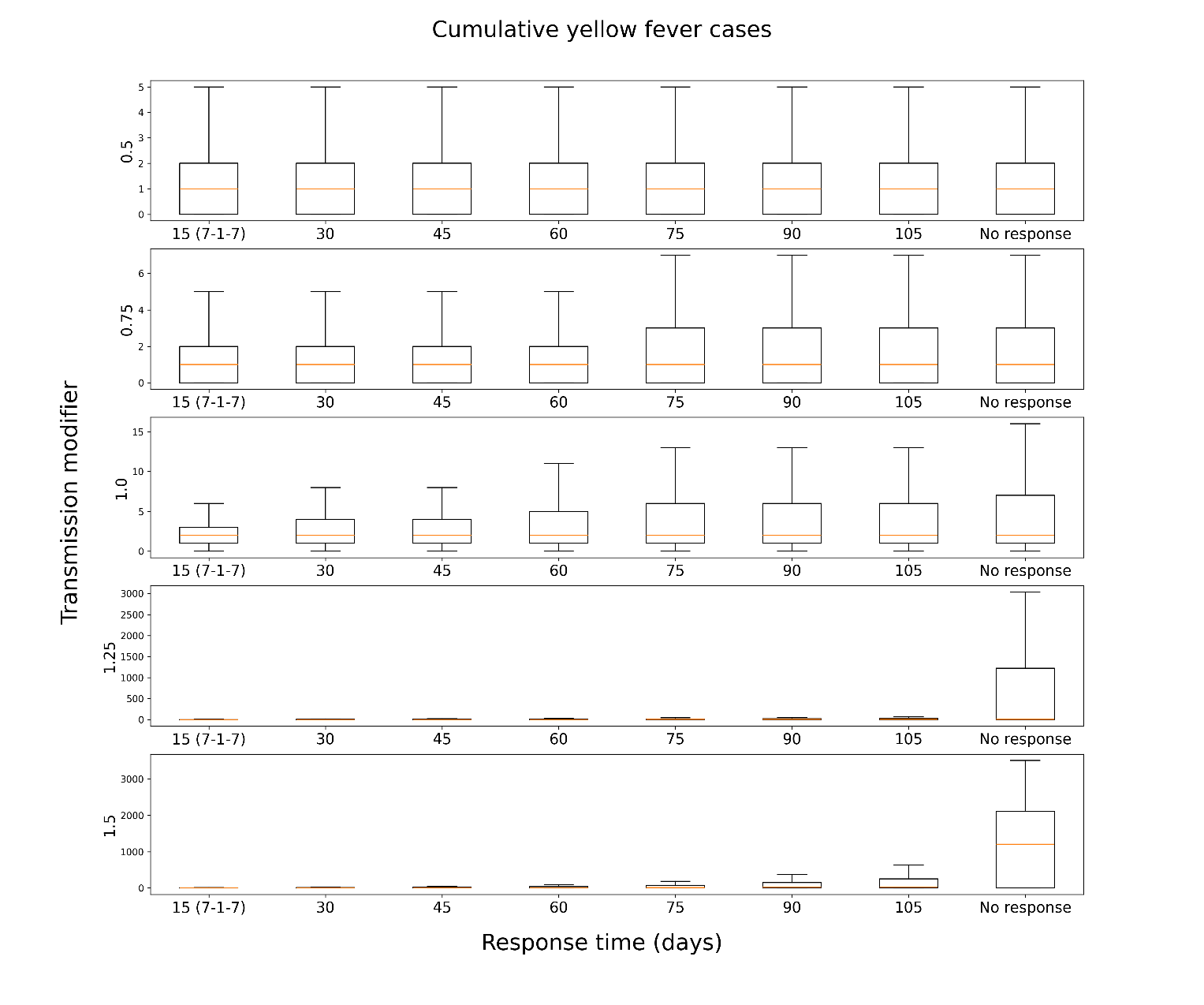

**Figure S18**: Distribution of cumulative cases produced by 1000 yellow fever outbreak simulations, given different ORI response times (15, 30, 45, 60, 75, 90, 105 days, or no response) and transmission modifier for the mosquito vectors in the model population before an outbreak (0·5, 0·75, 1·0, 1·25, or 1·5), assuming a routine vaccine coverage of 60%. Outbreaks without ORI produced the most cumulative cases, and when ORI occurred outbreaks which received faster responses tended to produce fewer cumulative cases, with the effect becoming more noticeable at higher transmission modifiers.

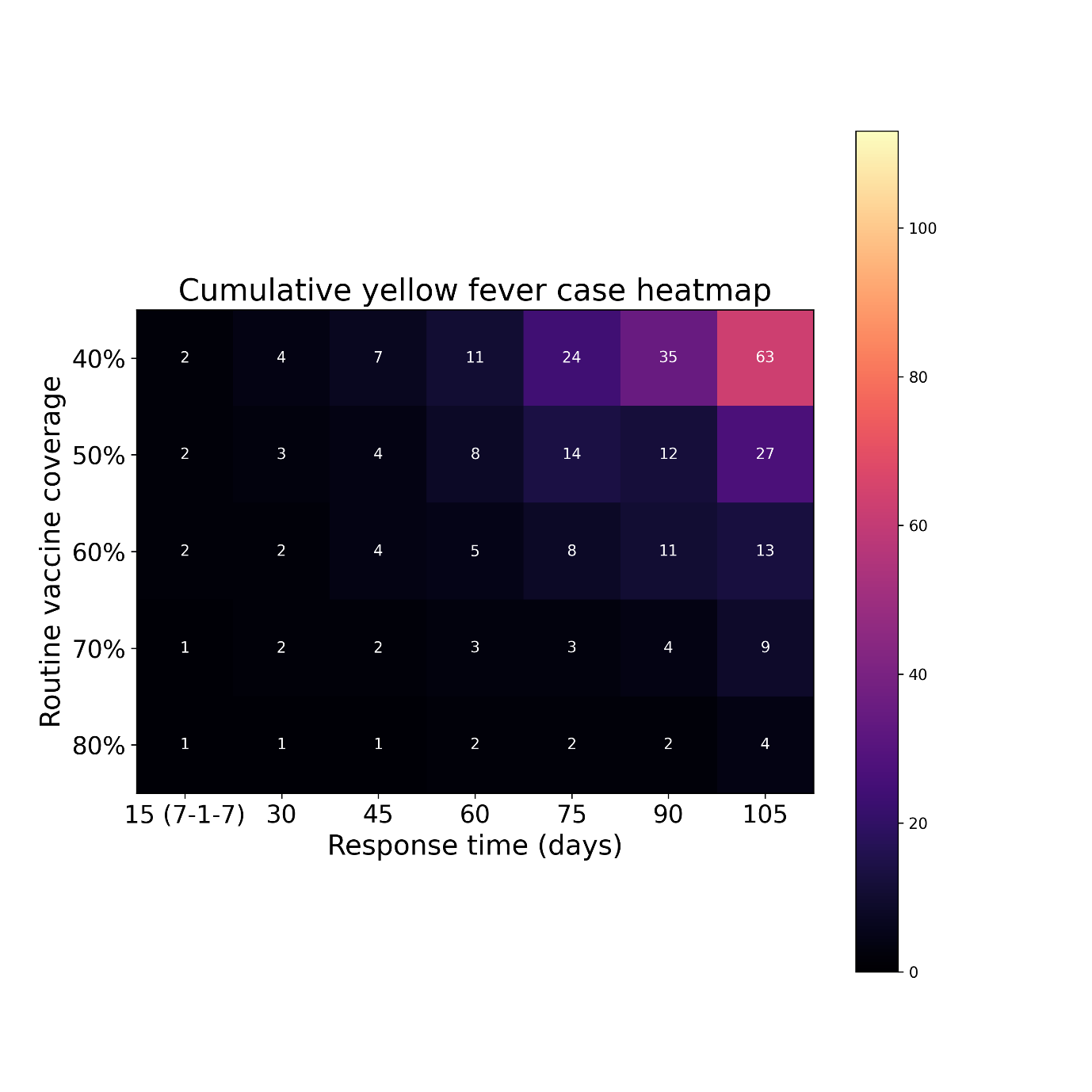

**Figure S19**: Heatmap of the mean cases produced by 1000 yellow fever outbreak simulations, given different ORI response times (15, 30, 45, 60, 75, 90, or 105 days) and routine vaccine coverage in the model population before an outbreak (40%, 50%, 60%, 70%, or 80%), assuming a transmission modifier of 1·0 for the mosquito vectors. Outbreaks without ORI produced the most cumulative cases, and when ORI occurred outbreaks which received faster responses tended to produce fewer cumulative cases, with the effect becoming more noticeable at lower coverages.

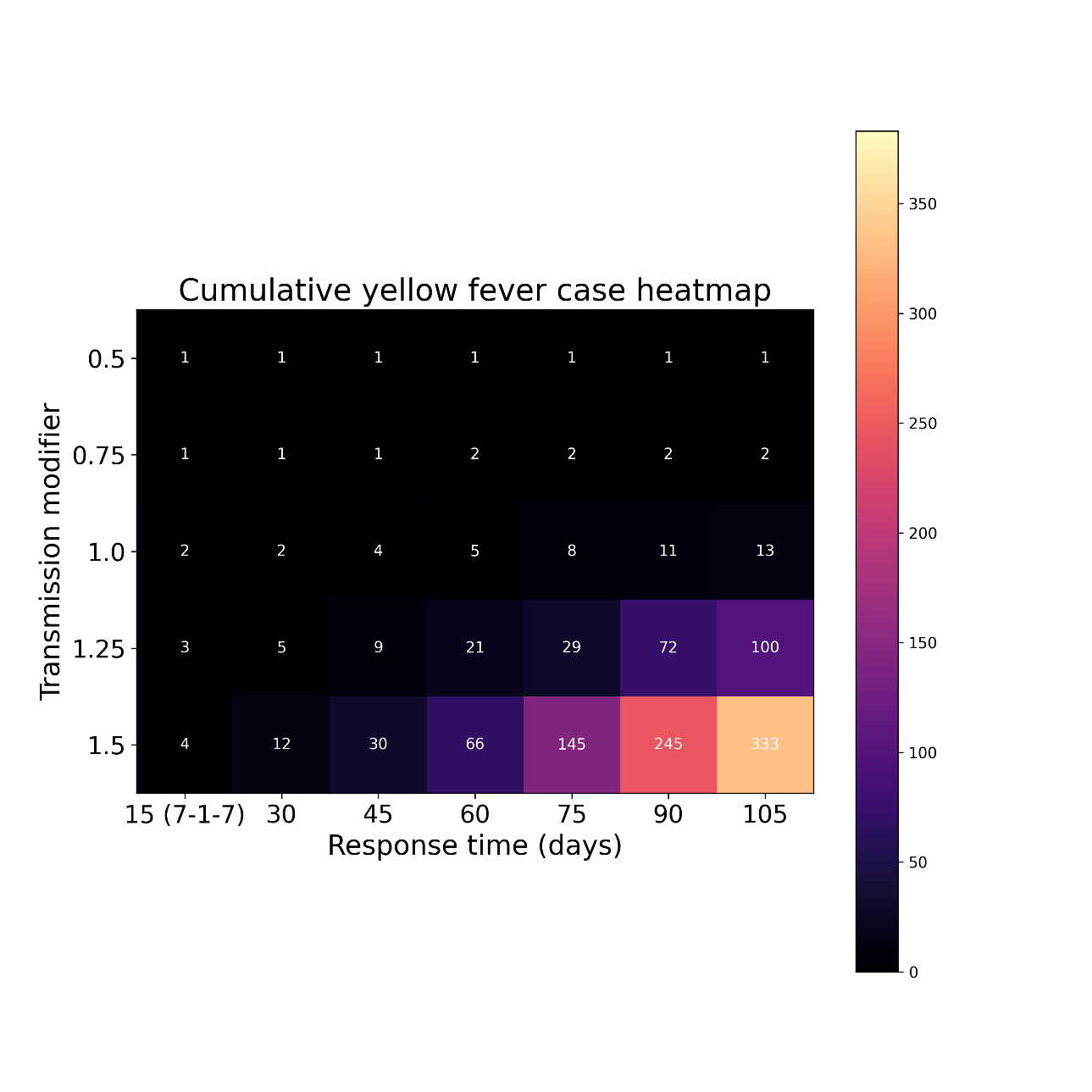

**Figure S20**: Heatmap of the mean cases produced by 1000 yellow fever outbreak simulations, given different ORI response times (15, 30, 45, 60, 75, 90, or 105 days) and transmission modifier for the mosquito vectors in the model population before an outbreak (0·5, 0·75, 1·0, 1·25,or 1·5), assuming a routine vaccine coverage of 60%. Outbreaks without ORI produced the most cumulative cases, and when ORI occurred outbreaks which received faster responses tended to produce fewer cumulative cases, with the effect becoming more noticeable at higher transmission modifiers.

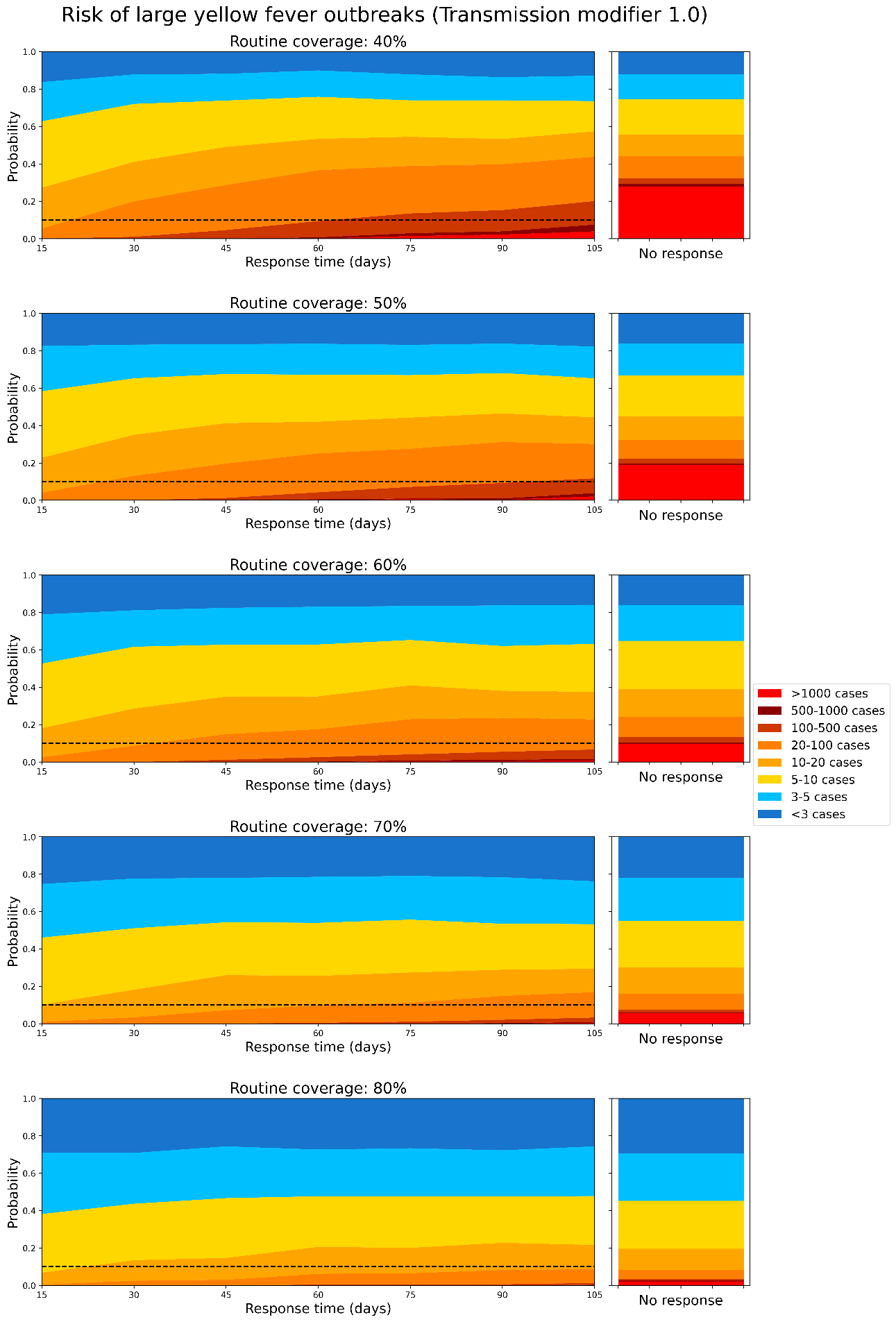

**Figure S21**: Area plot representing how the proportion of yellow fever outbreak simulations which exceed certain case thresholds changes as a function of ORI response time, assuming a routine vaccine coverage of: a)40%, b)50%, c)60%, d)70%, and e)80%, before the outbreak is seeded, and a transmission modifier of 1·0 for the mosquito vectors. The dashed line represents a 10% threshold. The righthand stacked bar represents the distribution of outbreak sizes if no ORI is delivered, for reference.

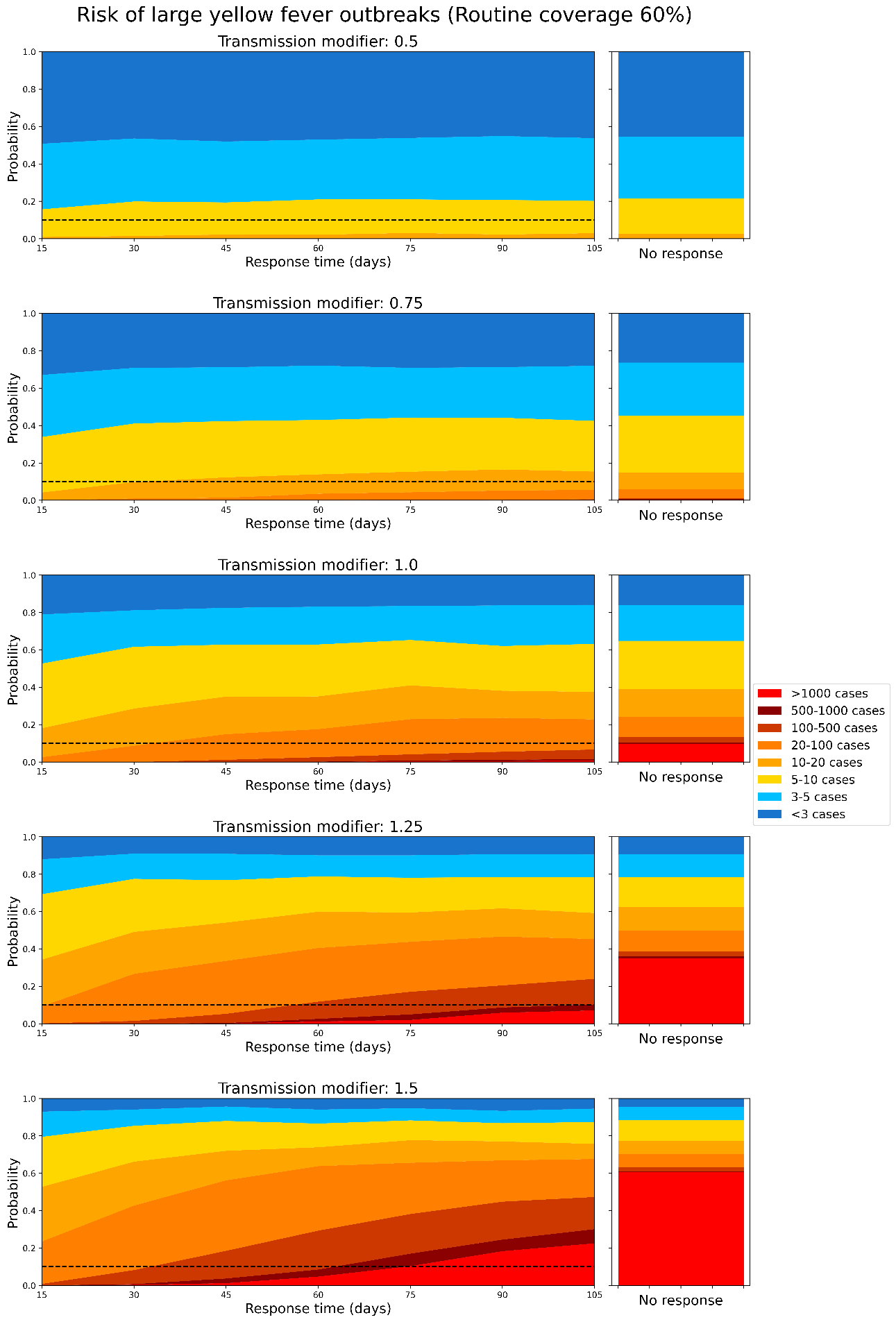

**Figure S22**: Area plot representing how the proportion of yellow fever outbreak simulations which exceed certain case thresholds changes as a function of ORI response time, assuming a transmission modifier for the mosquito vectors of: a)0·5, b)0·75, c)1·0, d)1·25, and e)1·5 before the outbreak is seeded, and a routine vaccine coverage of 60%. The dashed line represents a 10% threshold. The righthand stacked bar represents the distribution of outbreak sizes if no ORI is delivered, for reference.

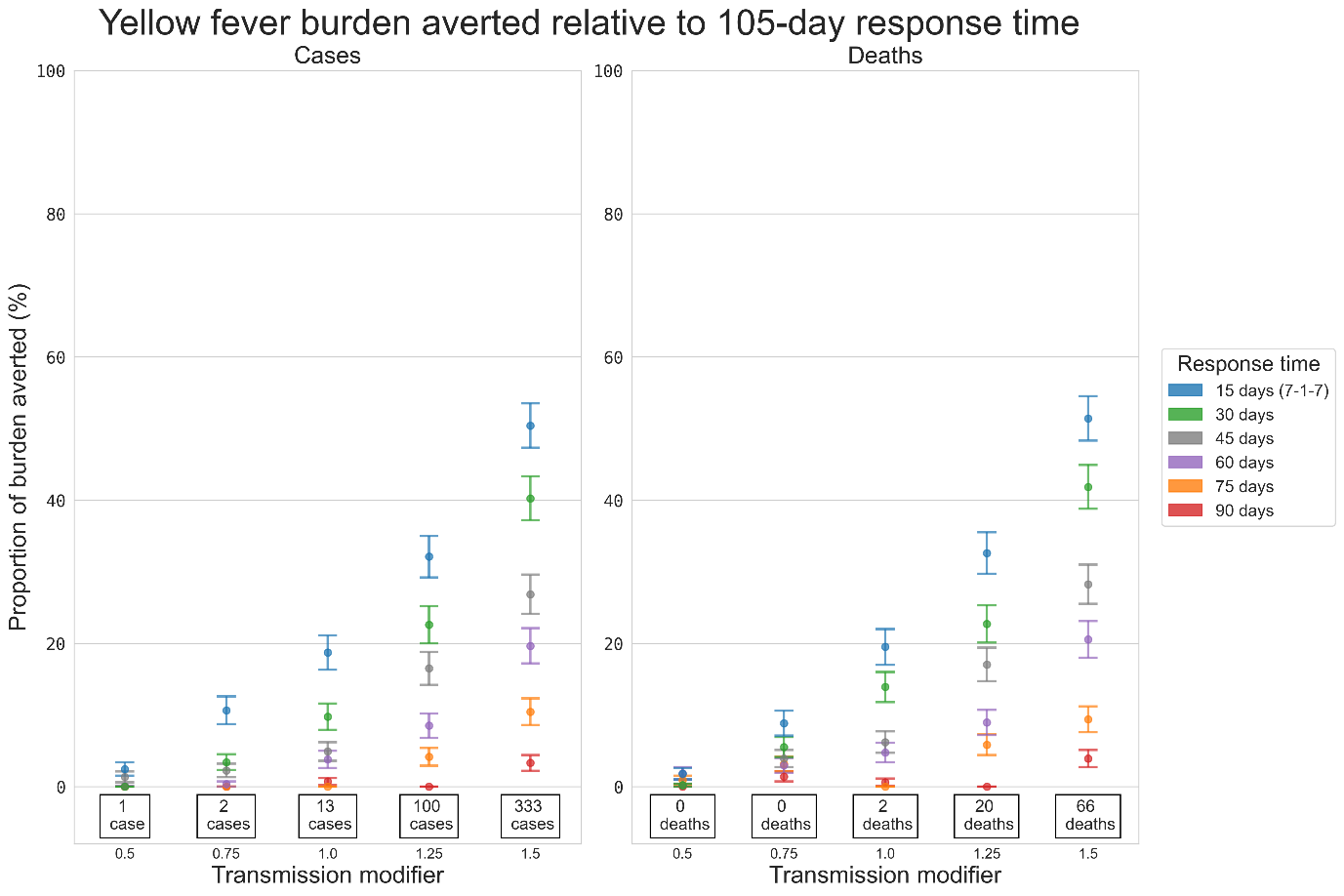

**Figure S23**: Proportional reduction in the mean cumulative cases and deaths for the set of yellow fever outbreaks with response times of 15, 30, 45, 60, 75, and 90 days relative to the set of outbreaks with a response time of 105 days, estimated using bootstrap resampling and assuming a routine vaccine coverage of 60%. Proportional impacts are grouped vertically by transmission modifier, and the values stated at the bottom of each column are the mean cumulative cases or deaths produced by outbreaks which received ORI after 105 days, i.e., the value serving as the denominator for the proportional reduction estimate.

**Table S5**: Proportional reduction in yellow fever cases, deaths, and DALYs relative to 105-day response, for each choice of routine vaccine coverage, assuming a mosquito-human transmission modifier of 1·0.

| Response time (days) | Routine coverage | Cases | Deaths | DALYs |
| --- | --- | --- | --- | --- |
| 90 | 40% | 0.045 (0.032, 0.058) | 0.04 (0.028, 0.052) | 0.032 (0.021, 0.043) |
| 75 |  | 0.07 (0.054, 0.086) | 0.064 (0.049, 0.079) | 0.06 (0.045, 0.074) |
| 60 |  | 0.085 (0.068, 0.102) | 0.081 (0.064, 0.097) | 0.075 (0.058, 0.091) |
| 45 |  | 0.164 (0.141, 0.187) | 0.165 (0.142, 0.188) | 0.161 (0.138, 0.184) |
| 30 |  | 0.233 (0.207, 0.259) | 0.233 (0.207, 0.259) | 0.227 (0.201, 0.253) |
| 15 |  | 0.349 (0.319, 0.378) | 0.334 (0.304, 0.363) | 0.343 (0.313, 0.372) |
| 90 | 50% | 0 (0.0, 0.0) | 0 (0.0, 0.0) | 0 (0.0, 0.0) |
| 75 |  | 0.031 (0.02, 0.042) | 0.049 (0.035, 0.062) | 0.033 (0.022, 0.044) |
| 60 |  | 0.062 (0.047, 0.077) | 0.074 (0.058, 0.09) | 0.068 (0.052, 0.083) |
| 45 |  | 0.092 (0.074, 0.11) | 0.119 (0.099, 0.139) | 0.098 (0.079, 0.116) |
| 30 |  | 0.157 (0.134, 0.179) | 0.168 (0.145, 0.192) | 0.153 (0.13, 0.175) |
| 15 |  | 0.239 (0.212, 0.265) | 0.242 (0.215, 0.268) | 0.247 (0.22, 0.273) |
| 90 | 60% | 0.007 (0.002, 0.012) | 0.006 (0.001, 0.011) | 0.006 (0.001, 0.011) |
| 75 |  | 0 (0.0, 0.0) | 0 (0.0, 0.0) | 0 (0.0, 0.0) |
| 60 |  | 0.038 (0.026, 0.05) | 0.048 (0.034, 0.061) | 0.026 (0.016, 0.036) |
| 45 |  | 0.049 (0.036, 0.062) | 0.062 (0.047, 0.077) | 0.047 (0.034, 0.06) |
| 30 |  | 0.098 (0.079, 0.116) | 0.139 (0.118, 0.16) | 0.109 (0.09, 0.128) |
| 15 |  | 0.187 (0.163, 0.211) | 0.195 (0.17, 0.22) | 0.197 (0.172, 0.221) |
| 90 | 70% | 0.02 (0.011, 0.028) | 0.029 (0.019, 0.039) | 0.025 (0.015, 0.035) |
| 75 |  | 0.012 (0.005, 0.018) | 0.039 (0.027, 0.05) | 0.016 (0.008, 0.024) |
| 60 |  | 0.042 (0.03, 0.055) | 0.069 (0.053, 0.084) | 0.054 (0.04, 0.068) |
| 45 |  | 0.048 (0.034, 0.061) | 0.068 (0.052, 0.083) | 0.055 (0.041, 0.069) |
| 30 |  | 0.096 (0.077, 0.114) | 0.117 (0.097, 0.137) | 0.108 (0.089, 0.127) |
| 15 |  | 0.152 (0.13, 0.174) | 0.202 (0.177, 0.226) | 0.155 (0.132, 0.177) |
| 90 | 80% | 0 (0.0, 0.0) | 0 (0.0, 0.0) | 0 (0.0, 0.0) |
| 75 |  | 0 (0.0, 0.0) | 0.01 (0.004, 0.016) | 0 (0.0, 0.0) |
| 60 |  | 0.014 (0.007, 0.022) | 0.031 (0.02, 0.042) | 0.02 (0.011, 0.028) |
| 45 |  | 0.039 (0.027, 0.051) | 0.05 (0.036, 0.063) | 0.041 (0.028, 0.053) |
| 30 |  | 0.058 (0.044, 0.073) | 0.058 (0.044, 0.073) | 0.06 (0.045, 0.074) |
| 15 |  | 0.112 (0.092, 0.131) | 0.079 (0.062, 0.095) | 0.102 (0.083, 0.12) |

**Table S6**: Proportional reduction in yellow fever cases, deaths, and DALYs relative to 105-day response, for each choice of mosquito-human transmission modifier, assuming a routine vaccine coverage of 60%.

| Response time (days) | Transmission modifier | Cases | Deaths | DALYs |
| --- | --- | --- | --- | --- |
| 90 | 0·5 | 0 (0.0, 0.0) | 0 (0.0, 0.0) | 0 (0.0, 0.0) |
| 75 |  | 0 (0.0, 0.0) | 0.009 (0.003, 0.015) | 0 (0.0, 0.0) |
| 60 |  | 0.001 (0, 0.001) | 0.019 (0.01, 0.027) | 0.006 (0.001, 0.01) |
| 45 |  | 0.014 (0.006, 0.021) | 0.002 (0, 0.004) | 0.013 (0.006, 0.019) |
| 30 |  | 0 (0.0, 0.0) | 0.002 (0, 0.004) | 0 (0.0, 0.0) |
| 15 |  | 0.025 (0.015, 0.034) | 0.018 (0.01, 0.026) | 0.028 (0.018, 0.038) |
| 90 | 0·75 | 0 (0.0, 0.0) | 0.014 (0.007, 0.021) | 0.003 (0, 0.006) |
| 75 |  | 0 (0.0, 0.0) | 0.032 (0.021, 0.042) | 0.001 (0, 0.001) |
| 60 |  | 0.004 (0, 0.007) | 0.03 (0.019, 0.04) | 0.012 (0.005, 0.018) |
| 45 |  | 0.023 (0.013, 0.032) | 0.039 (0.027, 0.051) | 0.028 (0.018, 0.039) |
| 30 |  | 0.034 (0.023, 0.045) | 0.055 (0.041, 0.069) | 0.042 (0.029, 0.054) |
| 15 |  | 0.107 (0.087, 0.126) | 0.089 (0.071, 0.106) | 0.114 (0.094, 0.133) |
| 90 | 1·0 | 0.007 (0.002, 0.012) | 0.006 (0.001, 0.011) | 0.006 (0.001, 0.011) |
| 75 |  | 0 (0.0, 0.0) | 0 (0.0, 0.0) | 0 (0.0, 0.0) |
| 60 |  | 0.038 (0.026, 0.05) | 0.048 (0.034, 0.061) | 0.026 (0.016, 0.036) |
| 45 |  | 0.049 (0.036, 0.062) | 0.062 (0.047, 0.077) | 0.047 (0.034, 0.06) |
| 30 |  | 0.098 (0.079, 0.116) | 0.139 (0.118, 0.16) | 0.109 (0.09, 0.128) |
| 15 |  | 0.187 (0.163, 0.211) | 0.195 (0.17, 0.22) | 0.197 (0.172, 0.221) |
| 90 | 1·25 | 0 (0.0, 0.0) | 0 (0.0, 0.0) | 0 (0.0, 0.0) |
| 75 |  | 0.042 (0.029, 0.054) | 0.058 (0.044, 0.073) | 0.038 (0.026, 0.049) |
| 60 |  | 0.085 (0.068, 0.102) | 0.09 (0.072, 0.107) | 0.081 (0.064, 0.098) |
| 45 |  | 0.165 (0.142, 0.188) | 0.17 (0.147, 0.194) | 0.161 (0.138, 0.184) |
| 30 |  | 0.226 (0.2, 0.252) | 0.227 (0.201, 0.253) | 0.22 (0.194, 0.245) |
| 15 |  | 0.321 (0.292, 0.35) | 0.326 (0.297, 0.355) | 0.315 (0.286, 0.344) |
| 90 | 1·5 | 0.033 (0.022, 0.044) | 0.039 (0.027, 0.051) | 0.042 (0.03, 0.055) |
| 75 |  | 0.105 (0.086, 0.123) | 0.094 (0.076, 0.112) | 0.091 (0.073, 0.109) |
| 60 |  | 0.197 (0.172, 0.221) | 0.206 (0.18, 0.231) | 0.198 (0.173, 0.222) |
| 45 |  | 0.268 (0.241, 0.296) | 0.283 (0.255, 0.31) | 0.261 (0.233, 0.288) |
| 30 |  | 0.402 (0.372, 0.433) | 0.419 (0.388, 0.449) | 0.404 (0.374, 0.435) |
| 15 |  | 0.504 (0.473, 0.535) | 0.079 (0.062, 0.095) | 0.493 (0.462, 0.524) |

When the routine vaccine coverage was fixed at 60%, in a medium-low (0·75) transmission modifier setting, a 15-day response time was estimated to avert ~10% of the burden, while a 90-day response time averted almost no additional burden. However, in a medium-high (1·25) transmission modifier setting, a 15-day response time was estimated to avert ~32% of the burden, while a 90-day response time averted almost no additional burden compared to a 105-day response time (Figure S23 and Table S5).

#### 5.8 Limitations

- Uncertain case fatality rate: The model relies on 20% based on the observed mortality rate in the dataset. However, literature reports a mortality rate of 39%[63]. If the case fatality rate is greater than 20%, then the model may have underestimated deaths and vaccine impact.
- Uncertain routine vaccine coverage: WUENIC data used for estimating the underlying immunity describes the percentage of surviving infants who received one dose of yellow fever vaccine in countries where yellow fever is part of the immunisation schedule for children or is recommended in at-risk areas[58]. Additionally, there is uncertainty around the vaccine coverage due to over-estimation of yellow fever vaccination reporting, which is accounted for in the model. However, there is high uncertainty around sensitivity and specificity of reporting.
- Uncertainty around re-vaccination: Re-vaccination of a proportion of the target population is expected as prior determination of the vaccination status is challenging in LMICs. The model incorporates re-vaccination of agents; however, there is no change in immunity. The results might therefore underestimate the impact of the ORI.
- Climate covariates: Mosquito population is controlled by rainfall and temperature (e.g., dry and wet seasons, hot and cold periods). The model assumes urban outbreaks without any initial spillover force of infection.
- Modelling mild vs. severe: The model is not disaggregating asymptomatic vs. symptomatic because little data is available to inform differences in disease duration and infectiousness, and because testing is driven by severe vs. non-severe.
- Uncertainty around the shapes of the epidemic curves for the yellow fever outbreaks which were included in the calibration, due to a lack of available data. We were unable to calibrate the model against detailed timeseries data for the outbreaks included in the calibration dataset, only the cumulative cases and outbreaks duration. As such, the model may not accurately reflect the impact of the ORI response time if it is over or underestimating the number of cases which have occurred during an outbreak before ORI programs can be implemented.
- Uncertainty around the expected coverage achieved by ORI during an outbreak: It is unclear what level of population coverage is typically achieved by ORI campaigns, and it is likely that it would vary widely based on the outbreak context. Our assumption of 75% achieved coverage would be optimistic if outbreaks occur in remote or otherwise hard to reach populations, and would be pessimistic if the population at-risk has a strong history of vaccine uptake. We attempt to understand the impact of this assumption in our *Alternate coverage* scenario.

### 6 Sensitivity analyses

#### 6.1 Alternate coverage

In this section we present the proportional impact of varying the assumption for the level of coverage achieved by ORI for outbreaks of meningococcal meningitis, cholera, measles, and yellow fever, disaggregated by ORI response time and setting archetype. The results of the alternate ORI coverage sensitivity analysis can be seen in Figures S24 – S40.

##### 6.1.1 Meningococcal meningitis

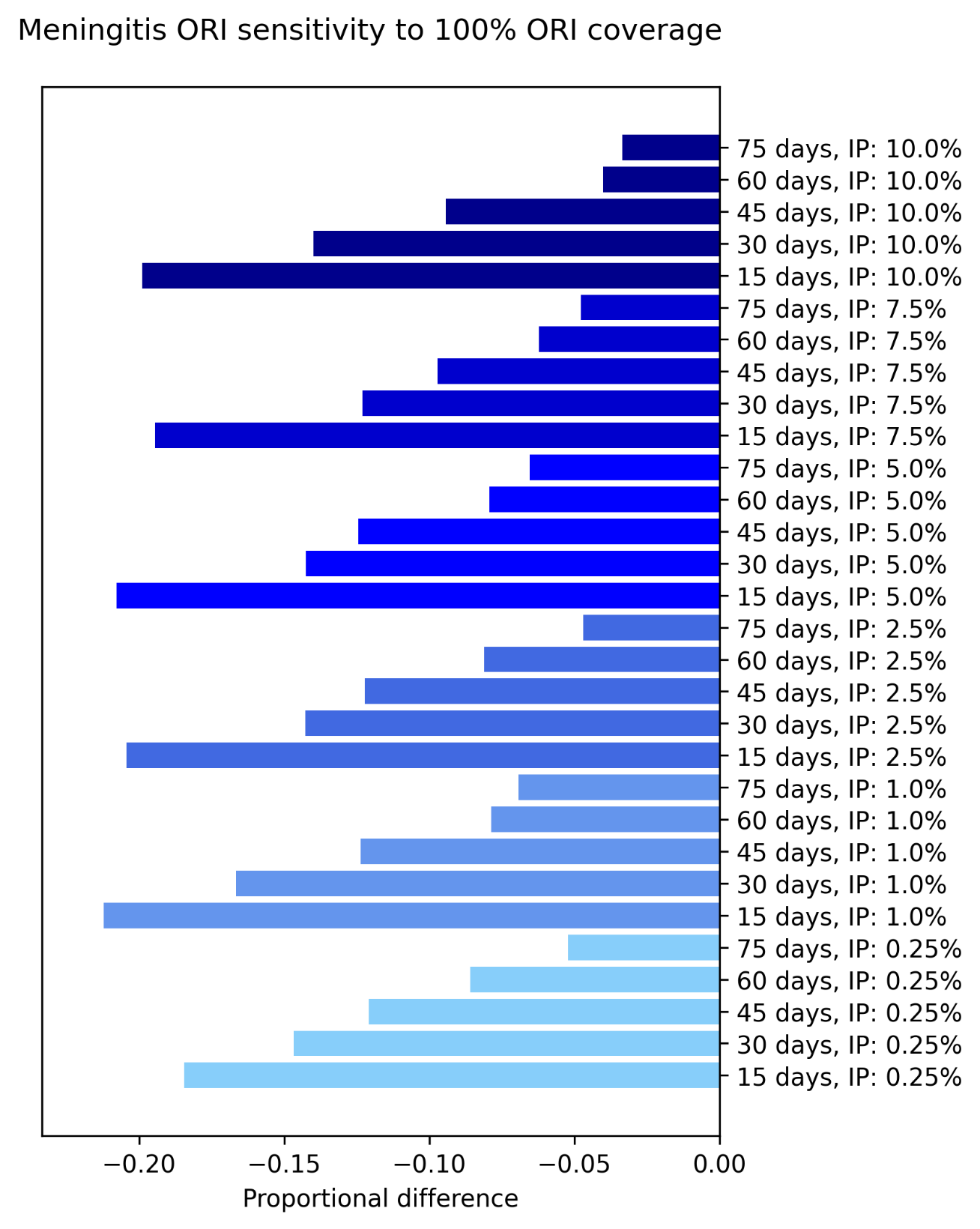

**Figure S24**: Proportional reduction in the mean cumulative cases and deaths for the set of outbreaks which received ORI that reached 100% of people aged 1 – 29 years, relative to the set of outbreaks which received ORI that reached 75% of people aged 1 – 29 years, estimated using bootstrap resampling. Proportional impacts are grouped by initial asymptomatic carriage prevalence (IP) and ordered by response time.

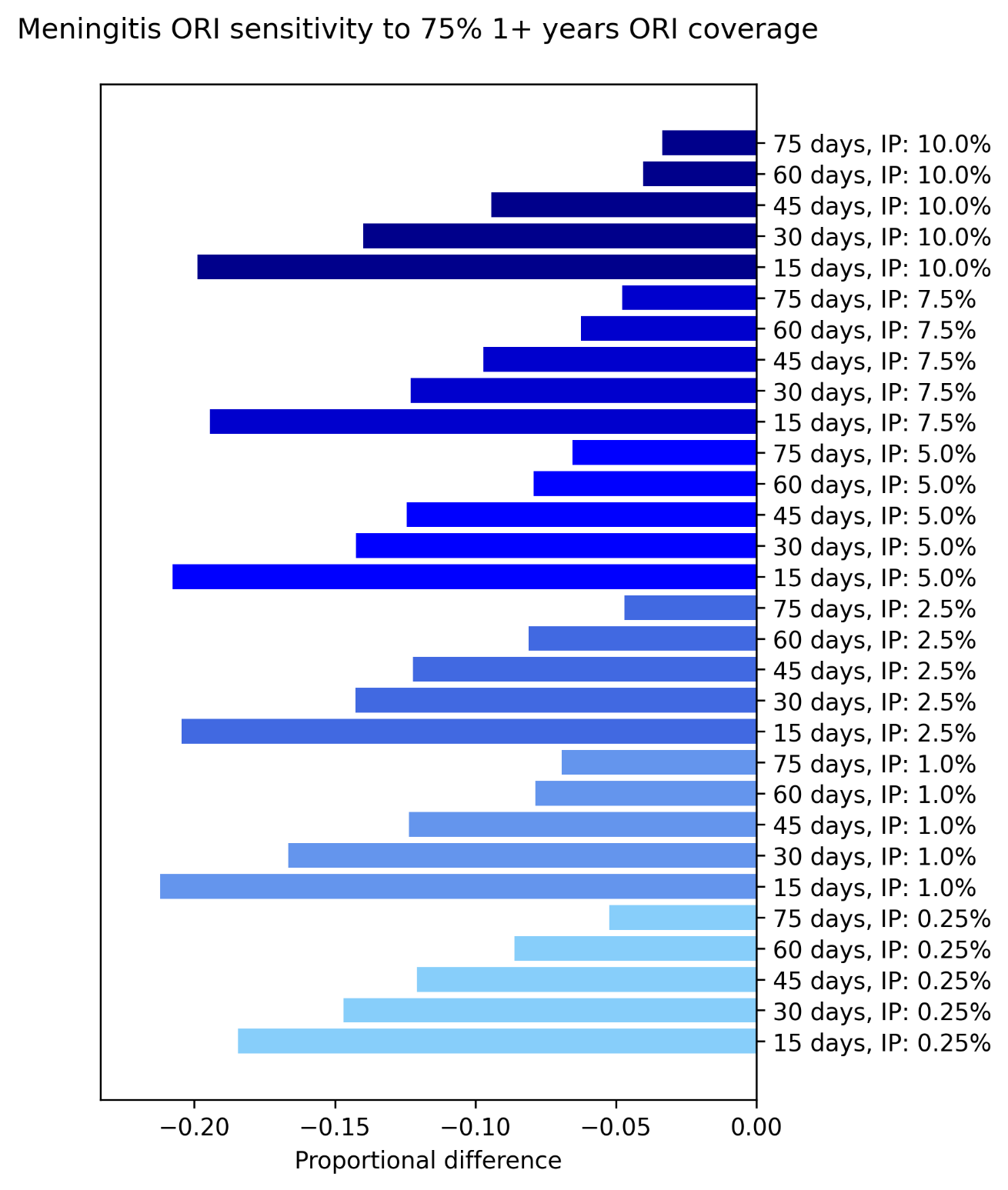

**Figure S25**: Proportional reduction in the mean cumulative cases and deaths for the set of outbreaks which received ORI that reached 75% of people aged 1+ years, relative to the set of outbreaks which received ORI that reached 75% of people aged 1 – 29 years, estimated using bootstrap resampling. Proportional impacts are grouped by initial asymptomatic carriage prevalence (IP) and ordered by response time.

Increasing achieved ORI coverage for meningitis outbreaks from 75% to 100% in people aged 1 – 29 years, or increasing the age-group targeted to people over one year of age (but maintaining 75% coverage) produced similar proportional impacts. Both scenarios decreased the observed cumulative cases by approximately 20 – 25% at most, for 15-day ORI response times. The impacts were greater for faster ORI response times. Overall, increasing the range of the age group targeted by the vaccines had a larger impact than increasing the coverage within 1 – 29 years, as immunising 75% of people aged over 30 years likely had a larger protective effect than reaching a further 25% of people aged 1 – 29 years despite the older age group’s lower susceptibility to infection and invasive disease.

##### 6.1.2 Cholera

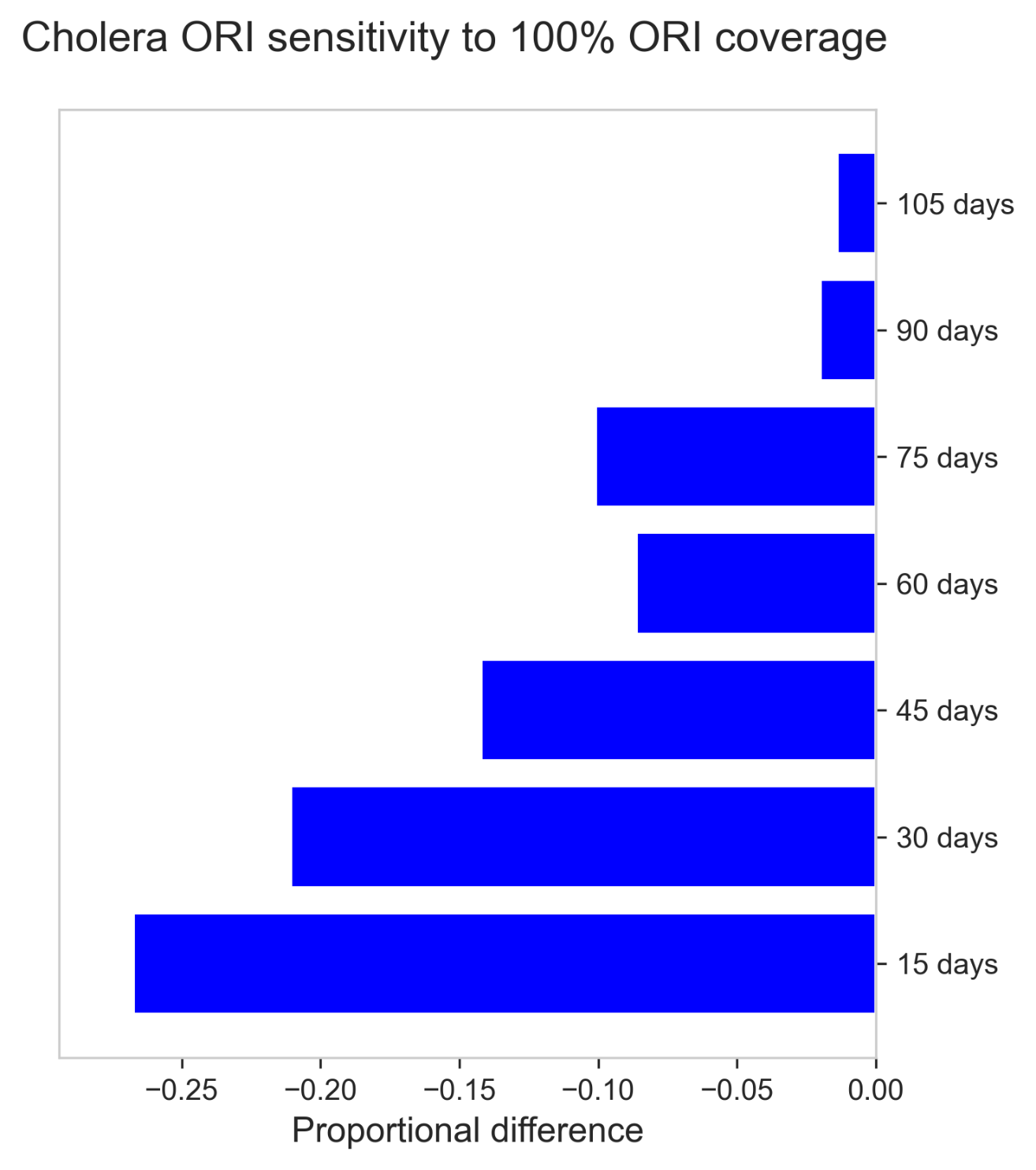

**Figure S26**: Proportional reduction in the mean cumulative cases and deaths for the set of outbreaks which received ORI that reached 100% of people, relative to the set of outbreaks which received ORI that reached 75% of people, estimated using bootstrap resampling.

Increasing the achieved ORI coverage for cholera outbreaks from 75% to 100% in the target population was estimated to decrease the expected number of cases observed by up to 27%, with impacts increasing as ORI response time decreased.

##### 6.1.3 Measles

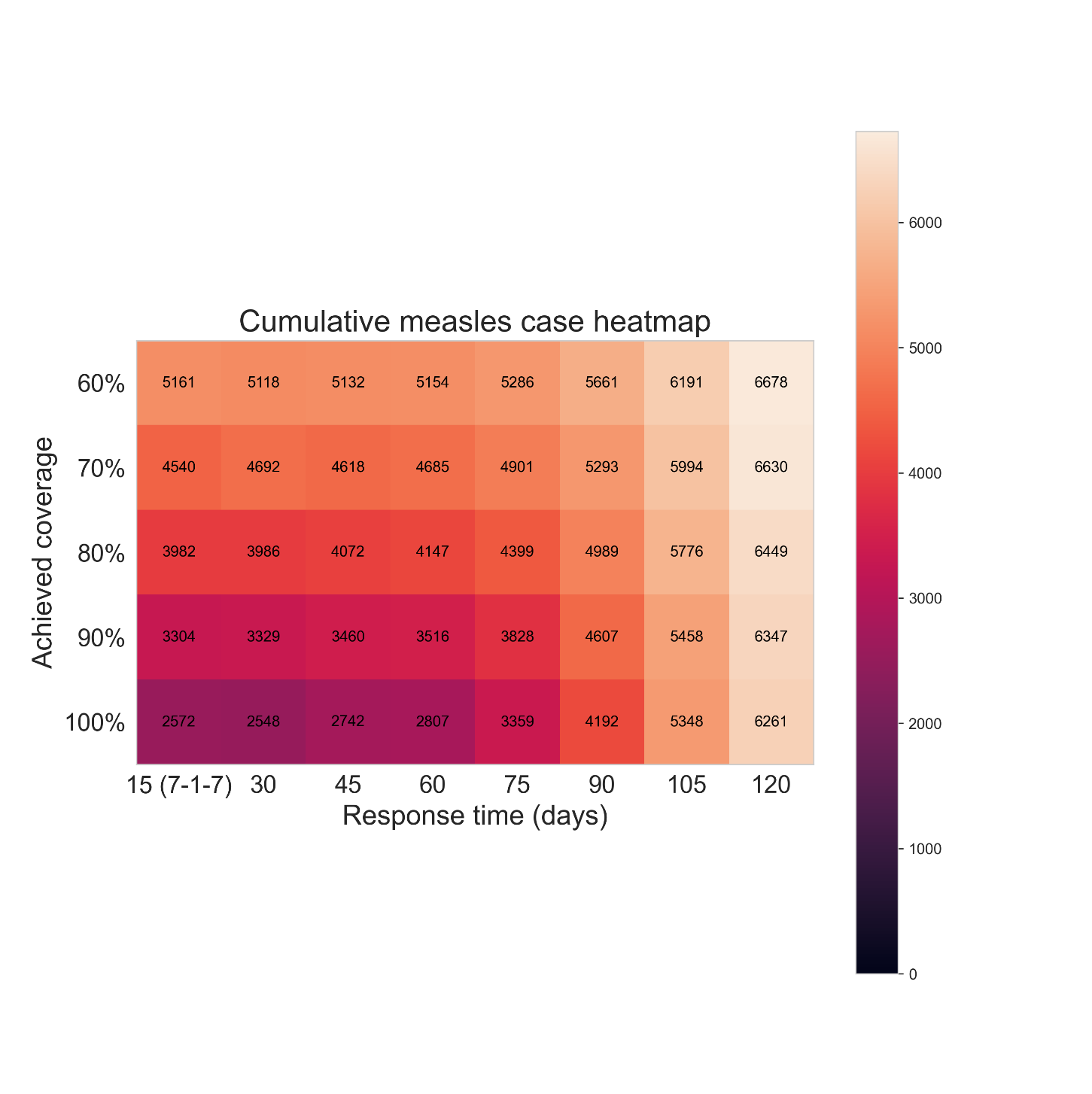

**Figure S27**: Heatmap of the mean cumulative cases by 1000 measles outbreak simulations, given different ORI response times (15, 30, 45, 60, 75, 90, 105, or 120 days) and ORI coverages achieved by the response (60%, 70%, 80%, 90% or 100%), assuming a routine vaccine coverage of 50%. For each achieved coverage, outbreaks which received faster responses of ORI tended to produce fewer cumulative cases, with the effect becoming more noticeable at higher achieved coverages.

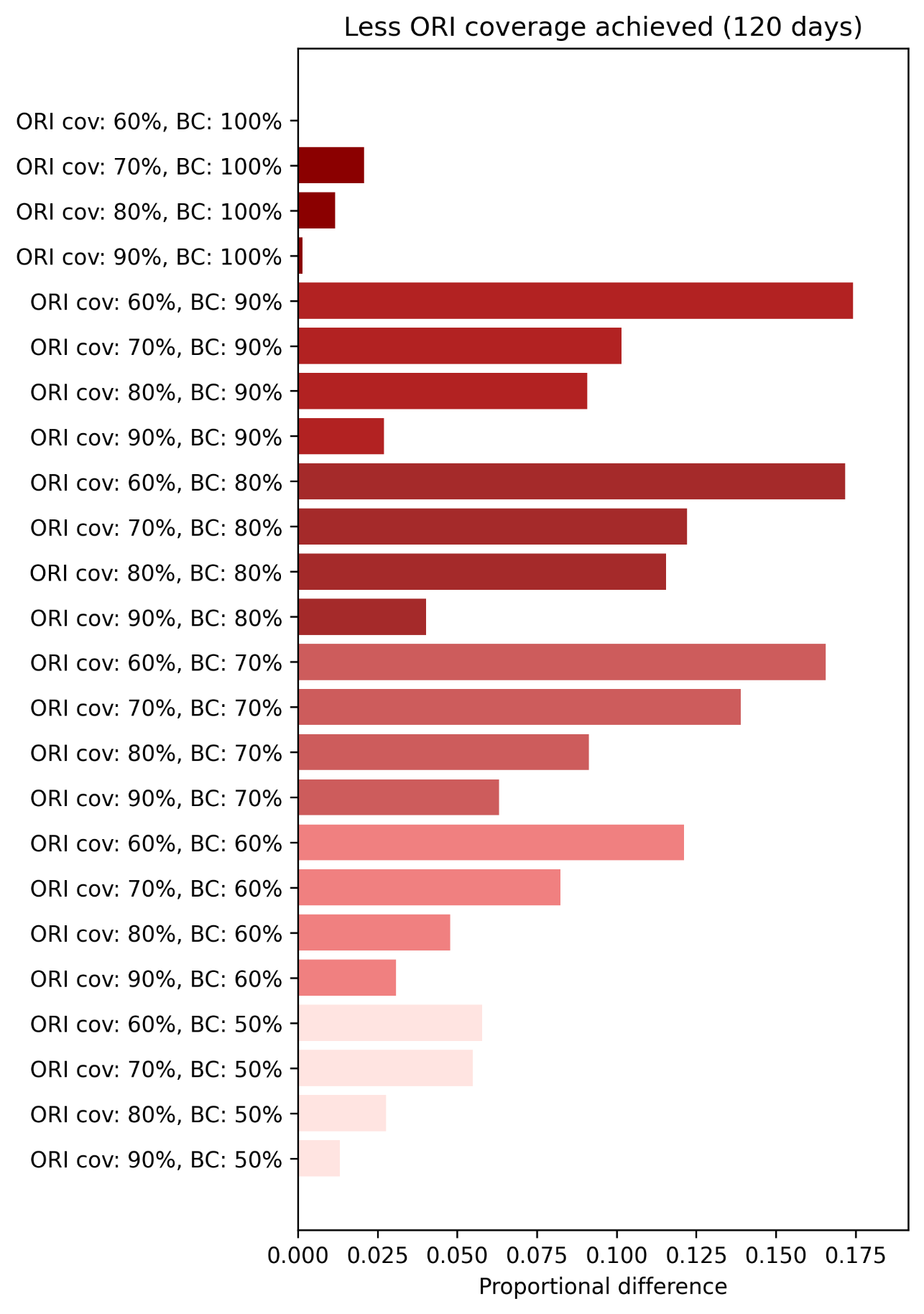

**Figure S28**: Proportional increase in the mean cumulative cases and deaths for the set of outbreaks with achieved ORI coverages of 60% ,70%, 80%, and 90%, grouped by routine vaccine coverage level in the population (BC), relative to the set of outbreaks with an achieved coverage of 100%, estimated using bootstrap resampling and assuming an ORI response time of 120 days. Proportional impacts are grouped by routine vaccine coverage and ordered by achieved ORI coverage (ORI cov).

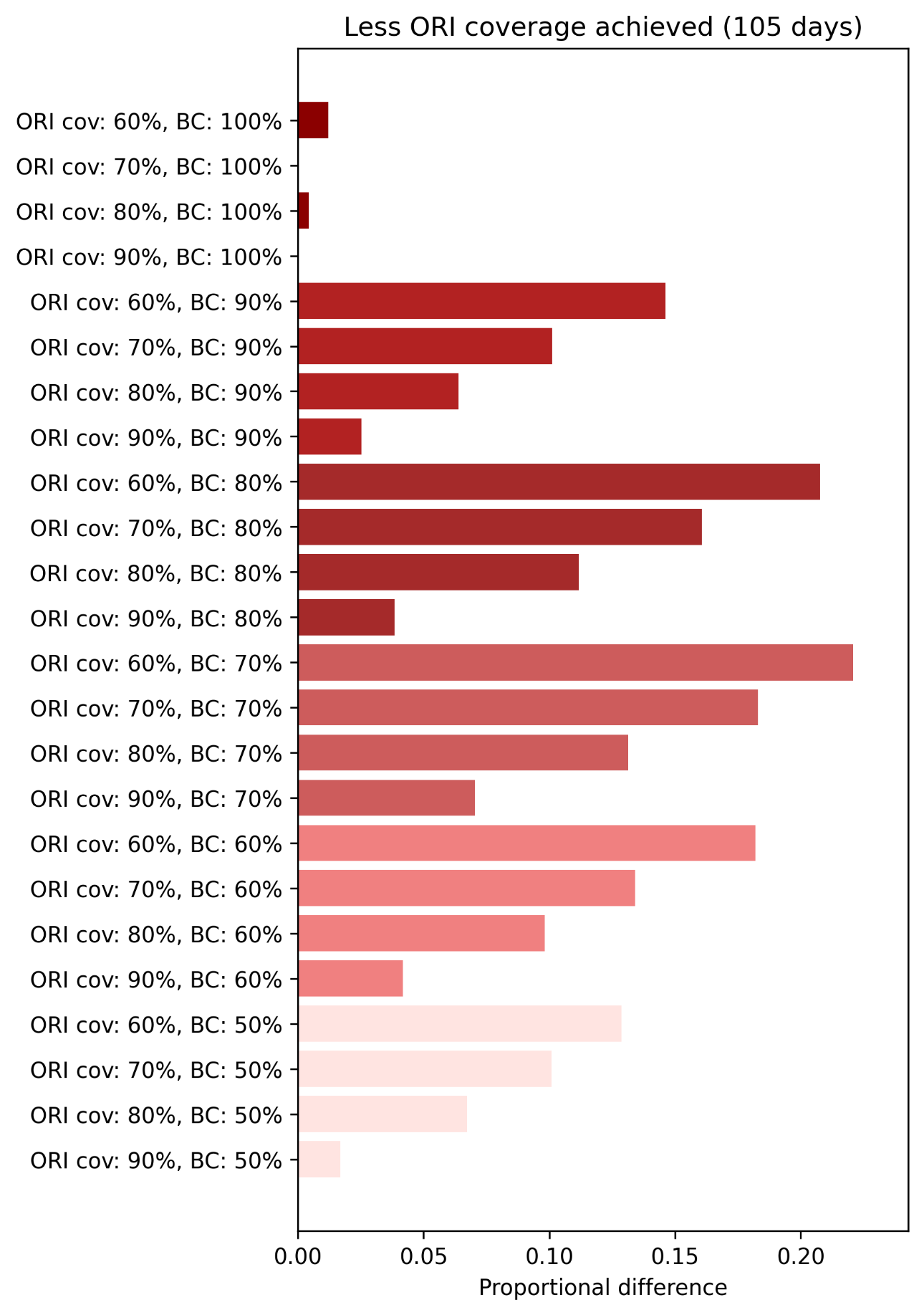

**Figure S29**: Proportional increase in the mean cumulative cases and deaths for the set of outbreaks with achieved ORI coverages of 60% ,70%, 80%, and 90%, grouped by routine vaccine coverage level in the population (BC), relative to the set of outbreaks with an achieved coverage of 100%, estimated using bootstrap resampling and assuming an ORI response time of 105 days. Proportional impacts are grouped by routine vaccine coverage and ordered by achieved ORI coverage (ORI cov).

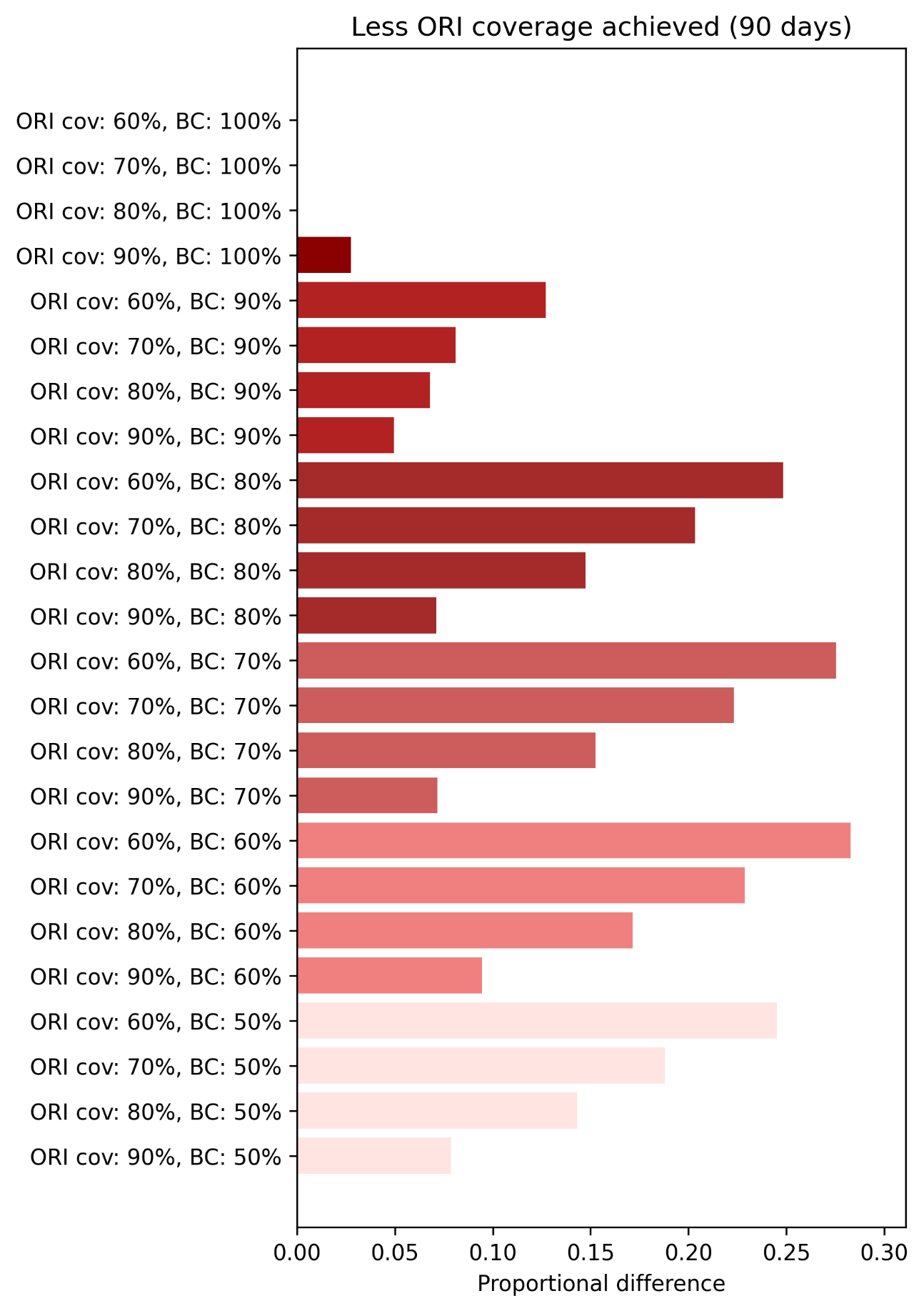

**Figure S30**: Proportional increase in the mean cumulative cases and deaths for the set of outbreaks with achieved ORI coverages of 60% ,70%, 80%, and 90%, grouped by routine vaccine coverage level in the population (BC), relative to the set of outbreaks with an achieved coverage of 100%, estimated using bootstrap resampling and assuming an ORI response time of 90 days. Proportional impacts are grouped by routine vaccine coverage and ordered by achieved ORI coverage (ORI cov).

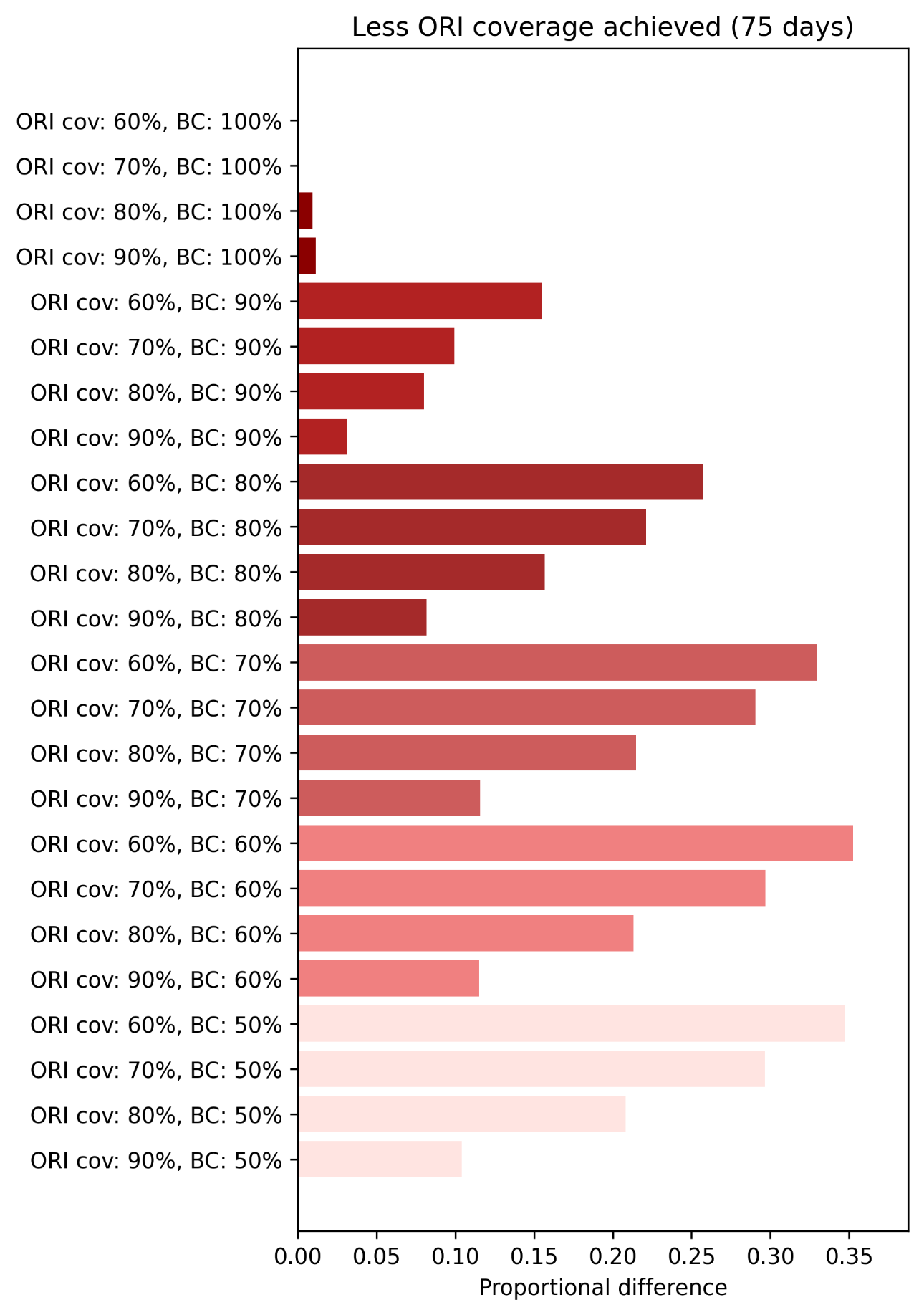

**Figure S31**: Proportional increase in the mean cumulative cases and deaths for the set of outbreaks with achieved ORI coverages of 60% ,70%, 80%, and 90%, grouped by routine vaccine coverage level in the population (BC), relative to the set of outbreaks with an achieved coverage of 100%, estimated using bootstrap resampling and assuming an ORI response time of 75 days. Proportional impacts are grouped by routine vaccine coverage and ordered by achieved ORI coverage (ORI cov).

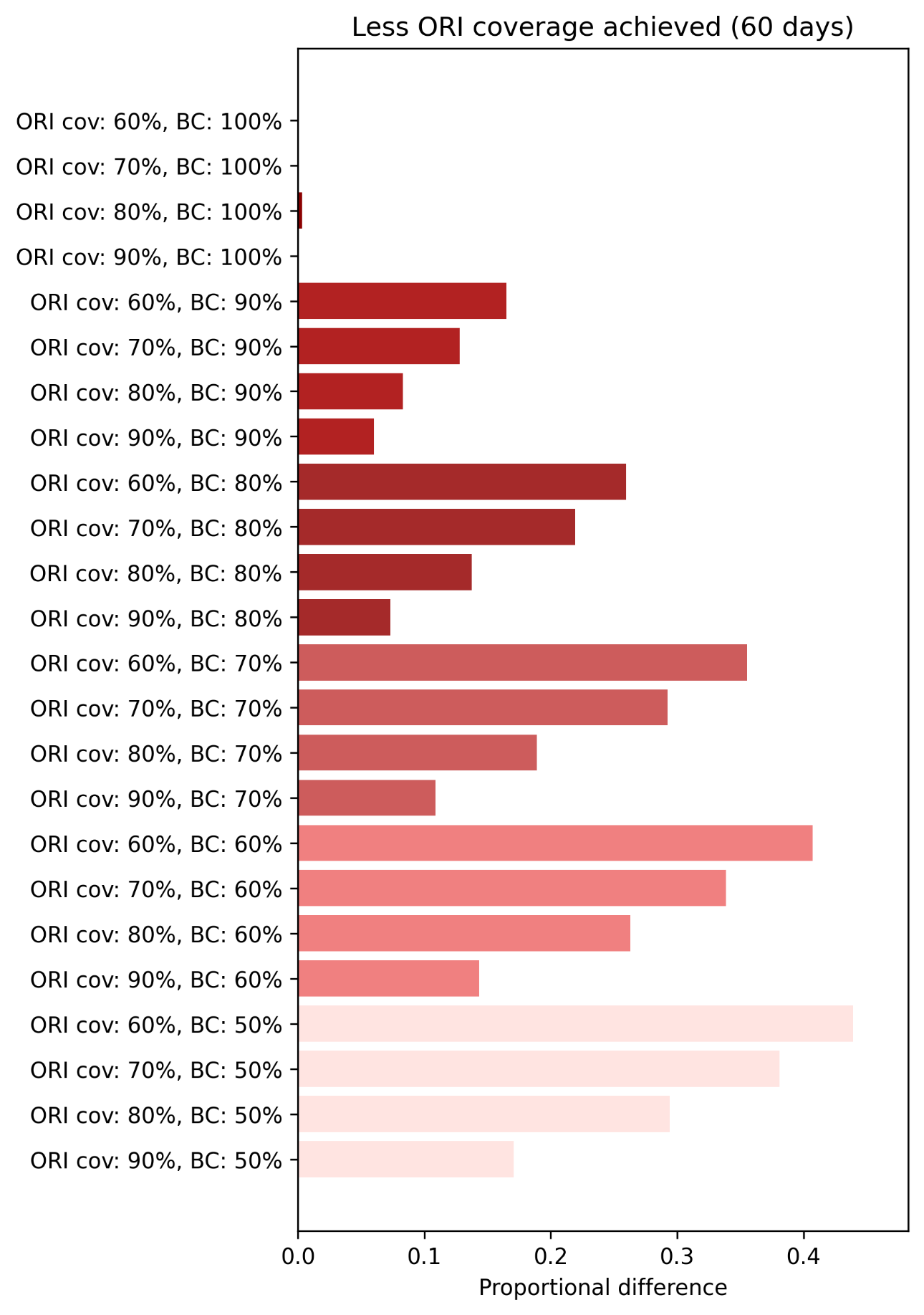

**Figure S32**: Proportional increase in the mean cumulative cases and deaths for the set of outbreaks with achieved ORI coverages of 60% ,70%, 80%, and 90%, grouped by routine vaccine coverage level in the population (BC), relative to the set of outbreaks with an achieved coverage of 100%, estimated using bootstrap resampling and assuming an ORI response time of 60 days. Proportional impacts are grouped by routine vaccine coverage and ordered by achieved ORI coverage (ORI cov).

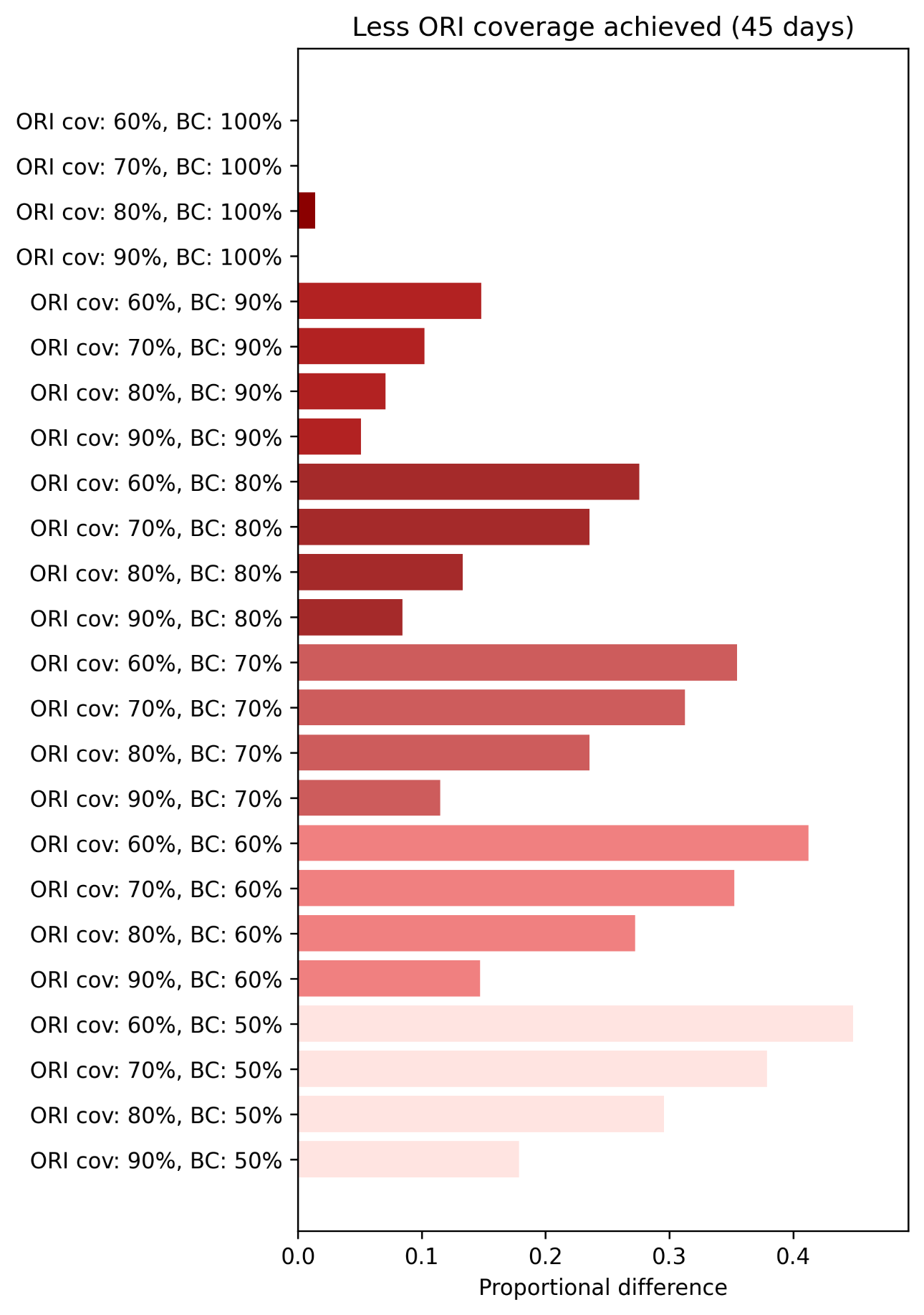

**Figure S33**: Proportional increase in the mean cumulative cases and deaths for the set of outbreaks with achieved ORI coverages of 60% ,70%, 80%, and 90%, grouped by routine vaccine coverage level in the population (BC), relative to the set of outbreaks with an achieved coverage of 100%, estimated using bootstrap resampling and assuming an ORI response time of 45 days. Proportional impacts are grouped by routine vaccine coverage and ordered by achieved ORI coverage (ORI cov).

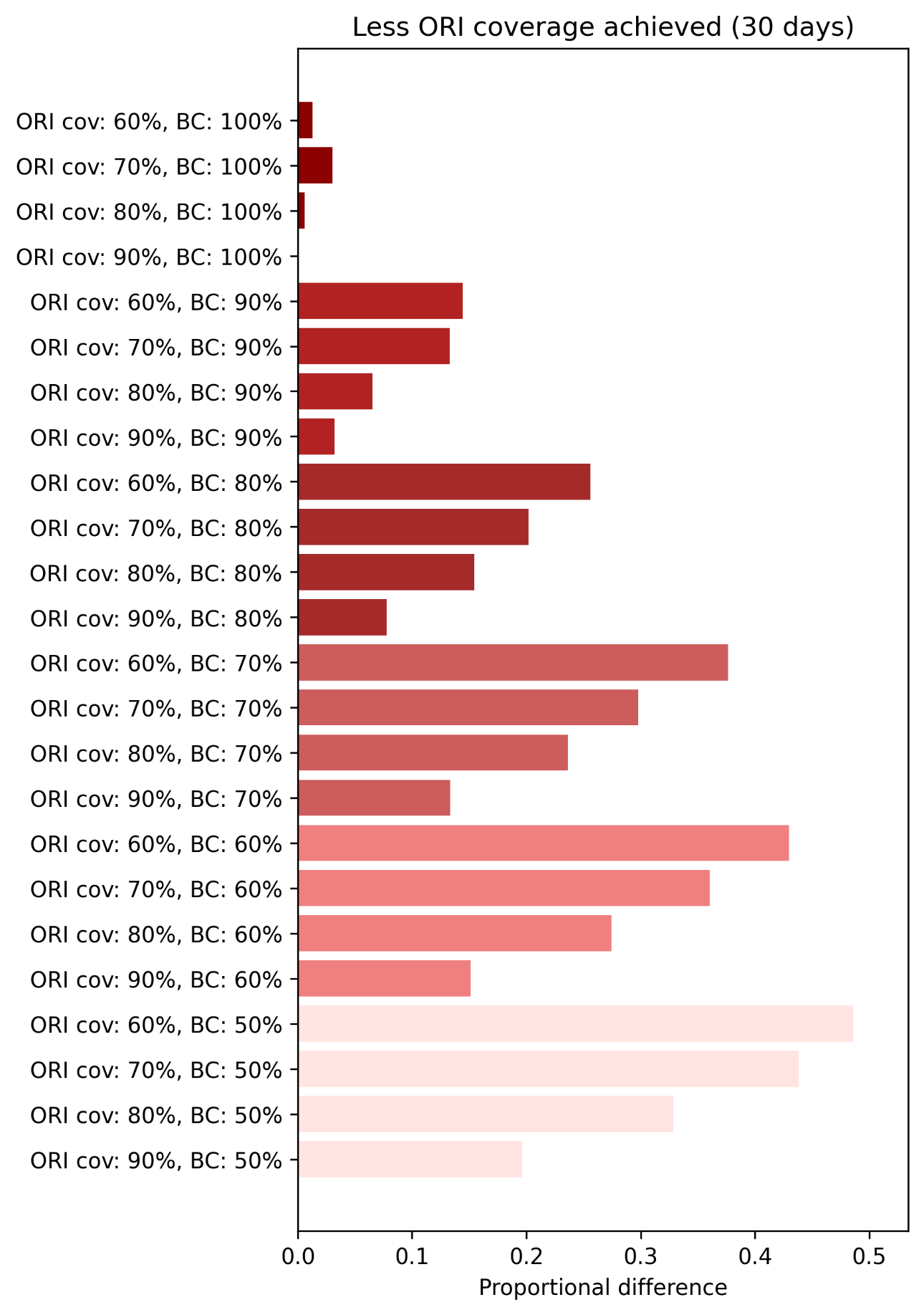

**Figure S34**: Proportional increase in the mean cumulative cases and deaths for the set of outbreaks with achieved ORI coverages of 60% ,70%, 80%, and 90%, grouped by routine vaccine coverage level in the population (BC), relative to the set of outbreaks with an achieved coverage of 100%, estimated using bootstrap resampling and assuming an ORI response time of 30 days. Proportional impacts are grouped by routine vaccine coverage and ordered by achieved ORI coverage (ORI cov).

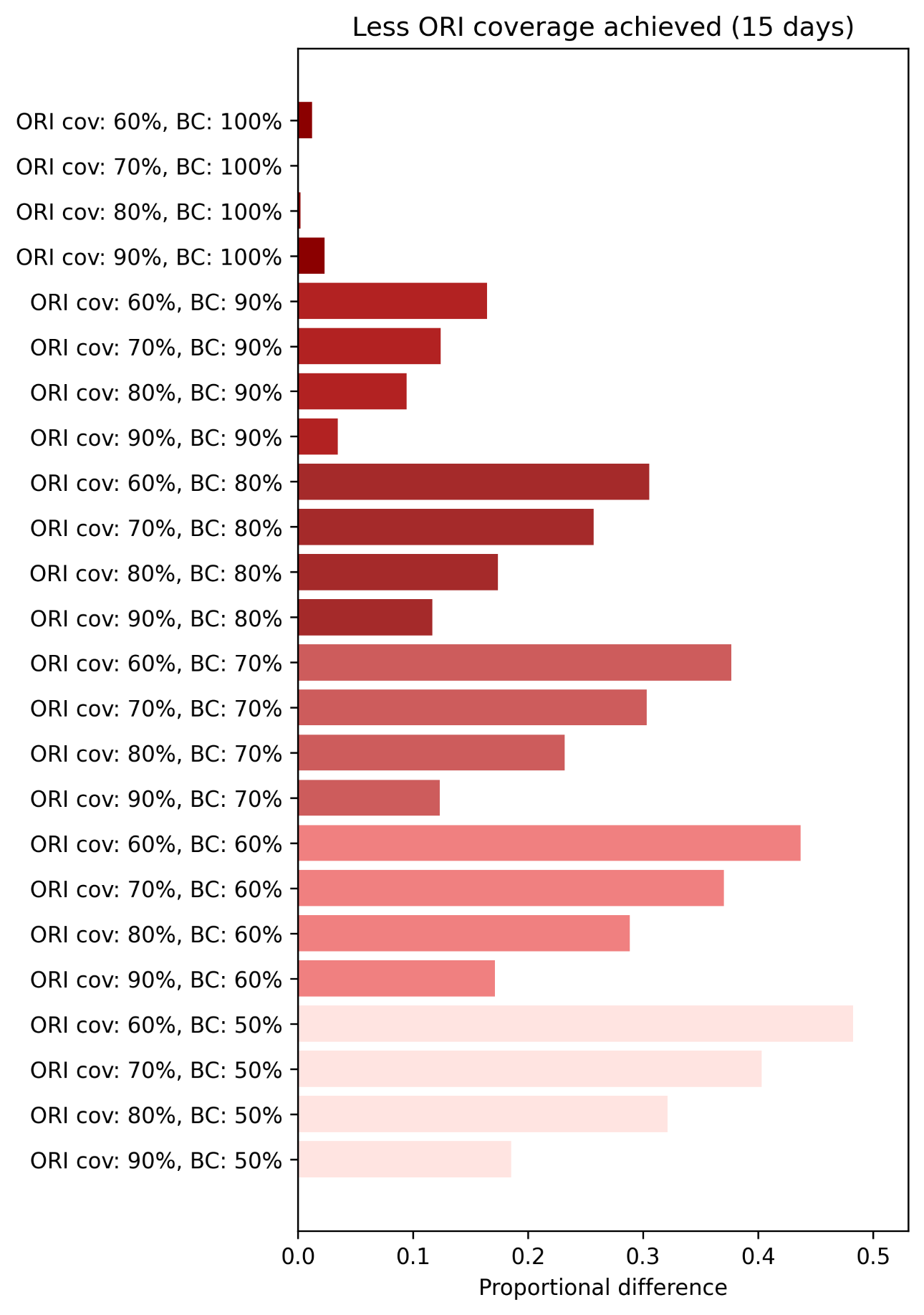

**Figure S35**: Proportional increase in the mean cumulative cases and deaths for the set of outbreaks with achieved ORI coverages of 60% ,70%, 80%, and 90%, grouped by routine vaccine coverage level in the population (BC), relative to the set of outbreaks with an achieved coverage of 100%, estimated using bootstrap resampling and assuming an ORI response time of 15 days (7-1-7). Proportional impacts are grouped by routine vaccine coverage and ordered by achieved ORI coverage (ORI cov).

##### 6.1.4 Yellow fever

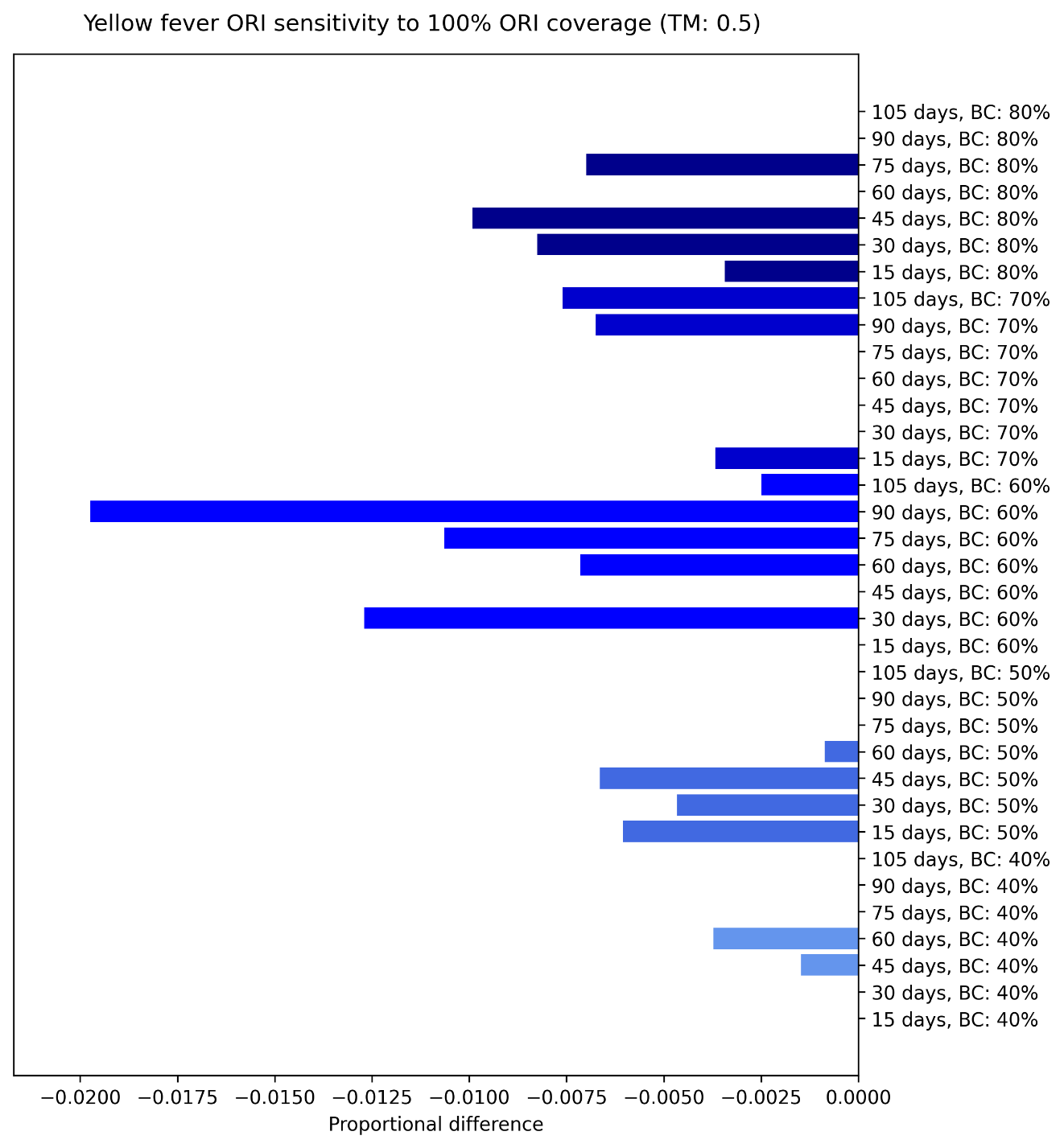

**Figure S36**: Proportional reduction in the mean cumulative cases and deaths for the set of outbreaks which received ORI that reached 100% of people, relative to the set of outbreaks which received ORI that reached 75% of people, estimated using bootstrap resampling and assuming a transmission modifier of 0·5 for the mosquito vectors. Proportional impacts are grouped vertically by routine vaccine coverage and ordered by ORI response time. There is no clear trend in the impacts as they are negligible and likely due to stochastic effects.

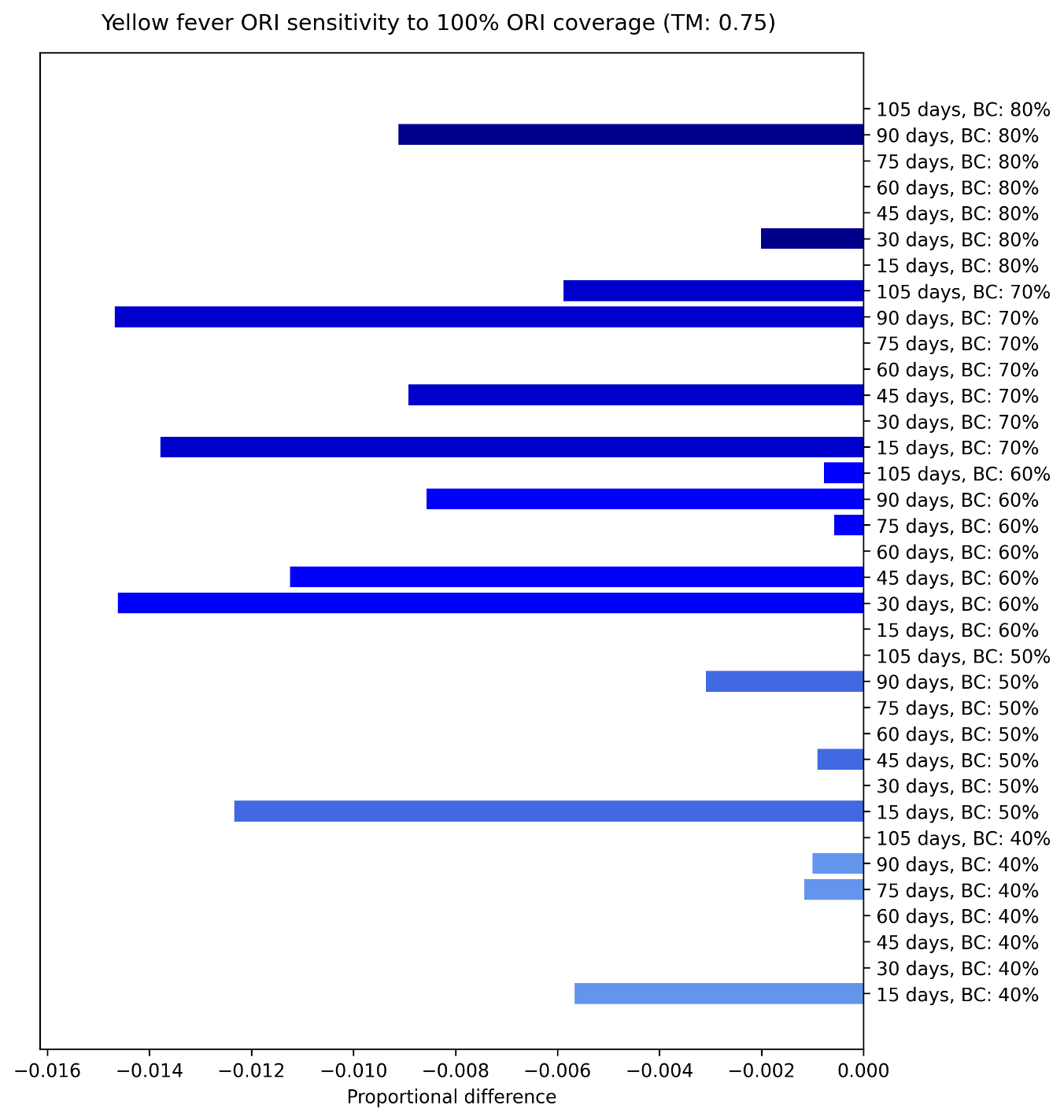

**Figure S37**: Proportional reduction in the mean cumulative cases and deaths for the set of outbreaks which received ORI that reached 100% of people, relative to the set of outbreaks which received ORI that reached 75% of people, estimated using bootstrap resampling and assuming a transmission modifier of 0·75 for the mosquito vectors. Proportional impacts are grouped vertically by routine vaccine coverage and ordered by ORI response time. There is no clear trend in the impacts as they are negligible and likely due to stochastic effects.

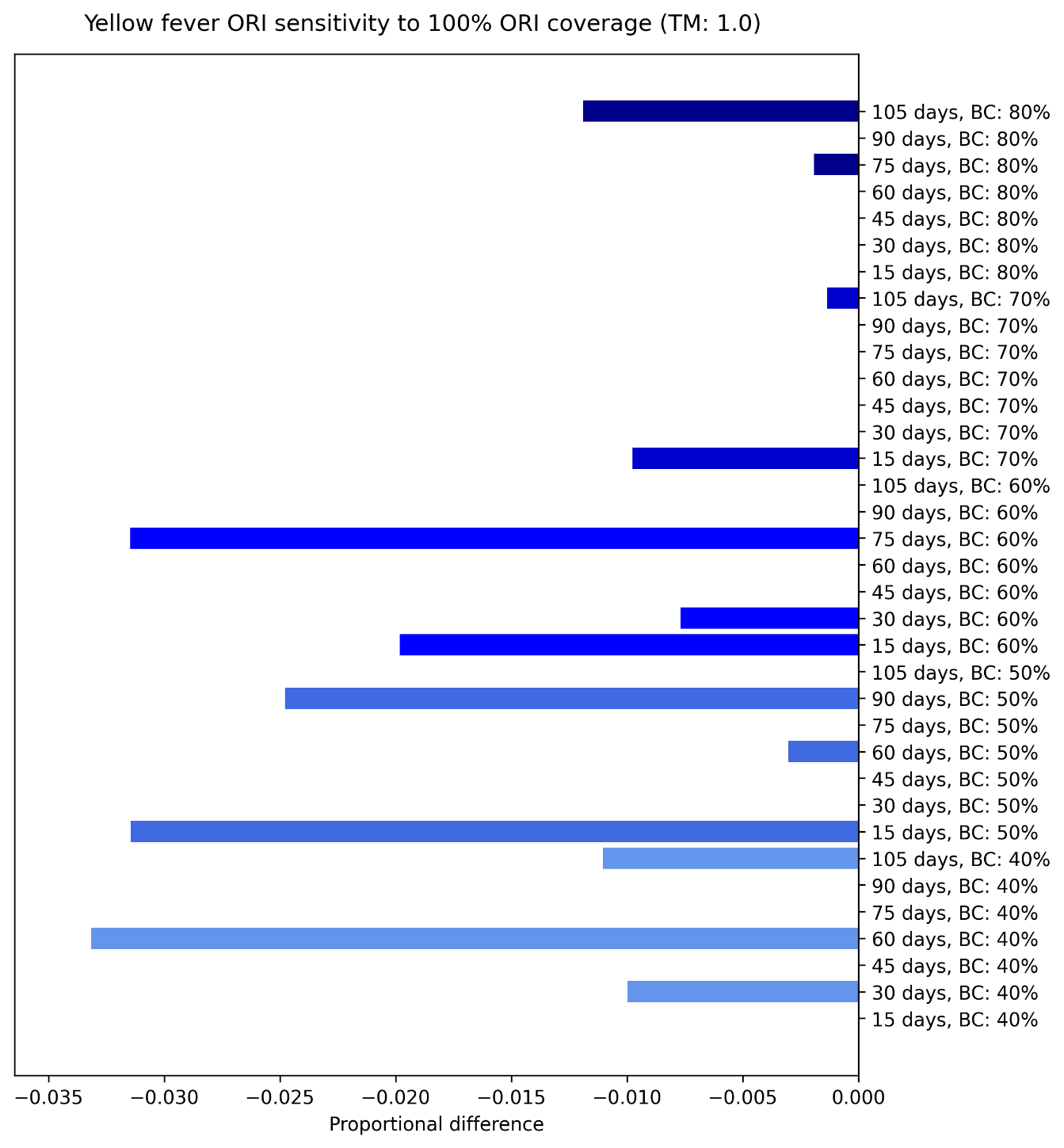

**Figure S38**: Proportional reduction in the mean cumulative cases and deaths for the set of outbreaks which received ORI that reached 100% of people, relative to the set of outbreaks which received ORI that reached 75% of people, estimated using bootstrap resampling and assuming a transmission modifier of 1·0 for the mosquito vectors. Proportional impacts are grouped vertically by routine vaccine coverage and ordered by ORI response time.

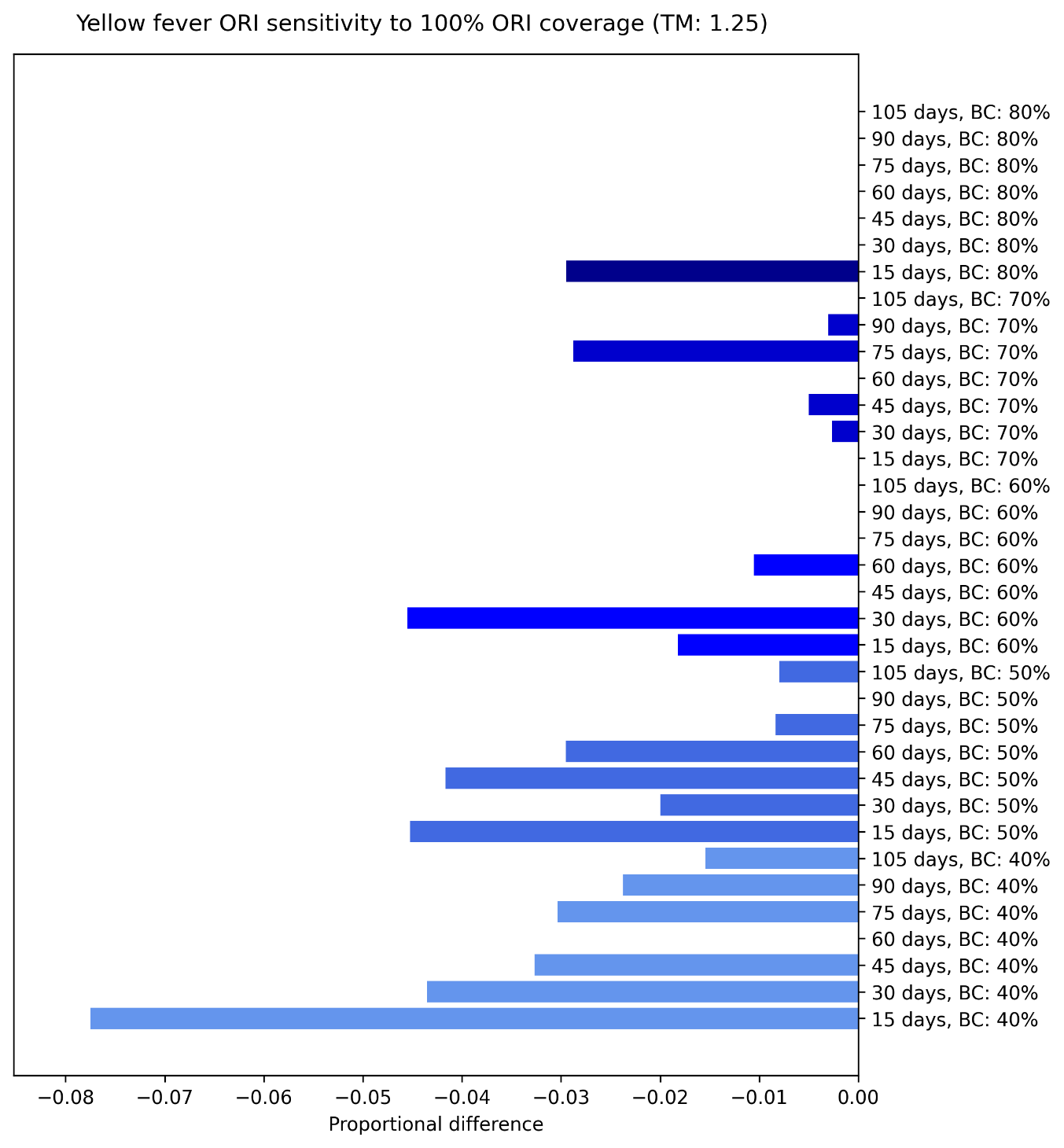

**Figure S39**: Proportional reduction in the mean cumulative cases and deaths for the set of outbreaks which received ORI that reached 100% of people, relative to the set of outbreaks which received ORI that reached 75% of people, estimated using bootstrap resampling and assuming a transmission modifier of 0·5 for the mosquito vectors. Proportional impacts are grouped vertically by routine vaccine coverage and ordered by ORI response time.

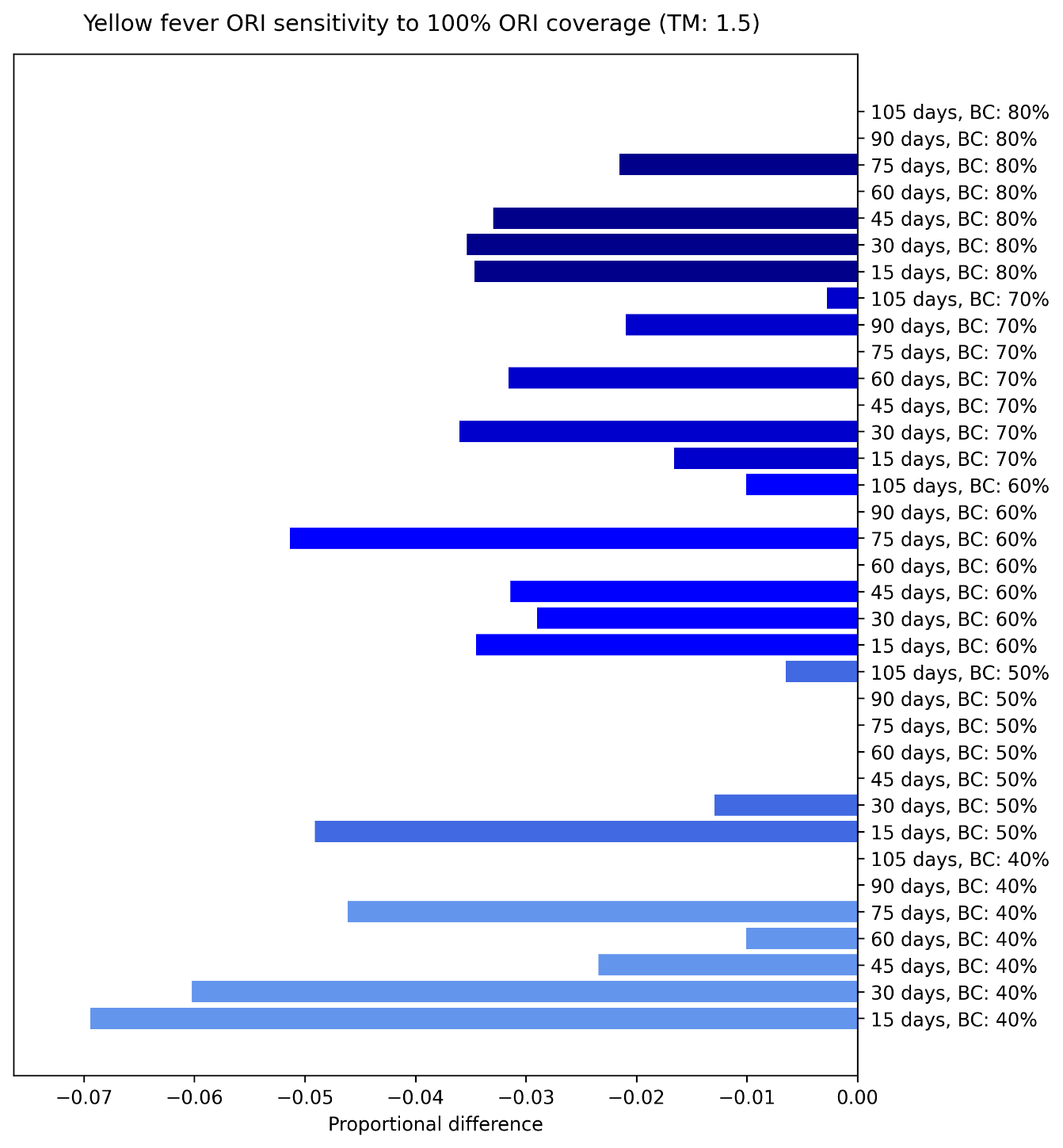

**Figure S40**: Proportional reduction in the mean cumulative cases and deaths for the set of outbreaks which received ORI that reached 100% of people, relative to the set of outbreaks which received ORI that reached 75% of people, estimated using bootstrap resampling and assuming a transmission modifier of 0·5 for the mosquito vectors. Proportional impacts are grouped vertically by routine vaccine coverage and ordered by ORI response time.

Increasing the achieved ORI coverage for yellow outbreaks from 75% to 100% in the target population was estimated to avert a minimal proportion of cumulative cases, unless ORI response times were very fast in high-risk setting archetypes (i.e., low routine vaccine coverage and high transmission modifier). In general, there was greater impact when ORI response times were faster, but stochastic effects could sometimes dominate within a given setting archetype.

#### 6.2 Alternate vaccination rate

In this section we present the proportional impact of varying the assumption for daily vaccination rate of the ORI for outbreaks of meningococcal meningitis, cholera, measles, and yellow fever, disaggregated by ORI response time and setting archetype. The sensitivity of our models to the slower vaccination rate seems to largely follow the trends in the main impact results (Figure 3 to Figure 6). We saw an increase in impact with ORI response time up to a point (e.g., 90 or 75 days for measles outbreaks) and then a plateau or decrease, which matches with the diminishing returns we saw with ORI response time in general, and was more pronounced for measles and yellow fever than cholera and meningococcal meningitis.

The results of the daily vaccination rate sensitivity analysis can be seen in Figures S41 – S48.

##### 6.2.1 Meningococcal meningitis

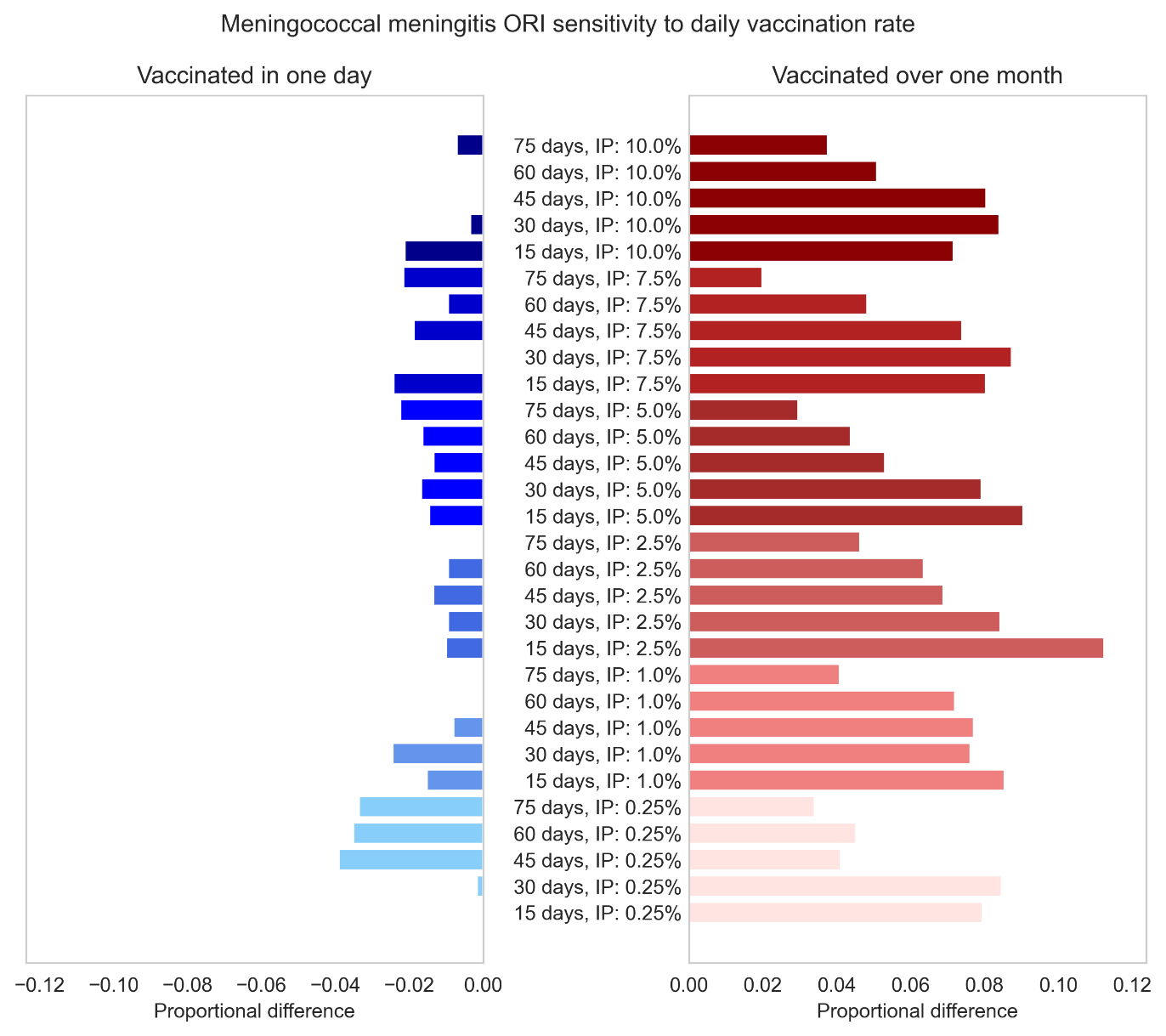

**Figure S41**: Proportional impact of simulating meningococcal meningitis ORI which vaccinates the target population (75% of people aged 1 – 29 years) in one day or over one month, compared to the Baseline assumption of approximately one week. Impacts are disaggregated by the response time of the ORI and the initial prevalence (IP) used for the outbreak simulations.

##### 6.2.2 Cholera

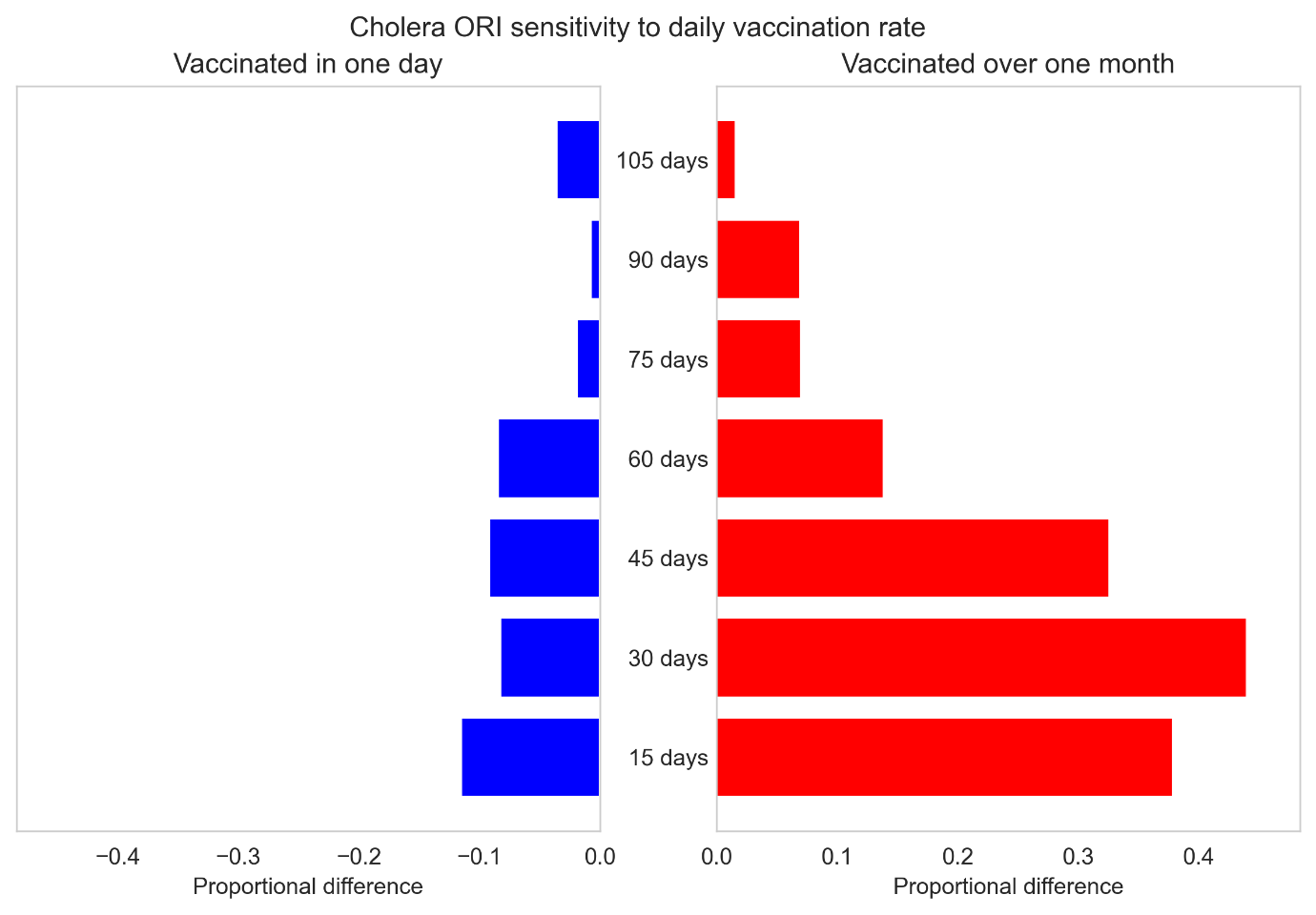

**Figure S42**: Proportional impact of simulating cholera ORI which vaccinates the target population (75% of people aged over one years) in one day or over one month, compared to the Baseline assumption of approximately one week. Impacts are disaggregated by the response time of the ORI used for the outbreak simulations.

##### 6.2.3 Measles

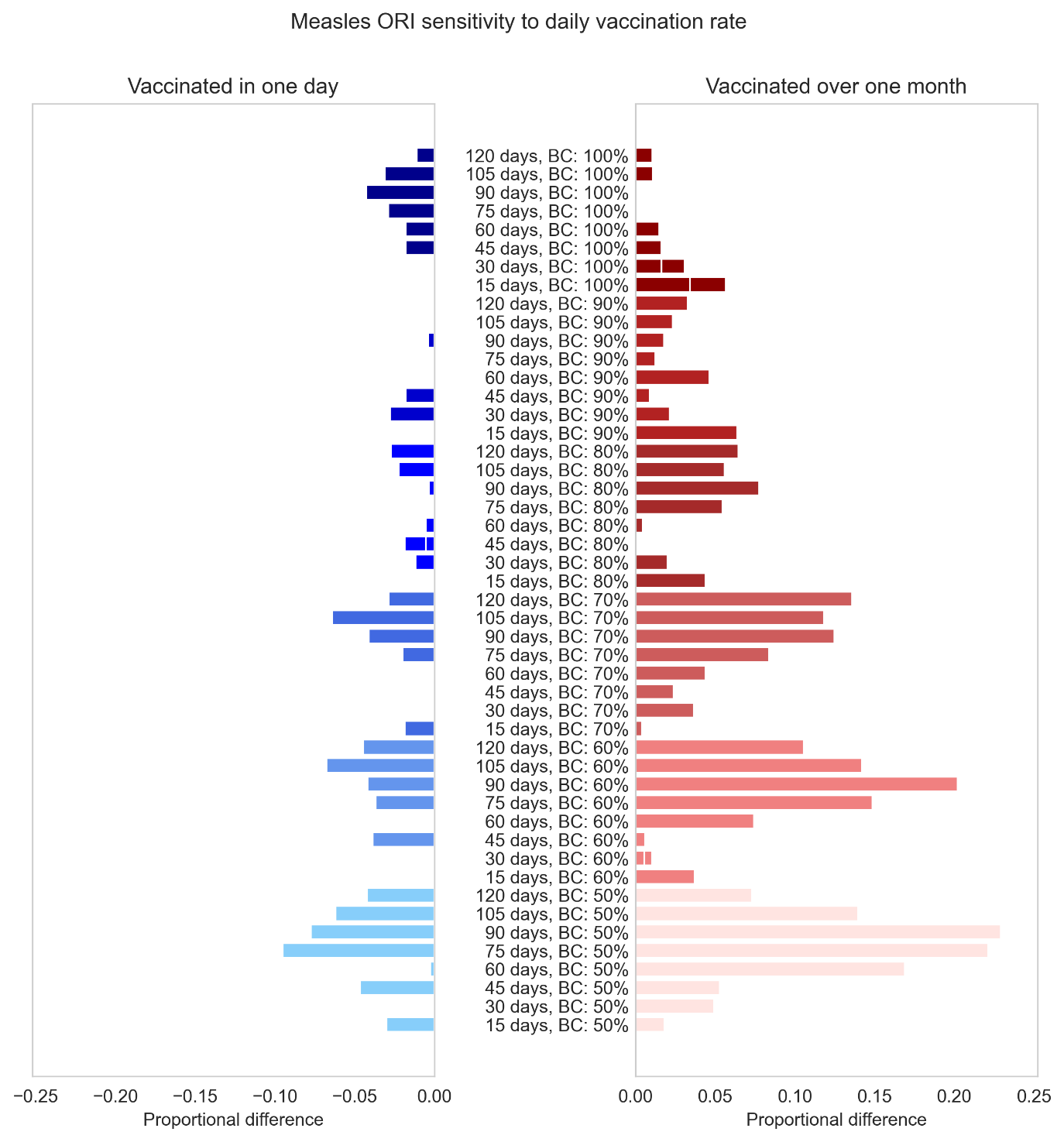

**Figure S43**: Proportional impact of simulating measles ORI which vaccinates the target population (100% of people aged nine months to five years) in one day or over one month, compared to the Baseline assumption of approximately one week. Impacts are disaggregated by the response time of the ORI and the routine vaccine coverage (BC) used for the outbreak simulations.

##### 6.2.4 Yellow Fever

The yellow fever simulations were very sensitive to the slower vaccination rate for all combinations of routine coverage and transmission modifier, with outbreaks with the highest transmission modifier and lowest routine vaccine coverage being up to three times larger when the rollout took one month (Figure S44 to Figure S48). The yellow fever simulations were also quite sensitive to the faster vaccination rate for all combinations of routine coverage and transmission modifier greater than 0·5, with outbreaks with the highest transmission modifier and lowest routine vaccine coverage being up to 40% smaller than the Baseline assumption of one week.

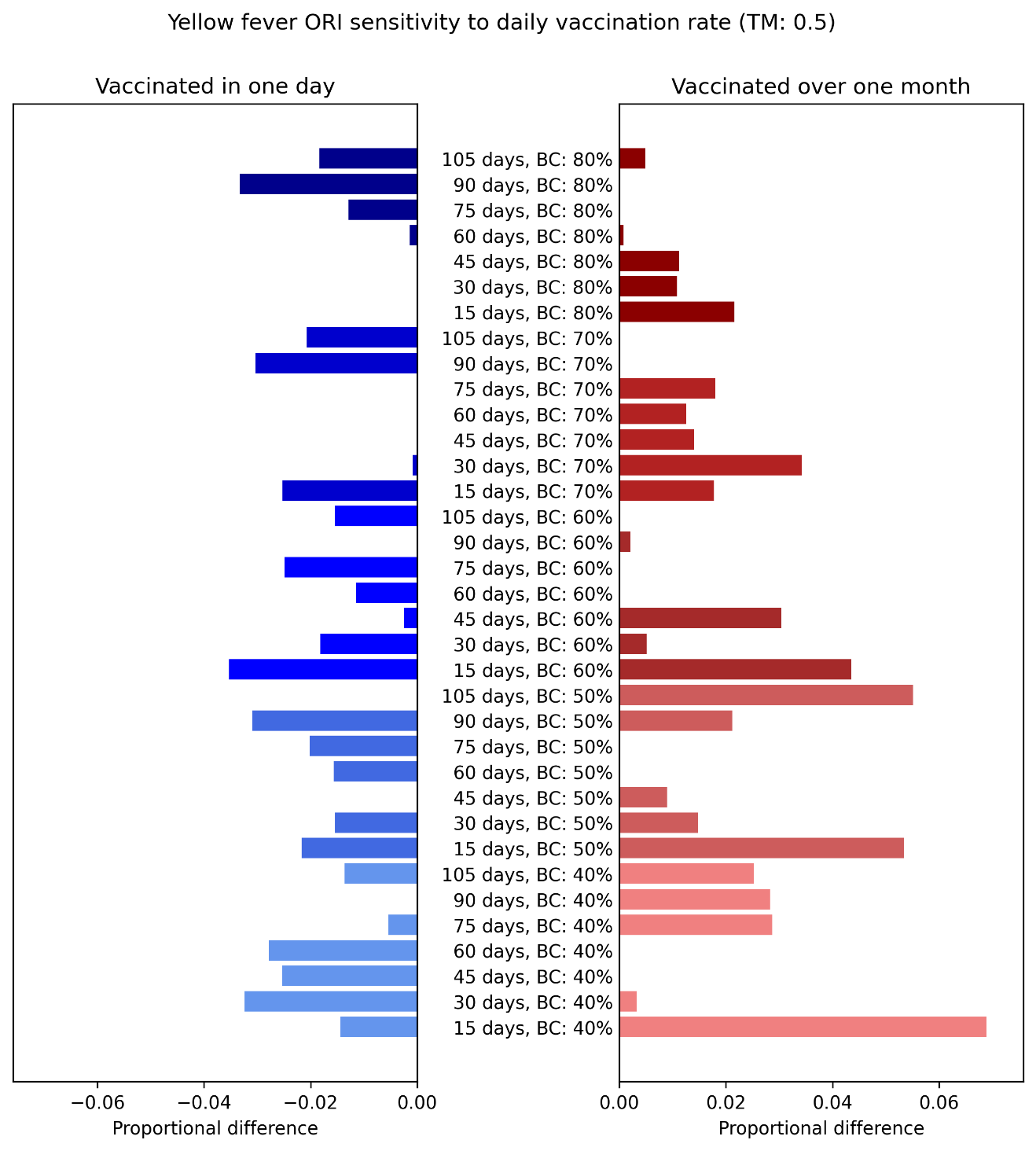

**Figure S44**: Proportional impact of simulating yellow fever ORI which vaccinates the target population (75% of people aged over nine months) in one day or over one month, compared to the Baseline assumption of approximately one week. Impacts are disaggregated by the response time of the ORI and the routine vaccine coverage (BC) used for the outbreak simulations, assuming a modifier of 0·5 for the mosquito to human transmission.

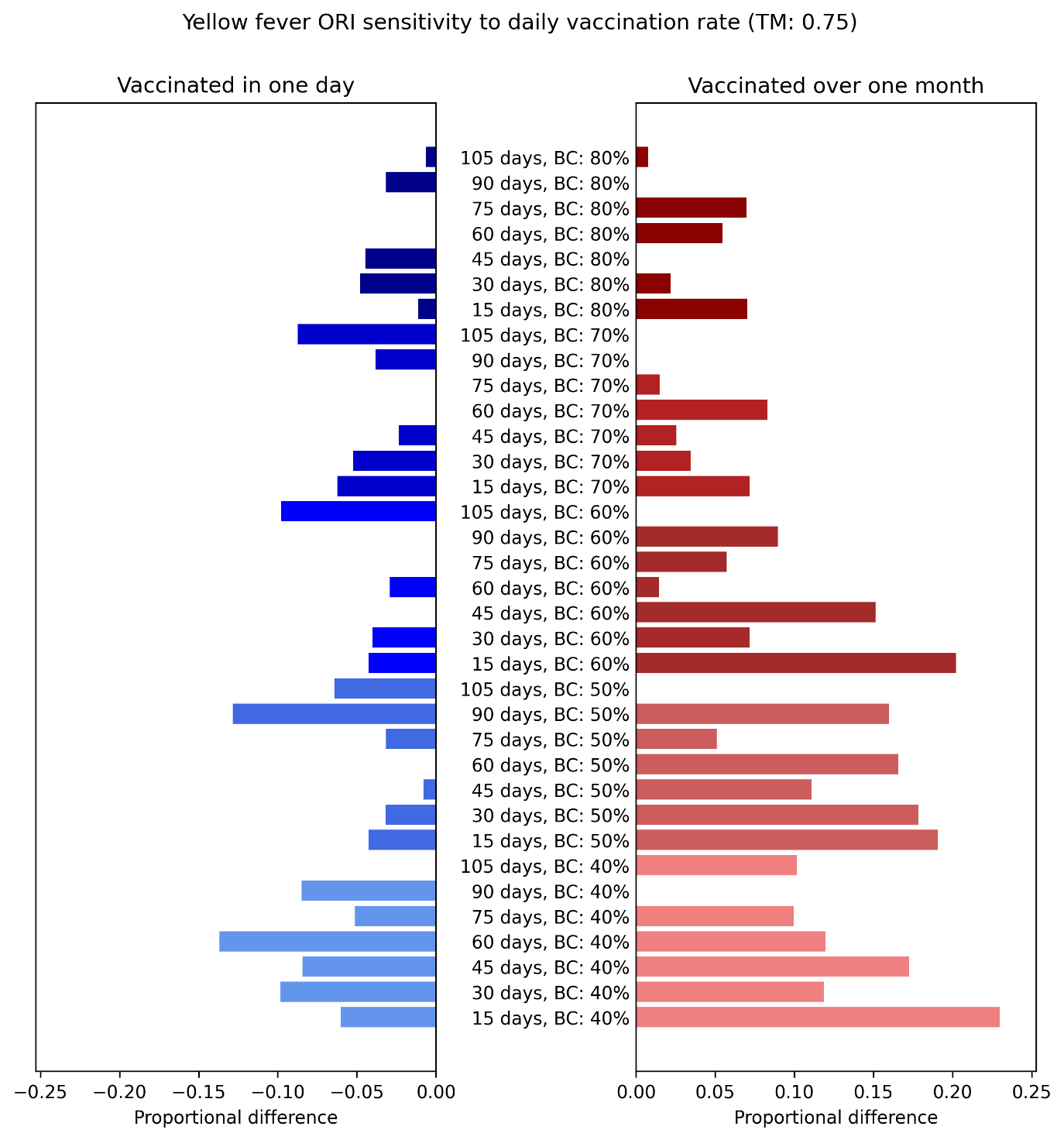

**Figure S45**: Proportional impact of simulating yellow fever ORI which vaccinates the target population (75% of people aged over nine months) in one day or over one month, compared to the Baseline assumption of approximately one week. Impacts are disaggregated by the response time of the ORI and the routine vaccine coverage (BC) used for the outbreak simulations, assuming a modifier of 0·75 for the mosquito to human transmission.

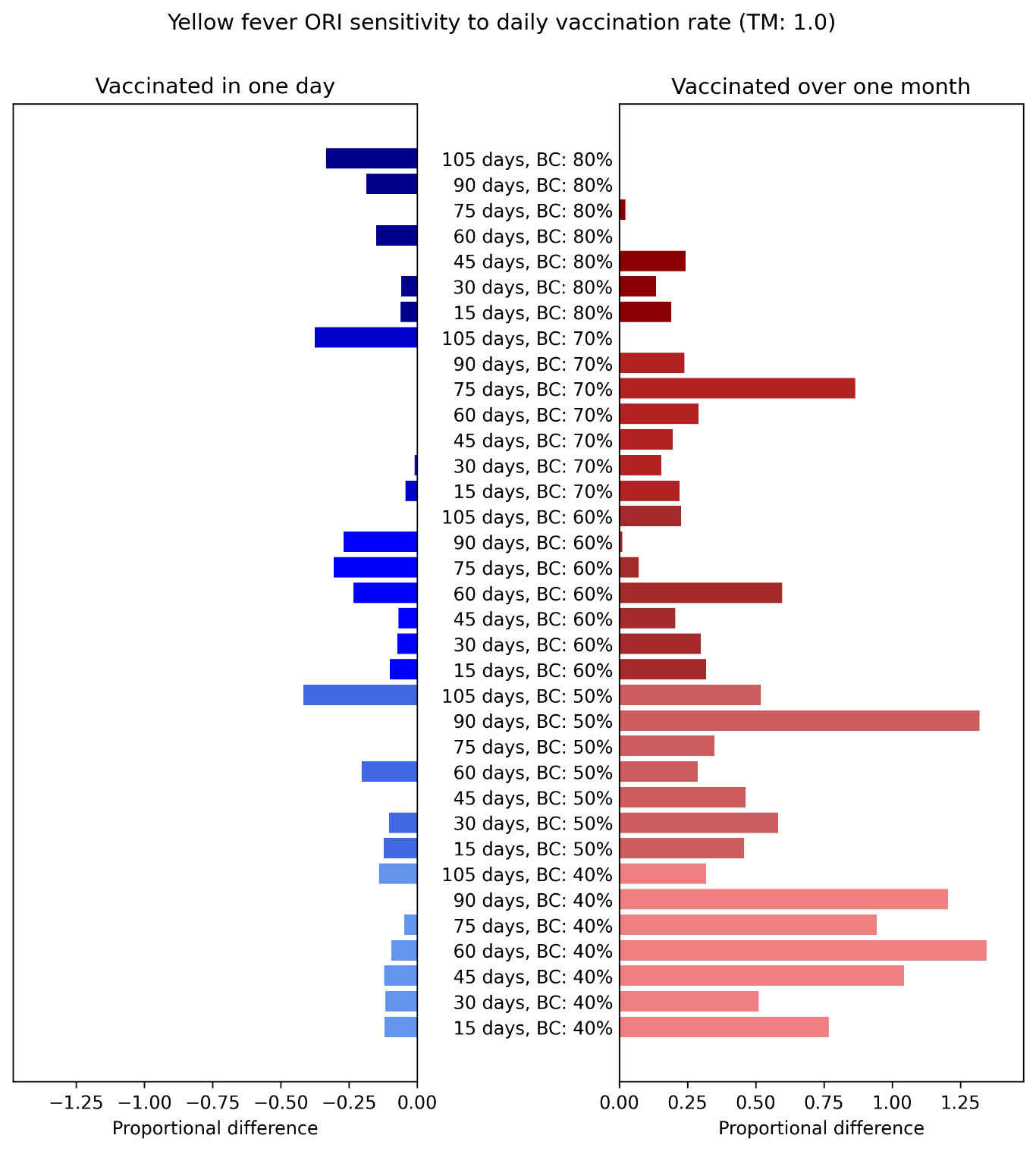

**Figure S46**: Proportional impact of simulating yellow fever ORI which vaccinates the target population (75% of people aged over nine months) in one day or over one month, compared to the Baseline assumption of approximately one week. Impacts are disaggregated by the response time of the ORI and the routine vaccine coverage (BC) used for the outbreak simulations, assuming a modifier of 1·0 for the mosquito to human transmission.

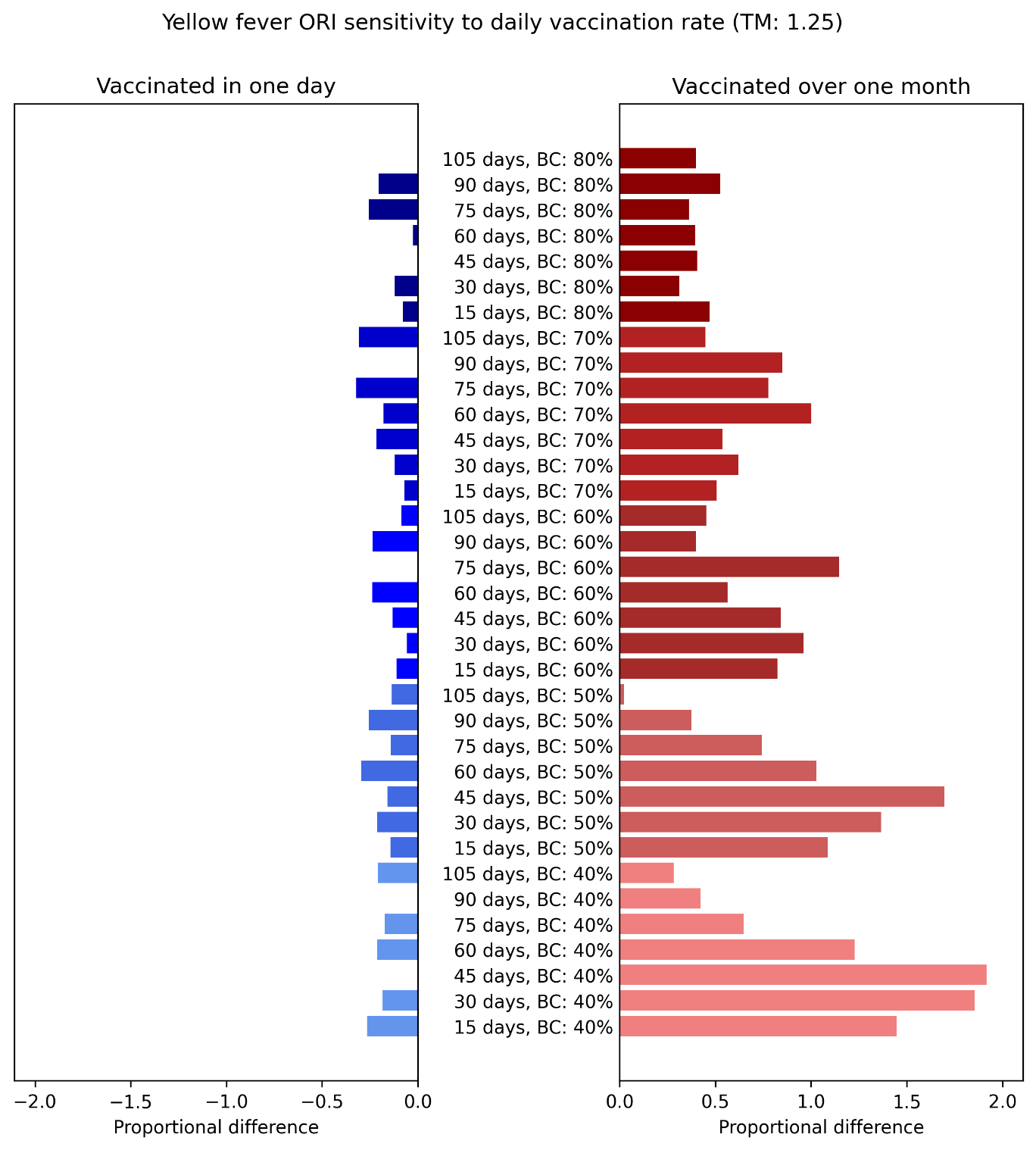

**Figure S47**: Proportional impact of simulating yellow fever ORI which vaccinates the target population (75% of people aged over nine months) in one day or over one month, compared to the Baseline assumption of approximately one week. Impacts are disaggregated by the response time of the ORI and the routine vaccine coverage (BC) used for the outbreak simulations, assuming a modifier of 1·25 for the mosquito to human transmission.

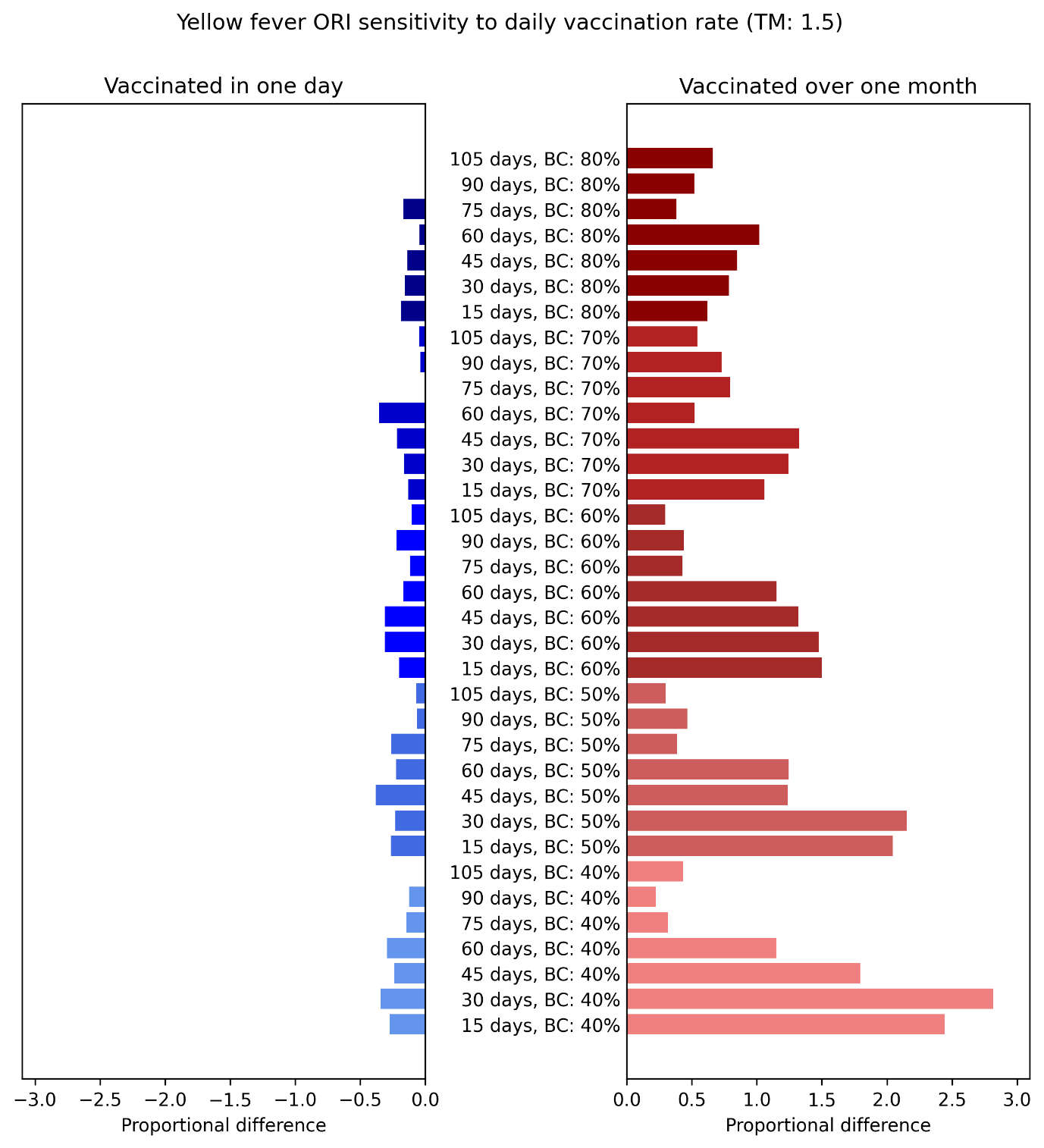

**Figure S48**: Proportional impact of simulating yellow fever ORI which vaccinates the target population (75% of people aged over nine months) in one day or over one month, compared to the Baseline assumption of approximately one week. Impacts are disaggregated by the response time of the ORI and the routine vaccine coverage (BC) used for the outbreak simulations, assuming a modifier of 1·5 for the mosquito to human transmission.
